# Advance Warning and Response Systems in Kenya: A Scoping Review

**DOI:** 10.1101/2025.04.23.25326250

**Authors:** Lisa M. Were, Jenifer A. Otieno, Moriasi Nyanchoka, Perpetua W Karanja, Dalmas Omia, Philip Ngere, Eric Osoro, M. Kariuki Njenga, Mercy Mulaku, Isaac Ngere

**Affiliations:** Research Department, Horn Population Research & Development, Nairobi, Kenya; Malaria Branch, Centre for Global Health Research, Kenya Medical Research Institute, Kisumu, Kenya; Health Economics Research Unit, KEMRI Wellcome Trust Research Programme, Nairobi, Kenya; Department of Health, Kirinyaga County, Kerugoya, Kenya; Institute of Anthropology, Gender and African Studies, University of Nairobi, Nairobi, Kenya; Washington State University Global Health Program, Nairobi, Kenya; Paul G Allen School of Global Health, Washington State University, Pullman, US; Department of Pharmacology, Clinical Pharmacy and Pharmacy Practice, Faculty of Health Sciences, University of Nairobi, Nairobi, Kenya

## Abstract

**Introduction:** Infectious diseases (IDs) cause approximately 13.7 million deaths globally. The Kenyan Advance Warning and Response Systems (AW&RS) against ID outbreaks is a core capacity of the 2005 International Health Regulations and a key indicator of health security. We mapped evidence on Kenya’s AW&RS and their enablers, and barriers for successfully detecting IDs, including climate-sensitive IDs.

**Methods:** We searched Cochrane Library, MEDLINE, EMBASE, Web of Science, Africa Index Medicus, and SCOPUS before August 26th, 2024. We also searched for grey literature on the Google Scholar search engine alongside the main repositories of Kenyan Universities. Two independent reviewers conducted study selection, while one reviewer extracted data. Discrepancies were resolved through discussion. Results were synthesised narratively and thematically.

**Results:** The search yielded 4,379 records from databases and 1,363 articles from websites, university repositories, and citations; we included 166 articles in the analysis. Integrated Disease Surveillance and Response (IDSR) and cohort surveillance systems were the most common (37.2%). Most studies were concentrated in Nairobi County (25.7%) and reported on malaria (23.6%). Most systems (82.4%) monitored the disease burden and outbreaks using hospital-based data (35.1%) and automated alert mechanisms (27.7%). National bulletins report a temporal association between environmental factors and disease prevalence. Malaria, Rift Valley Fever (RVF), and cholera cases increased with higher precipitation, lower temperatures and increased vegetative index. AW&RS used the accuracy and reliability of the model prediction to measure the system’s performance. Effectiveness was evaluated based on system acceptability and timeliness. Health system factors were predominant, with 121 enablers and 127 barriers. Key enablers included skilled personnel (13 studies), whereas inadequate finances were a major barrier (21 studies).

**Conclusion:** Most AW&RS were IDSR and cohort-based surveillance. Climate changes have resulted in observed trends in diseases such as malaria and RVF, but further studies are needed to determine causal links. Insufficient funding hinders the effective implementation of AW&RS. Future research should assess the cost drivers influencing system effectiveness.

## Background

Advance Warning and Response Systems (AW&RS) play a crucial role in timely monitoring, prevention and response to disease outbreaks. Infectious diseases (IDs) remain a significant public health concern in Africa, accounting for 20% of the global burden of disease in 2023^1^. In Kenya, IDs are the primary cause of hospital admissions and outpatient clinic visits, surpassing non-communicable diseases ^2^.

The East African region faces ongoing challenges from endemic and emergent infections, which pose serious threats to both people and health systems^3^. With more than 75% of IDs of public health concern being zoonotic, there is a growing focus on animal and human health ^4^. Furthermore, climate change significantly affects the seasonality and distribution of climate-sensitive infectious diseases (CSIDs)^5,6^. The One-Health Approach has gained prominence in addressing these complex health challenges. This approach examines the interplay between humans, animals and their shared environment, to optimise resource allocation at all levels for better health outcomes ^4^.

Public health emergencies due to IDs pose significant threats to population health and healthcare systems ^7–9^. To mitigate these risks, AW&RS are crucial for early detection and rapid intervention. These systems provide timely alerts to potential acute public health epidemics and facilitate immediate public health response^10^. The AW&RS typically focuses on detecting diseases with a potentially high disease burden (for example, cholera), emerging or re-emerging infectious/zoonotic diseases (for example, anthrax) and diseases targeted for eradication (for example, poliomyelitis)^8–10^.

In most African countries, the national public health authorities lead the implementation of the AW&RS, with financial and/or technical support from donors or non-governmental organisations ^10^. Kenya has made strides in enabling emergency preparedness units by investing in local capacity and infrastructure for AW&RS ^11–13^. Kenya adopted the Integrated Disease Surveillance and Response (IDSR) strategy in 2006. This system, initially developed by the World Health Organization (WHO), is designed to facilitate early detection and rapid response to 36 diseases of public health concern ^14^.

Kenya has significantly improved its health surveillance system over the past two decades. It transitioned from manual reporting to a desktop-based surveillance system in 2008, followed by a web-based electronic IDSR system (e-IDSR) in 2011^4^. In 2016, Kenya upgraded the IDSR reporting to the District Health Information System 2 (DHIS 2), enabling comprehensive data monitoring across different health programs^4^. The system later evolved to the more integrated Kenya Health Information System (KHIS). In response to the Coronavirus disease 2019 (COVID-19) pandemic, Kenya implemented an event-based surveillance (EBS) for rapid threat detection ^12^. The Ministry of Health has invested in training and equipment to enhance timely reporting and response capabilities and ensure the effective implementation of these advancements ^11^.

Despite these advancements, several barriers impede their implementation of these systems. Firstly, limited financial allocations from county governments and declining donor funding places challenges on their development and maintenance^4^. Secondly, inadequate infrastructure and technology, particularly in remote areas, hinder widespread adoption and utilisation of these systems. Additionally, data management issues further complicate the situation, including lack of specialised laboratory equipment, outdated information systems, and limited computer and internet access^4^.

A global systematic review examined the effectiveness of AW&RS in detecting IDs in various settings. The review included 68 studies, categorising them into emergency care and triage-based systems (20 studies), hospital/public health record systems (13 studies) and web and internet-based systems (11 studies)^4^. Despite this comprehensive analysis, the review did not capture any Kenyan literature, highlighting a significant gap in the current understanding of AW & RS implementation in Kenya.

Although multiple primary studies and reports indicate the presence and development of AW&RS in Kenya, no comprehensive review has systematically mapped their scope and effectiveness^4,11^. This gap in knowledge is particularly noteworthy as the Kenyan AW&RS landscape continues to evolve. A systematic examination of Kenya’s progress in this field could provide valuable insights to both national and global stakeholders. Given this context, this study aimed to systematically map the AW&RS landscape in Kenya, and explore enablers, and barriers affecting its effectiveness of these systems in detecting IDs, including CSIDs. This study seeks to contribute to the growing body of knowledge on AW&RS and provide a foundation for improving ID detection and response.

## Methods

We conducted this review according to the Arksey and O’Malley scoping review framework^15^. The steps of the framework are: (i) identifying the research question, (ii) identifying relevant studies, (iii) study selection, (iv) charting the data, (v) collating, summarising, and reporting the results, and. Our protocol was deposited in the Open Science Framework (OSF) registry ^16^.

### Identifying the research question

The overall review question was, “What is the available evidence on the AW&RS, their enablers, and barriers in detecting infectious and climate-sensitive diseases in Kenya?”

The specific review questions were:

1. What evidence exists regarding AW&RS for detecting infectious and climate-sensitive diseases currently in use at the national and subnational level?
2. What are the effects of the current AW&RS on ID risk and health outcomes?
3. What is the impact of climate on health and existing climate-sensitive diseases?
4. What are the enablers and barriers to successfully implementing the AW&RS to wastewater management and IDs risk?
5. What are the cost drivers for AW&RS in Kenya?

### Eligibility Criteria

The Population, Concept, and Context (PCC) framework guided our study eligibility criteria below ^17^;

***Population:*** Studies on humans, livestock, wildlife, the environment, and wastewater management.

***Concept:*** Studies on AW&RS and climate impact on IDs, including CSIDs, examining their enablers, barriers, and cost drivers.

***Context:*** all healthcare levels and settings (rural and urban) across Kenyan counties where the AW&RS have been utilised for ID outbreaks, CSIDs and potential pandemics.

**Types of Studies:** We included primary studies in English or with an English translation, encompassing quantitative, qualitative, and mixed-methods approaches and analytical and descriptive studies, relevant policies, guidelines, and reports from governmental and non-governmental bodies. We excluded commentaries, case series, and opinion papers because of their potential bias, and pilot studies and reviews to minimise data duplication.

### Identifying relevant studies

with the aid of an information specialist, we conducted a literature search on the following electronic databases from 1946 to August 26^th^, 2024: Cochrane Library, MEDLINE (OVID), EMBASE, Web of Science (all databases), Africa Index Medicus, SCOPUS (Elsevier), and CINAHL (via EBSCO host).

We also searched for grey literature on the Google Scholar search engine and the following websites: WHO sources and Centers for Disease Control (CDC) Sources, Ministry of Environment and Forestry, Kenya Meteorological Department (KMD), Kenya National Drought Management Authority, and Enhancing National Climate Services (ENACTS). We sought additional grey literature on online repositories of Kenyan universities (n=20) offering health-related courses.

We also searched the reference lists of included studies and systematic reviews. We used the following key search terms alongside their synonyms: “early warning systems,” “wastewater system,” “infectious disease,” “climate-sensitive diseases,” “enabler,’’ barrier,” and “Kenya.” The full search strategy for MEDLINE (OVID) is provided under **<u>Annex 1</u>**.

### Study Selection

Two independent reviewers screened the titles, abstracts and full texts of all eligible studies against the eligibility criteria using the Covidence® platform—a systematic review management software and settled all discrepancies by discussion to achieve consensus ^18^. The average Inter-rater reliability at the title and abstract screening stage and full-text review stage was a Cohen’s Kappa statistic of 0.3732 and 0.3493, respectively, indicating a fair agreement between reviewers ^19^.

### Charting the Data

Two independent reviewers piloted the pre-designed data extraction form on five eligible studies, and necessary modifications were made to the form. The data extraction form is available under **<u>Annex 2</u>** ^16^. One reviewer per article used the modified data extraction form in Covidence to extract data independently. The senior reviewers were consulted to resolve reviewers’ discrepancies on extraction.

We charted the following data items;

**General study details:** Study title, lead author’s surname, year of publication, study objective(s), study setting, and study design;

**Characteristics according to the PCC framework:** participant details, type of AW&RS, the enablers and barriers to AW&RS activities according to the Supporting the Use of Research Evidence (SURE) framework, lessons learned, dates of implementation, and information regarding study funding ^20^.

We used the SURE framework to systematically categorise the enablers and barriers to the effective implementation of AW&RS^20^. The SURE framework categorises factors into five groups: recipients of care, providers of care, other stakeholders, health systems, and social and political constraints ^20^.

### Collating, Summarizing, and Reporting the Results

We summarised the extracted data using descriptive statistics for the quantitative data using Microsoft Excel software Version 2021^21^. We presented the key findings using tables, graphs, and charts. We have reported the results per the Preferred Reporting Items for Systematic Reviews and Meta-Analyses for Scoping Reviews (PRISMA-ScR) guidance ^22^.

## Results

### Study selection

We identified 4379 records through electronic database searching. After removing 1539 duplicates, we screened 2840 titles and abstracts and excluded 2522 irrelevant reports. We retrieved 318 reports for full-text review, excluded 169 based on eligibility criteria, and included 137 studies in the scoping review (Figure 1). We also identified studies via other sources, including 1,363 articles from websites, academic institution repositories, and citation searching. After removing duplicates, we sought 1361 reports for retrieval, two of which were not retrieved. We assessed 1359 articles for eligibility and excluded 1330 studies. We included 29 reports from other sources, bringing the total articles to 166.

**Figure 1:**
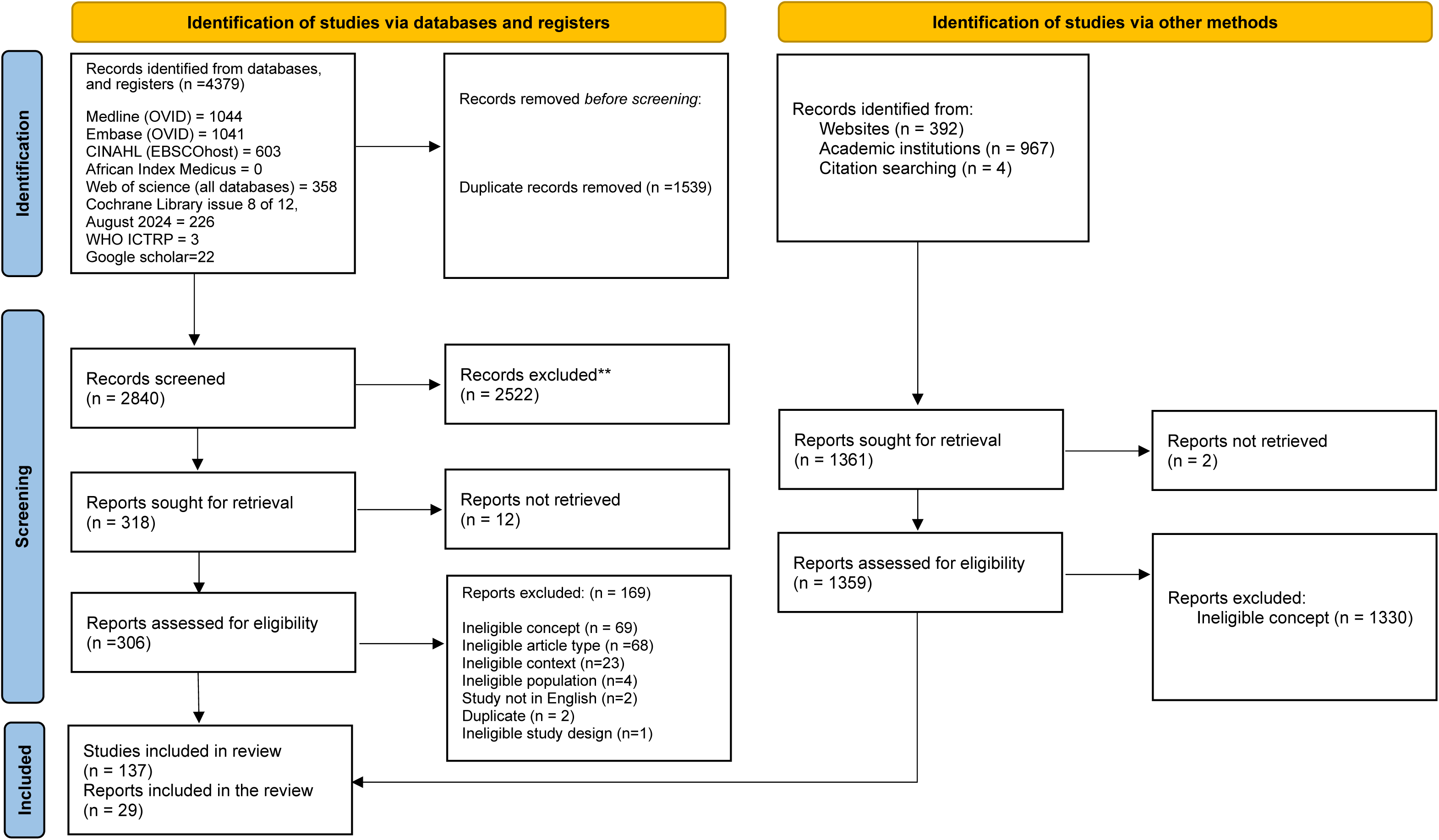
PRISMA 2020 flow diagram of included studies and reports.

### General characteristics of included studies

Of the 166 included articles, 75.3% (n=125/166) were journal articles, 7.2% (n=12/166) were university theses, 6.6% (n=11/166) were NGO reports, and 10.8% (n=18/166) were national climate bulletins sourced from relevant website searches (Table 1). The 18 articles on climate data, comprising bulletins, are summarised separately in the supplementary file 1. Of the 148 articles, more than half (55.4%, n=82/148) used observational designs, while 12.2% (n=18/148) used secondary data analysis.

**Table 1:** Characteristics of Included Articles.

| Study characteristics | No of studies (%) |
| --- | --- |
| **Publication year** |  |
| Before 2000 | 2/166 (1.2%) |
| 2000-2010 | 15/166 (9.0%) |
| 2011- 2020 | 95/166 (57.2%) |
| 2021- August 2024 | 54/166 (32.5%) |
| **Geographical locations** |  |
| Counties (with over 10 systems) | Nairobi 11.73% (38/324); Kilifi 7.72% (25/324); Siaya 7.41% (24/324); Kisumu 6.79% (22/324); Kwale 3.70% (12/324); Mombasa 3.70% (12/324); Baringo 3.40% (11/324); Tana River 3.40% (11/324); Garisa 3.40% (11/324); Nakuru 3.09% (10/324) |
| **Participants** |  |
| Human participants only | 118/148 (79.7%) |
| Humans and animals | 11/148 (7.4%) |
| Environment only | 7/148 (4.7%) |
| Animals only | 5/148 (3.4%) |
| Climate only | 4/148 (2.7%) |
| Humans and climate | 2/148 (1.4%) |
| Animal and climate | 1/148 (0.6%) |
| Humans and environment | 1/148 (0.6%) |
| **Scope of AW&RS systems** |  |
| Disease burden and outbreak surveillance | 122/148 (82.4%) |
| Disaster preparedness | 3/148 (2.0%) |
| Disease burden, outbreak surveillance, and disaster preparedness | 6/148 (4.1%) |
| Improved forecasting | 2/148 (1.4%) |
| **AW&R System (Name)** |  |
| Integrated Disease Surveillance and Response (IDSR) (Most frequently mentioned surveillance system) | 18/148 (12.2%) |
| Population-Based Infectious Disease Surveillance (PBIDS) /Cohort Surveillance | 12/148 (8.1%) |
| Health and Demographic Surveillance System (HDSS)/ Cohort Surveillance<br>(Includes Kilifi HDSS, Nairobi Urban HDSS, Kombewa HDSS, Rusinga HDSS, etc.) | 14/148 (9.5%) |
| Other cohort surveillance systems | 11/148 (7.4%) |
| Sentinel Surveillance (Includes antenatal care (ANC) sentinel, Human Immunodeficiency Virus (HIV) sentinel, influenza sentinel, polio/Acute Flaccid Paralysis (AFP) surveillance, etc.) | 13/148 (8.8%) |
| Modelling / Early Warning Systems (Statistical modelling, RVF prediction models, malaria early warning, etc.) | 14/148 (9.4%) |
| Malaria Surveillance<br>(Malaria Early Warning, passive surveillance, vector surveillance, etc.) | 10/148 (6.8%) |
| Digital Surveillance (mHealth)<br>(Short Message Service (SMS) for Life, mSOS, mobile-based systems, Early Warning Alert and Response System (EWARS), etc.) | 8/148 (5.4%) |
| Wastewater/Environmental Surveillance<br>(Bag-mediated filtration system (BMFS), polio environmental surveillance, etc.) | 6/148 (4.1%) |
| Community-Based Surveillance<br>(Rabies, RVF, syndromic surveillance, etc.) | 6/148 (4.1%) |
| Polio (AFP) Surveillance | 9/148 (6.1%) |
| Influenza Surveillance | 5/148 (3.4%) |
| Zoonotic/Animal Surveillance | 5/148 (3.4%) |
| Research Studies (Unspecified) | 10/148 (6.8%) |
| Not Reported / Unclear | 7/148 (4.7%) |
| Other Systems (Neglected Tropical Diseases (NTDs), Tuberculosis Bacillus (TB), Cholera, etc.)<br>(Includes DHIS2, rotavirus surveillance, enteric surveillance, etc.) | 13/148 (8.8%) |

Most studies were conducted in five Kenyan counties: Nairobi (25.7%, n=38/148); Kilifi (16.9%, n=25/148); Siaya (15.5%, n=23/148); Kisumu (14.9%, n= 22/148), with Mombasa and Kwale having a similar number of studies (8.1%, n=12/148). Twenty-two percent (n=33/148) of the studies were conducted in rural areas, 27.0% (n=40/148) in urban areas, and 7.4% (n=11/148) in both rural and urban settings. Most systems are implemented at the county level only (35.8%, 53/148), while systems that were internationally applied comprised 8.7% (n=13/148). Most systems (79.7%, n=118/148) included human participants only; 4.7% (n=7/148) focused on the environment only.

Most studies reported system enablers and barriers (64.9%, n=96/148). Health system-level enablers (n=121 enablers) and barriers (n=127 enablers) were the most reported, with fewer studies reporting on the stakeholder’s/actor’s level enablers (n=8) and barriers (n=2). No study explicitly reported the costs of the enablers and barriers to successfully implementing the AW&RS.

## Synthesis of findings

### 1. Available evidence on AW&RS

#### Scope of AW&RS in Kenya

The included studies reported different types of AW&RS. Most were human disease surveillance systems (n=122/148). Some systems combined human disease surveillance with surveillance for animal diseases and syndromes (8.1%, n=12/148), while a smaller proportion focused exclusively on animal diseases and syndromes (5.4%, n=8/148) or environmental surveillance (4.1%, n=6/148). Cohort surveillance systems and IDSR are the most common AW&RS in Kenya, comprising 37.2% of the systems (**Table 1**). Modelling (14/148 (9.4%)) and digital surveillance (8/148 (5.4%)) are emerging trends. Polio (AFP) 9/148 (6.1%), malaria 10/148 (6.8%), and influenza 5/148 (3.4%) have dedicated disease specific surveillance systems. The systems predominantly purposed to monitor disease burden and outbreak surveillance (82.4%, n=122/148). A smaller proportion focused on integrated disease burden, outbreak surveillance, and disaster preparedness (4.1%, n=6/148).

The top diseases reported were malaria (n=35/148, 23.6%), RVF (12.2%, n=18/148), Human Immunodeficiency Virus and Acquired Immunodeficiency Syndrome (HIV/AIDS) (11.5%, n=17/148) and COVID-19 (8.8%, n=13/148) (Table 2). Regarding broader ID categories, six studies (4.0%, n=6/148) reported human enteric infections, and five studies (3.4%, n=5/148) reported zoonotic and IDSR priority diseases each. Eighty-five studies (57.4%, n=85/148) reported on CSIDs.

Some systems combined disease surveillance with improved forecasting or environmental monitoring. Systems focused solely on disaster preparedness (2.0%, n=3/148) or improving forecasting (1.4%, n=2/148) are less common. Seventy-six studies (51.3%) reported on the alert mechanisms within the AW&RS. Automated alerts such as Short Message Service (SMS)-based, event-based, and digitalised platforms were the most used (27.7%, n=41/148). Digital-based methods such as remote satellite sensing was mainly used for data collection, storage, reporting, and disease mapping. Manual notifications comprised 14.9% (n=22/148), while internal reports constituted 3.4% (n=5/148). A smaller portion of the systems use a combination of automated alerts and manual notifications (2.7%, n=4/148).

Of the 148 records, 131 studies reported the data sources. Most of the sources (35.1%, n=52/148) were from hospital-based data, followed by a combination of community and hospital sources (18.2%, n=27/148) and community sources alone (10.8%, n=16/148). A smaller portion of the data came from environmental sources (4.1%, n=6/148) and animal sources (4.1%, n=6/148) (Table 3).

##### Key performance indicators

More than half of the studies (66.2%, n=98/148) reported on key performance indicators (Table 4). Studies used diverse techniques to measure the system’s performance, such as the accuracy and reliability of the model predictions^23^, annual and super-annual variations in disease incidence^24^, capability to detect, confirm, report, analyse and interpret surveillance data^25–31^, data completeness^13,32–36^. Other systems focused on the reported proportion of death counts^37–40^. Other surveillance systems focused on measuring the feasibility of the systems, looking into their simplicity, acceptability, stability, usefulness, data quality and surveillance data reporting timeliness^25,41,42^. Further, some systems assessed performance using prompt immediate public health actions^47^, level of utilisation of the data produced by the surveillance system ^48^ and vaccination coverage^49^.

##### Lessons learnt

More than half of the studies (57.4%, n=85/148, 57.4%) reported lessons learnt from the systems’ operations (Table 4). The key lessons reported were using inexpensive platforms to encourage reporting, such as using a toll-free number which enhanced the efficiency of the reported cases^50^. They also learnt that integration of facility-based screening and community-based outreaches provided a comprehensive approach to addressing barriers and reaching underserved populations^51–54^ and the scope of the diseases under monitoring should be increased to achieve comprehensive surveillance^55^. Three systems reported that continuous surveillance which is crucial for early detection could be maintained using virtual remote supervision, thus enabling timely public health action^49,56,57^. Also, the studies reported that engaging with local stakeholders in the field and providing timely feedback through regular public engagement sessions are essential for ensuring ongoing compliance^58^. Effective surveillance, vaccination, and management strategies are needed to mitigate its impact on public health^59–61^. Effective supervision and monitoring of AFP surveillance are crucial for identifying, reporting, and investigating all actual AFP cases^62,63^. Studies reported that AW&RS should be designed to be flexible and responsive to changing environmental conditions, such as shifts in climate or land use, which can influence disease dynamics^40^. Additionally, passive surveillance was reported to often lead to incomplete data due to underreporting, inconsistencies in data collection, and reliance on healthcare providers to voluntarily submit reports^64,65^. From the findings, training staff on outbreak response is essential for strengthening public health systems and enhancing readiness for future outbreaks^66,67^. Using geo-referenced data for monitoring malaria prevalence allows for a better understanding of spatial and temporal patterns^68^.

##### Extent of Success

Most studies (60.1%, n=89/148) reported on the extent of success of the systems, (Table 4), while some (40.5%, n=60/148) did not report on it as summarised by the following themes.

###### System Coverage and Reporting Efficiency

The AW&R surveillance systems successfully collected data from both urban and rural settings, covering human and animal health, environmental variables, and disease outbreaks ^51,52,69–71^. A high volume of data was reported, including 20,340 animal and death events, 11,399 domestic and 205 wild animal disease reports via Kenya Animal Bio Surveillance system, and 8,734 event signals through Community Early Warning Surveillance ^39,72–74^. Completeness (87% of the reports) and timeliness (93%) of reporting were high, ensuring rapid response to public health threats ^34^.

###### Disease Detection and Monitoring

The AW&RS successfully identified disease outbreaks, including haemorrhagic fevers, cholera, rabies, polio and influenza ^75–79^. They enhanced early warning capabilities, predicting RVF outbreaks and using rainfall and temperature data for forecasting ^80–82^.

###### Public Health Impact and Response

The malaria surveillance system facilitated better malaria case detection using georeferenced data, enabling the identification of high-risk geographical hotspots^68^. Successful introduction of Piperonyl Butoxide-treated nets significantly reduced malaria incidence in targeted counties ^54^. The antenatal care surveillance system supported maternal and child health programs, ensuring continuity of care and adherence to Prevention of Mother-To-Child Transmission of HIVtesting ^52^. Contact tracing and prophylactic treatment were effectively used for cholera response, highlighting its role in epidemic control ^43^.

###### Technological and Data Innovations

Adoption of electronic data collection improved timeliness, accuracy, and cost-effectiveness ^34,35,83,84^. Cloud-based databases adopted by the Mobile EBS system enabled real-time reporting and analysis, improving response time ^72^. The metagenomics approach was used by the PBIDS for early pathogen detection before widespread outbreaks ^85^.

### 2. CSIDs and Climate Impacts on Health

According to the review findings, malaria and RVF were the most reported CSIDs in 35 and 18 studies, respectively (Table 2). Cholera and Anthrax ranked second, as reported in eight and seven studies. Ebola and Chikungunya were among the least reported CSIDS, captured in just two studies each. In most parts, the country experiences heavy rainfall between March to May, with peaks in April ^86,87^. <u>Tables 5.1, 5.2, 5.3, 5.4, and 5.5</u> present extensive information on the CSIDs which are summarised below;

Three bulletins found temporal associations between the occurrence of low Rainfall/drought conditions, high temperatures, reduced vegetative index, and occurrences of IDs such as anaplasmosis, babesiosis, East Coast Fever (ECF) and abortions among goats due to suspected Q-Fever disease in different regions of the country, such as Samburu Embu, West Pokot, and Narok counties ^87,88^. However, one bulletin reported low disease outbreaks during the dry months of February and March 2022 in Kakamega, Kisii, and Nandi counties^89^.

On the contrary, nine articles reported that high Rainfall/flooding, cold temperatures and increased vegetative index saw an Increased incidence of diseases such as cholera, respiratory, waterborne and livestock diseases; ECF, Contagious Caprine Pleuropneumonia (CCPP), Contagious Bovine Pleuropneumonia (CBPP), Peste des Petits Ruminants (PPR) and Foot and Mouth Disease ^24,40,80–82,89,90^.

#### Barriers and enablers to effective AW&RS

Of the 148 studies, 52 (35.1%) reported no enablers or barriers to AW&RS implementation (Table 6). Among those reporting, health system-level factors were the most frequent (121 enablers, 127 barriers), whereas stakeholder’s level factors were infrequent (8 enablers,2 barriers) (Figure 2).

**Figure 2:**
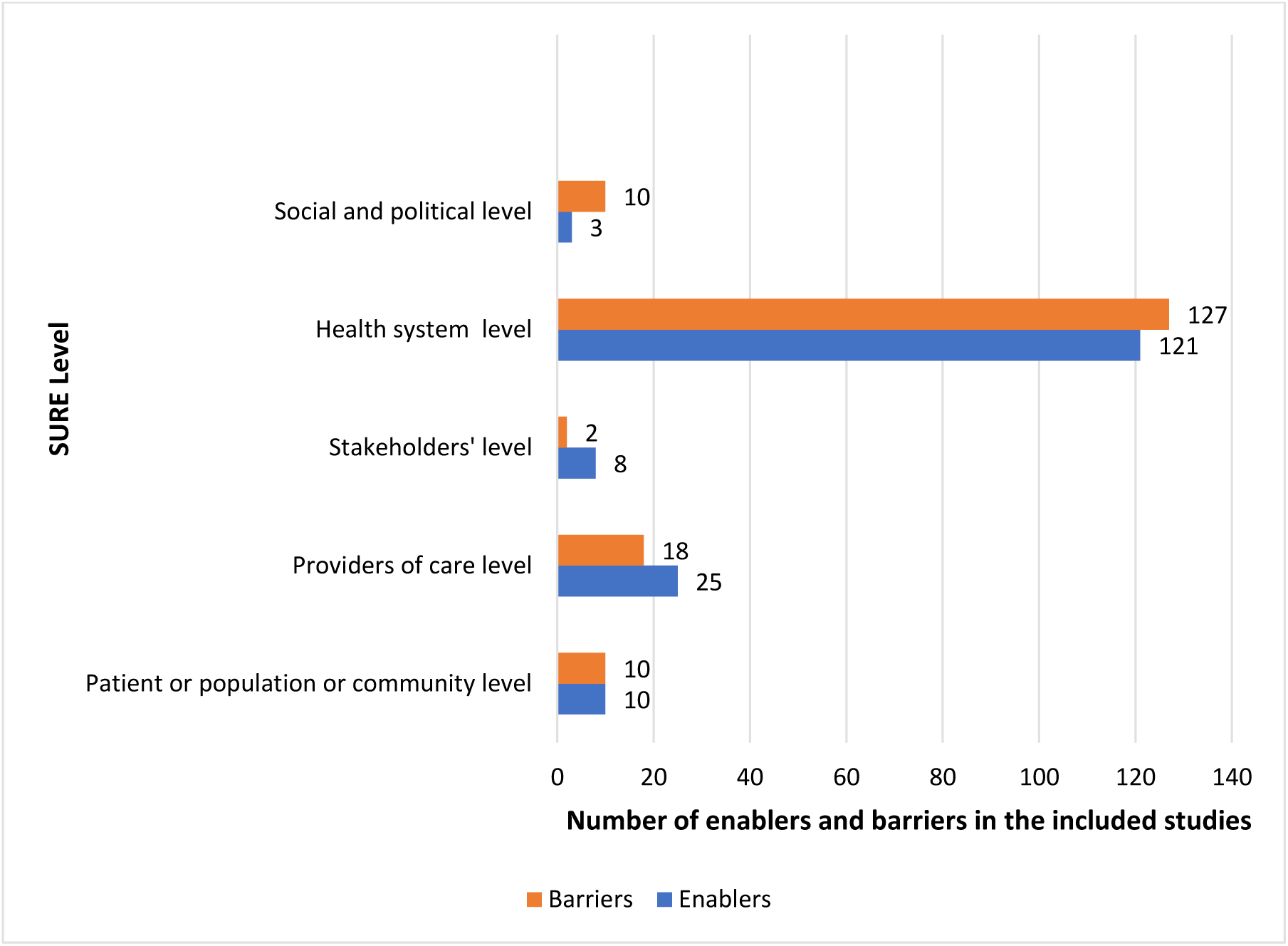
Enablers and barriers to implementation of AW&RS using the SURE framework.

We highlight the following themes according to the SURE framework:

*Knowledge, skills and experience* at the patient/population/community level; the key enabler was identified as adequate and regular training of participants on the AW&RS system, its processes, and early warning signs, as reported in five studies^32,43,47,81,91^. Conversely, the main barrier was poor health-seeking behaviour among participants, as reported in 3 studies^29,92,93^.

*Knowledge, skills and experience* at the providers of care level; 14 studies reported training on data collection, management, and analysis at the health centre as a key enabler^13,25,31,41,42,46,58,61,71,77,91,93–95^. In contrast, a low disease awareness and suspicion index, especially for NTDs, was a notable barrier reported in four studies^28,29,54,96^. At the stakeholders’ level; two studies reported the involvement of communities and health workers in surveillance processes as the enablers^97,98^.

Looking at the health system level, seven studies reported improved healthcare infrastructure, facilities and supplies for implementing the AW&RS as an enabler^26,70,94,99–102^. *Financial resources* captured several barriers, including inadequate financial resources for procurement and staff payments, reported in 21 studies^13,14,27,31,32,35,41–43,48,60,61,73,76,96,103–108^, and increased costs of modern, more acceptable, and user-friendly systems and equipment, reported in 13 studies^13,26,27,31,32,35,42,60,76,96,103–105^. *Human resources* were reported in multiple studies, with a sufficient number of skilled and trained healthcare personnel reported as enablers in 13 studies^34,35,41,65,70,73,93,94,97,98,109–111^ and the inverse reported as a barrier in nine studies^14,33,46,48,60,99,112–114^.

At the social and political level, *political stability* was highlighted as a barrier through political instability and protracted conflict hindering execution of the systems, as captured in four studies^26,62,107,108^. A notable enabler which serves as a policy and political milestone is the existence of Legislation or regulations such as the Rabies Act (Cap 365), the Animal Diseases Act (Cap 364), the Meat Control Act (Cap 356), and the Food, Drugs & Chemical Substances Act (Cap 254) ^107^. These pieces of legislation provided veterinary officers and the Ministry of Health the authority to act in a coordinated manner within differing parts of the system ^107^.

## DISCUSSION

This review has mapped out the available evidence on the existing AW&RS; the impact of climate on health and existing CSIDs; effects of the current AW&RS on ID risk and health outcomes and enablers and barriers to detecting infectious and CSIDs in Kenya. Our study observed that the most common AW&RS were IDSR and cohort surveillance systems, mainly focusing on disease burden and outbreak surveillance. Our results also indicate outbreaks of cholera and malaria were reported with increased rainfall in different regions. Data completeness, the reported proportion of death count, and timeliness of the surveillance data collected were notable performance indicators reported. The most reported enabler was adequate training of the healthcare providers, whereas inadequate financial resources were the most reported barrier.

Our review identified diverse coverage of AW&RS in the country. Most were IDSR and cohort surveillance systems focusing on human disease burden/outbreak surveillance, mainly using automated alerts over manual alerts. This could be attributed to Kenya adopting the One Health approach in 2006, which called for the establishment of coordinative frameworks that align with the global recommendation to ensure preparedness efforts to contain zoonotic and CSIDs^4^. Looking at alert mechanisms, the Global Disaster Preparedness Centre also acknowledged the use of automatic technological alerts such as Geographic Information Systems, SMS and Indicator-based Situation Reports over traditional manual alerts, citing timely detection for early response^115^.

The systems included in our review mainly surveyed malaria, RVF, HIV/AIDS and COVID-19 in Nairobi, Kilifi, Siaya, Mombasa and Kwale, sourcing data from hospitals or both hospitals and communities. This could be due to the high HIV/AIDS and malaria disease burden in these counties and its attraction for bilateral and multilateral donor agencies funding disease control programs and research in high morbidity/mortality counties, with most programs being attached to health facilities or vulnerable communities^2,116^. The recent COVID-19 pandemic, characterised by a high morbidity rate, called for increased research and surveillance to understand and predict the disease and its patterns within the country^73^. We could attribute the increased surveillance of RVF to its previous catastrophic outbreak effects in the country in the years 1996-1997 and 2006-2007, both causing over 200 reported human and animal deaths and over 25,000 infections ^117–119^. Therefore, it seems that the burden of endemic or emerging diseases is a key driver of the types, scope, and extent of AW&RS in Kenya.

In Kenya, June to August have been cool and dry seasons, followed by short rains in October-December. The hot and dry season is often experienced in January and February. Our study highlighted that CSIDs such as malaria, RVF, and cholera increase with increased precipitation and vegetative index. This is similarly reported by the Intergovernmental Panel on Climate Change, which highlights that climate change is predicted to affect human health by causing changes in the abundance and distribution of disease vectors and pathogens^120^. A systematic review looking at the impact of mosquito-borne diseases in Africa included 21 studies and reported a significant relationship between climatic changes and mosquito-borne diseases, predicting an increase in the prevalence of malaria, RVF, Chikungunya and dengue as a consequence of increased temperature and precipitation ^121^. With this possible association, it is essential to highlight that climate change is set to intensify in the East African region, including Kenya; mean annual temperatures are projected to be 2.1°C warmer than the 1994–2005 average, with heavy precipitation projected to increase at 2°C and above ^122^. However, drought frequency and intensity are predicted to decrease or remain unchanged in the region^122^.

A noteworthy key performance indicator and extent of success reported in this study were disease burden (reported proportion of death count) and reduced disease prevalence, respectively. A review reporting on AW&RS of ID found similar results, reinforcing the importance of disease or vector burden and distribution as a performance indicator, specifying the relevance of a high sensitivity to give a high confidence in the ability of disease identification, as well as identifying increasing or decreasing trends^123^.

The extent of success of the AW&RS was assessed by aspects such as its acceptability at the different levels of healthcare, which influenced the completeness and timeliness of reporting. The completeness of data on infectious or CSIDs is vital in accurate disease/vector predictions and timely responses, as iterated by a global scoping review including 37 studies by Hussein-Alkhateeb and colleagues^124^.

Our findings indicated that integrating facility-based screening and community-based outreaches provides a comprehensive approach to addressing barriers and reaching underserved populations. These findings resonate with those of Namoe and colleagues, who conclude that AW&RS have helped in livelihood resilience through the aversion of outbreaks and adaptive capacity in the arid and semi-arid areas of Turkana, which comprises underserved populations^126^.

Additionally, continuous surveillance, crucial for early detection, could be maintained using virtual remote supervision, thus enabling timely public health action. This could be impactful in the surveillance of CSIDs where modern advances in airborne remote sensing allow early detection of climate-related pathogens. Also, this can be impactful where satellite data are most commonly used to study geographically widespread human diseases^127^. Opportunities are available for combining remote sensing with animal data to support decision-making by providing crucial interlinkages in the zoonotic disease cycle^127^.

The importance of training healthcare providers on data collection, management, and analysis was reported as a key enabler for the successful implementation of AW&RS. Regular and clear stepwise training is necessary for healthcare providers since data collection for early detection and reporting is the cornerstone for effective AW&RS^128^. The importance of this is echoed by the International Federation of Red Cross and Red Crescent societies that developed a community AW&RS training tool kit to train project and partner staff members who would be responsible for running, supporting or evaluating AW&RS efforts in at-risk communities^129^. This is further backed by efforts by the European Commission to offer grants for the training of health professionals in European Union member states in ID emergencies such as anthrax^128^.

Inadequate financial resources for procurement and staff payments significantly hindered the implementation of AW&RS. A study looking at the challenges faced by AW&RS in Iraq also identified a scarcity of financial resources, ultimately affecting the purchasing of equipment and payment of skilled staff^130^. This is despite the World Bank’s estimation that investing one billion dollars in AW&RS could avoid losses of 35 billion dollars through lost lives, livelihoods and healthcare system expenditures^131^. With this in mind, a significant effort must be made to enhance investment through bilateral and multilateral funds providers and public-private sector partnerships and simplify access to financial aid for AW&RS ^132^. None of the included studies reported on the costs of the enablers and barriers to successfully implementing the AW&RS, despite having 97 studies reporting on this objective. This leaves a knowledge gap that needs to be filled.

### Implications for practice, policy and future research

This review emphasises the vital importance of knowledge enhancement for both the public and healthcare practitioners to improve AW&RS system effectiveness. Two key areas require attention: community education and sensitisation initiatives to inform the public about surveillance activities and their role, and continuous medical education (CMEs) for healthcare practitioners focusing on priority diseases.

This review underscores the links between animal, human, and environmental diseases. This emphasises the need to advocate for policies that incorporate the One Health Approach in the AW&RS through multidisciplinary integration. Integrated One Health Data management can assist predict zoonotic patterns and enable early response.

A significant knowledge gap exists in quantifying the costs associated with enablers and barriers to implementing the AW&RS. To address this, research is need in three key areas: mapping the political economy of these systems, predicting the cost implications of various factors, and estimating future expenditures. This comprehensive analysis provides crucial insights into the economic dynamics that shape these systems and facilitates evidence informed decision-making. Understanding these factors could potentially improve the efficiency and effectiveness of AW&RS.

### Strengths and limitations

The strengths of this review include a comprehensive literature search, aided by an information specialist (VL), encompassing research databases and grey literature from websites and university repositories without date restrictions. We also conducted study screening in duplicate, discussing among reviewers in case of discrepancies. These findings can be nationally generalised since incorporate articles from most counties. We identified enablers and barriers to the implementation of AW&RS and reported findings following the PRISMA-ScR guidance.

A critical limitation is the possible reviewer bias arising from conducting data extraction by one reviewer per study due to time constraints. However, we averted this by consulting with the senior reviewer (MM) about any challenges encountered. Although considered an optional step, we could not complement our review findings with follow-up discussions from the relevant stakeholders^15^.

### Conclusion

This review highlights a widespread distribution of AW&RS for infectious and CSIDs in Kenya, with the most common systems being IDSR and cohort surveillance systems, largely focusing on disease burden and outbreak surveillance. Although grey literature reported the influence of climatic changes on climate-sensitive vectors and pathogens, analytical studies are required to determine associations and draw scientific conclusions for AW&RS.

Knowledge enhancement at the community and healthcare provider level, as well as adequate funding and human resources, can ease the feasibility and effectiveness of the AW&RS. This is the most recent comprehensive review of existing AW&RS in Kenya, and it draws valuable lessons that can inform the scale-up of AW&RS in Kenya. However, research is needed to fill the knowledge gap and identify the costs of the enablers and barriers to their success.

## Ethical Considerations

This scoping review is nested in a larger study that sought ethical approval at KEMRI/SERU; approval number 5094. However, the review itself did not seek ethical considerations as it used publicly available literature.

## Author Contributions

Study conceptualisation: DO, PN, EO, KN, MM, IN. Drafting of the initial manuscript: LW.

Intellectual input on subsequent manuscript versions: LW, JO, MN, PK, DO, PN, EO, KN, MM, IN. Approval of the final draft of the manuscript: LW, JO, MN, PK, DO, PN, EO, KN, MM, IN.

## Supporting information

Supplemental file 2_Annexes

## Data Availability

The supplementary files provide additional data that support our findings. The corresponding author will provide further details upon request via email.

https://osf.io/98qsp

## Acknowledgements

We acknowledge the efforts of the information specialist, Vittoria Lutje, in conducting the database search.

