## Supplemental file 1_Tables for "Advance Warning and Response Systems in Kenya: A Scoping Review"

### Supplementary Document 1

#### TABLES

**Table 2: Distribution of Diseases in the Included Studies**

Rows typed in bold present climate-sensitive infectious diseases.

| Disease | Number of studies |
|---|---|
| **Malaria** | **35** |
| **Rift Valley Fever** | **18** |
| HIV/AIDS | 17 |
| COVID 19 | 13 |
| Polio | 8 |
| Influenza | 8 |
| **Cholera** | **8** |
| **Anthrax** | **7** |
| Human Enteric infections | 6 |
| Rabies | 6 |
| Shigellosis | 6 |
| IDSR priority diseases | 5 |
| Acute Flaccid paralysis | 5 |
| Zoonotic diseases | 5 |
| **Avian Influenza** | **5** |
| Tuberculosis | 5 |
| Pneumonia | 5 |
| **Brucellosis** | **4** |
| Acute respiratory infections | 4 |
| Measles | 4 |
| **Leishmaniasis** | **3** |
| Lymphatic Filariasis | 3 |
| **Dengue fever** | **3** |
| **Schistosomiasis** | **3** |
| Sexually-transmitted diseases | 3 |
| **Trypanosomiasis** | **3** |
| **Typhoid fever** | **3** |

| Disease | Number of studies |
|---|---|
| Q-fever | 2 |
| **Chikungunya** | **2** |
| Respiratory Distress syndrome | 2 |
| Respiratory Syncytial virus | 2 |
| Acute Febrile illness | 2 |
| **Rotavirus-diarrhoea** | **2** |
| **Severe Acute Respiratory illness** | **2** |
| **Ebola** | **2** |
| **Meningococcal meningitis** | **2** |
| Human Rhinovirus infection | 2 |
| Neglected tropical diseases | 2 |
| Hepatitis A | 2 |
| Yellow fever | 2 |
| Mumps | 1 |
| Neonatal tetanus | 1 |
| New Castle disease | 1 |
| O' Nyong Nyong Viruses | 1 |
| Orthobunyavirus disease | 1 |
| Pertussis | 1 |
| Plague | 1 |
| Amoebiasis | 1 |
| Arbovirus infection | 1 |
| Bronchitis | 1 |
| Crimean-Congo Haemorrhagic fever | 1 |
| Cysticercosis | 1 |
| Hepatitis E virus infection | 1 |
| Swine Flu | 1 |
| Marburg virus | 1 |
| Tetanus | 1 |
| Toxoplasmosis | 1 |

| Disease | Number of studies |
|---|---|
| Trachoma | 1 |
| Vaccine-preventable diseases | 1 |
| Viral haemorrhagic fever | 1 |
| Lassa fever | 1 |
| Taeniasis | 1 |
| Respiratory Viral Healthcare-Associated Infections (RHAI) | 1 |

**Table 3: Key Features of Advance warning and response Systems**

| Author, year | AW&R system name | Objective | Description | Diseases(s) [1] | Implementation level | Implementation dates of the AW&R system | AW&R System type | Purpose and scope | Alert mechanisms | Data sources |
|---|---|---|---|---|---|---|---|---|---|---|
| **Adazu 2005**[1] | Health and Demographic Surveillance System | To measure the burden of infectious diseases and evaluate public health interventions | Key actors include the community, study staff, healthcare workers, and CDC/KEMRI, operating different sections of surveillance under the | **Malaria**; HIV/AIDS | County; Healthcare facility; Community | Since 2002 | Surveillance for human diseases | Disease burden & outbreak surveillance | Not specified | Community sources; Hospital sources; Climate sources; Laboratory sources |

[1] Diseases in bold were classified as climate sensitive.

| Author, year | AW&R system name | Objective | Description | Diseases(s) [1] | Implementation level | Implementation dates of the AW&R system | AW&R System type | Purpose and scope | Alert mechanisms | Data sources |
|---|---|---|---|---|---|---|---|---|---|---|
| | | | Health and Demographic Surveillance System (HDSS) | | | | | | | |
| **Anyamba 2010**[2] | Rift Valley Fever surveillance | To conduct risk mapping and prediction assessments for Rift Valley Fever (RVF) | RVF risk mapping and prediction assessment | **Rift Valley Fever** | International | Sept 2006 to May 2007 | Surveillance for human diseases; Animal diseases and disease syndromes surveillance | Disease burden & outbreak surveillance | Not specified | Disease modelling |
| **Arale 2019**[3] | Acute flaccid paralysis surveillance | To enhance collaboration between health and administrative authorities in border regions through the | In Kenya, the government-led committees consisted of representatives from the five counties and sub | Poliomyelitis | Multi-county | 2015 to 2018 | Surveillance for human diseases | Disease burden & outbreak surveillance | Internal reports | Community sources; Hospital sources |

| Author, year | AW&R system name | Objective | Description | Diseases(s) [1] | Implementation level | Implementation dates of the AW&R system | AW&R System type | Purpose and scope | Alert mechanisms | Data sources |
|---|---|---|---|---|---|---|---|---|---|---|
| | | following initiatives: improving the sensitivity of acute flaccid paralysis (AFP) surveillance, increasing coverage of supplemental immunization activities (SIAs), and improving access to quality routine immunization services in the Horn of Africa (HOA) | counties-director for health, the disease surveillance coordinator, the Expanded Program on Immunization (EPI) coordinator, the health records and information officer, and the community health strategy focal person. | | | | | | | |

| Author, year | AW&R system name | Objective | Description | Diseases(s) [1] | Implementation level | Implementation dates of the AW&R system | AW&R System type | Purpose and scope | Alert mechanisms | Data sources |
|---|---|---|---|---|---|---|---|---|---|---|
| | | border regions. | | | | | | | | |
| **Bello 2021**[4] | Integrated supportive supervision | To conduct vaccine preventable disease surveillance | Not specified | Vaccine-preventable Diseases | National | Jan 2019 to August 2020 | Surveillance for human diseases | Disease burden & outbreak surveillance | Automated alerts | Hospital sources |
| **Bigogo 2018** [5] | Population Based Infectious Disease Surveillance | Initially used to characterize four disease syndromes and jaundice then later on used for Tb | Integrated into an existing surveillance system. | Tuberculosis; HIV/AIDS | Healthcare facility; Community | Home screening: Lwak, October to December 2011 and Kibera January to March 2012. Facility to based screening; Lwak, May 2012 to April 2014 and Kibera April 2012 to March 2014 | Surveillance for human diseases | Disease burden & outbreak surveillance | Not specified | Community sources; Hospital sources |

| Author, year | AW&R system name | Objective | Description | Diseases(s) [1] | Implementation level | Implementation dates of the AW&R system | AW&R System type | Purpose and scope | Alert mechanisms | Data sources |
|---|---|---|---|---|---|---|---|---|---|---|
| **Bisanzio 2015**[6] | Surveillance of febrile cases | To identify febrile cases | Not specified | **Malaria** | County | 2012 to 2015 | Surveillance for human diseases; Climate surveillance | Disease burden & outbreak surveillance; Environmental monitoring | Not specified | Community sources; Hospital sources; Environment sources; Climate sources; Animal sources; Remote sensing data |
| **Bolu 2007**[7] | Antenatal care sentinel surveillance | To document HIV/AIDS prevalence and monitor trends among pregnant women | Integrated with PMTCT (Prevention of Mother-to-Child Transmission) services. | HIV/AIDS | County | Since 2005 | Surveillance for human diseases | Disease burden & outbreak surveillance | Not specified | Hospital sources |
| **Boru 2023**[8] | Integrated Disease Surveillance and Response | To ensure the reporting of priority diseases, | Not specified | **Cholera** | National; County | January 1, 2015, to Dec 31, 2020 | Surveillance for human diseases | Disease burden & outbreak surveillance | Not specified | Hospital sources |

| Author, year | AW&R system name | Objective | Description | Diseases(s) 1 | Implementation level | Implementation dates of the AW&R system | AW&R System type | Purpose and scope | Alert mechanisms | Data sources |
|---|---|---|---|---|---|---|---|---|---|---|
| | | conditions, and events to the national level in accordance with International Health Regulations (IHR) requirements | | | | | | | | |
| **Breiman 2014**[9] | Population Based Infectious Disease Surveillance | The surveillance platform has provided opportunities to assess the burden of a variety of syndromes and their aetiology | Integrated with the Kenya Medical Research Institute (KEMRI) and the Centres for Disease Control and Prevention (CDC). | **Rotavirus Diarrhoeal Disease** | Healthcare facility; Community | January 1, 2007, and December 31, 2010 | Surveillance for human diseases | Disease burden & outbreak surveillance | Not specified | Community sources; Hospital sources |
| **Byrne 2020**[126] | Community Epidemic | To build community | Implemented in eight | Measles; Poliomyeliti | International | Since 2018 | Surveillance for | Disease burden & | Automated alerts | Community sources |

| Author, year | AW&R system name | Objective | Description | Diseases(s) [1] | Implementation level | Implementation dates of the AW&R system | AW&R System type | Purpose and scope | Alert mechanisms | Data sources |
|---|---|---|---|---|---|---|---|---|---|---|
| | and Pandemic Preparedness Programme | -level epidemic and pandemic preparedness with a One Health approach encompassing both human and animal disease | countries using mobile phone technology, allowing volunteers to submit reports via short message service (SMS), applications, or electronic forms for real-time automated data processing. | s; **Rift valley fever; Cholera; Anthrax;** Rabies | | | human diseases; Animal diseases and disease syndromes surveillance | outbreak surveillance | | |
| **Campbell 2019**[10] | Rift Valley Fever surveillance | To predict the abundances of adult *Aedes mcintoshi* mosquitoes | The system uses a combination of georeferenced mosquito | **Rift Valley Fever** | Not specified | 2007 to 2012 | Animal diseases and disease syndromes surveillance; Environme | Environmental monitoring; Improve forecasting | Not specified | Climate sources; Animal sources; Remote sensing data; |

| Author, year | AW&R system name | Objective | Description | Diseases(s) [1] | Implementation level | Implementation dates of the AW&R system | AW&R System type | Purpose and scope | Alert mechanisms | Data sources |
|---|---|---|---|---|---|---|---|---|---|---|
| | | | abundance data, remotely sensed environmental variables, and a predictive modelling approach. | | | | ntal surveillance | | | Disease modeling; Secondary sources |
| **Cheluget 2004**[11] | HIV/AIDS sentinel surveillance | To monitor the level of HIV infection | Surveillance was conducted annually for a period of up to 3 months during which patients with STDs (first visit of the symptom episode) were consecutiv | Sexually Transmitted Diseases | National | 1990 to 2001 | Surveillance for human diseases | Disease burden & outbreak surveillance | Not specified | Hospital sources |

| Author, year | AW&R system name | Objective | Description | Diseases(s) [1] | Implementation level | Implementation dates of the AW&R system | AW&R System type | Purpose and scope | Alert mechanisms | Data sources |
|---|---|---|---|---|---|---|---|---|---|---|
| | | | ely recruited from selected sentinel sites. | | | | | | | |
| **Curran 2018**[12] | Cholera outbreak surveillance | To assess cholera outbreak preparedness | Key actors include Community Health Extension Workers (CHEWs) and Community Health Workers (CHWs). | **Cholera** | Multi-county | June to July 2015 | Surveillance for human diseases | Disaster preparedness | Manual notifications | Hospital sources |
| **Ear 2013**[139] | Global Emerging Infections Surveillance and Response System | To improve surveillance and outbreak response capacities in most sub-Saharan | Cooperative arrangement with KEMRI (Kenya Medical Research Institute). | Influenza; **Ebola Viral Hemorrhagic Fever**; Acute Febrile Illnesses; **Leishmaniasis; Enteric Diseases;** | Multi-county | Since 2012 | Surveillance for human diseases | Disease burden & outbreak surveillance | Not specified | Not specified |

| Author, year | AW&R system name | Objective | Description | Diseases(s) 1 | Implementation level | Implementation dates of the AW&R system | AW&R System type | Purpose and scope | Alert mechanisms | Data sources |
|---|---|---|---|---|---|---|---|---|---|---|
| | | African countries | | Sexually Transmitted Diseases; Rodent-Borne Viral Diseases | | | | | | |
| **Fagnant 2018**[13] | Bag-mediated filtration environmental surveillance | To improve the usability and sensitivity of the previously developed bag-mediated filtration system (BMFS) | This system allows for in-field filtration of samples without requiring power, is compact enough to fit in a backpack, and is easy to use. | Not Specified | County | Not specified | Environmental surveillance | Environmental monitoring | Manual notifications | Environment sources |
| **Falzon 2019**[14] | Integrated surveillance system for zoonotic diseases | To inform the public sector integration of surveillance across animal and | Integrated surveillance system for zoonotic diseases in western Kenya by | Q-Fever; **Meningitis;** Bovine Tuberculosis; **Rift Valley Fever;** Cysticercosi | Multi-county | July to August 2016 and September to December 2016 | Surveillance for human diseases; Animal diseases and disease syndromes | Disease burden & outbreak surveillance | Not specified | Hospital sources; Animal sources |

| Author, year | AW&R system name | Objective | Description | Diseases(s) [1] | Implementation level | Implementation dates of the AW&R system | AW&R System type | Purpose and scope | Alert mechanisms | Data sources |
|---|---|---|---|---|---|---|---|---|---|---|
| | | human sectors, providing key data on efficacy, cost-effectiveness, coverage and effort | integrating staff and structures for sample collection and analysis across animal and human sectors and fostering engagement. | s; **Anthrax; Brucellosis; Trypanosomiasis** | | | surveillance | | | |
| **Geteri 2024**[15] | Integrated Disease Surveillance and Response | To ensure the reporting of priority diseases, conditions, and events to the national level in accordance with International Health | Surveillance data held by the Disease Surveillance and Response Unit (DSRU) of the Ministry of Health, Kenya, | **Anthrax; Chikungunya; Cholera; Dengue Fever; Diarrheal Diseases; Leishmaniasis (Kala-azar); Malaria; Rift valley fever;** | National | 2007 to 2022 | Surveillance for human diseases | Disease burden & outbreak surveillance; Disaster preparedness | Internal reports | Not specified |

| Author, year | AW&R system name | Objective | Description | Diseases(s) [1] | Implementation level | Implementation dates of the AW&R system | AW&R System type | Purpose and scope | Alert mechanisms | Data sources |
|---|---|---|---|---|---|---|---|---|---|---|
| | | Regulations (IHR) requirements | 2007 to 2022. | **Schistosomiasis (Bilharzia); Yellow Fever;** Acute flaccid paralysis; aflatoxicosis; COVID-19; Influenza A; Measles; Mumps; Pertussis; Q-fever; SARI; Viral Hepatitis | | | | | | |
| **Feikin 2010**[16] | Population Based Infectious Disease Surveillance | To define disease burden, describe epidemiologic patterns of disease and evaluate the health | Disease is characterized in the clinic and through biweekly home visits by field workers, who | Infectious Diseases | Multi-county | Since 2005 | Surveillance for human diseases | Disease burden & outbreak surveillance | Automated alerts | Community sources |

| Author, year | AW&R system name | Objective | Description | Diseases(s) [1] | Implementation level | Implementation dates of the AW&R system | AW&R System type | Purpose and scope | Alert mechanisms | Data sources |
|---|---|---|---|---|---|---|---|---|---|---|
| | | impact of interventions over time in these populations | inquire about recent symptoms. | | | | | | | |
| **Feikin 2011**[17] | Population Based Infectious Disease Surveillance | Not specified | Not specified | Acute Respiratory Infection | County | Since late 2005 | Surveillance for human diseases | Disease burden & outbreak surveillance | Automated alerts | Community sources; Hospital sources |
| **Fujii 2014**[18] | Health and Demographic Surveillance System | Not specified | Not specified | Amoebiasis; **Leishmaniasis;** Toxoplasmosis; Lymphatic Filariasis; HIV/AIDS; **Cholera** | County | July 2011 to August 2011 | Surveillance for human diseases | Disease burden & outbreak surveillance | Not specified | Community sources; Hospital sources |
| **Furukawa 2016**[19] | Population Based Infectious Disease | To track changes and enhance early | PBIDS has been operating in Kibera since 2005, | Hepatitis E Virus infection | County | Since 2005 | Surveillance for human diseases | Disease burden & outbreak surveillance | Manual notifications | Community sources |

| Author, year | AW&R system name | Objective | Description | Diseases(s) 1 | Implementation level | Implementation dates of the AW&R system | AW&R System type | Purpose and scope | Alert mechanisms | Data sources |
|---|---|---|---|---|---|---|---|---|---|---|
| | Surveillance | warning detection of infectious diseases | serving an estimated population of 25,000 to 29,000 individuals within a 0.37 $km^2$ area. | | | | | | | |
| **Gachohi 2012**[20] | Rift Valley Fever surveillance | Not specified | Not specified | **Rift Valley Fever** | National; County; Sub-County | Not specified | Surveillance for human diseases; Animal diseases and disease syndromes surveillance | Disease burden & outbreak surveillance | Internal reports | Community sources; Media scanning; Laboratory sources |
| **Gardner 2018**[127] | Not specified | To perform surveillance for acute flaccid paralysis (AFP) among children | Not specified | Acute Flaccid Paralysis | National | 2011 to 2017 | Surveillance for human diseases | Disease burden & outbreak surveillance | Not specified | Community sources; Hospital sources |

| Author, year | AW&R system name | Objective | Description | Diseases(s) [1] | Implementation level | Implementation dates of the AW&R system | AW&R System type | Purpose and scope | Alert mechanisms | Data sources |
|---|---|---|---|---|---|---|---|---|---|---|
| | | aged <15 years | | | | | | | | |
| **Geiger 2024**[128] | Global Polio Laboratory Network | To support the global polio eradication initiative by 1) detecting poliovirus circulation, 2) supporting outbreak response efforts, 3) monitoring vaccine coverage and efficacy, and 4) providing data for strategic decision-making | The Global Polio Laboratory Network (GPLN) is an extensive network of laboratories that play a crucial role in laboratory testing, genetic sequencing, and data sharing related to polio. | Poliomyelitis | National | January 1, 2022, to December 31, 2023 | Surveillance for human diseases | Disease burden & outbreak surveillance | Automated alerts | Hospital sources; Environment sources; Laboratory sources |

| Author, year | AW&R system name | Objective | Description | Diseases(s) 1 | Implementation level | Implementation dates of the AW&R system | AW&R System type | Purpose and scope | Alert mechanisms | Data sources |
|---|---|---|---|---|---|---|---|---|---|---|
| **Gerken 2022**[21] | Rift Valley Fever surveillance | Not specified | The system is implied to monitor and assess regions for potential RVFV outbreaks, signalling risk areas to guide preventive and response measures. | **Rift Valley Fever** | County | Since November 2021 | Animal diseases and disease syndromes surveillance | Disease burden & outbreak surveillance | Not specified | Animal sources |
| **Gikungu 2016**[22] | Rift Valley Fever surveillance | A dynamic risk model based on historical RVF outbreaks and climate data to guide veterinary and public health policies on | The Kenya Meteorological Department (KMD), International Research Institute for Climate and Society (IRI), and the Centre for Disaster | **Rift Valley Fever** | Multi-county | Not specified | Climate surveillance | Disease burden & outbreak surveillance; Improve forecasting | Not specified | Community sources; Climate sources; Remote sensing data; Disease modelling |

| Author, year | AW&R system name | Objective | Description | Diseases(s) [1] | Implementation level | Implementation dates of the AW&R system | AW&R System type | Purpose and scope | Alert mechanisms | Data sources |
|---|---|---|---|---|---|---|---|---|---|---|
| | | prevention and control of RVF outbreaks | Management and Humanitarian Assistance (CDMHA) at Masinde Muliro University of Science and Technology are collaborating with farmers in Garissa, Murang'a, and Kwale counties to develop a model that complements existing early warning systems. | | | | | | | |

| Author, year | AW&R system name | Objective | Description | Diseases(s) [1] | Implementation level | Implementation dates of the AW&R system | AW&R System type | Purpose and scope | Alert mechanisms | Data sources |
|---|---|---|---|---|---|---|---|---|---|---|
| **Githinji 2014**[23] | Mobile short Message Service for Life system | To utilize a web-based reporting tool that captures data sent via SMS by registered health workers, making it accessible in real-time to designated members of the District Health Management Team (DHMT) and national officials through the internet | As one approach towards this realization, the system piloted a project SMS for Life, using mobile phone text messages to transmit data directly from health facilities. | **Malaria** | County; Sub-County | August 2011 to March 2012 | Surveillance for human diseases | Disease burden & outbreak surveillance | Automated alerts | Hospital sources |

| Author, year | AW&R system name | Objective | Description | Diseases(s) 1 | Implementation level | Implementation dates of the AW&R system | AW&R System type | Purpose and scope | Alert mechanisms | Data sources |
|---|---|---|---|---|---|---|---|---|---|---|
| **Githure 2022**[24] | Malaria surveillance | To provide a platform for early detection and control of malaria | Not specified | **Malaria** | County | Not specified | Surveillance for human diseases | Disease burden & outbreak surveillance | Automated alerts | Hospital sources |
| **Hammitt 2019**[25] | Health and Demographic Surveillance System (Kilifi) | To track the incidence of invasive pneumococcal disease (IPD) in children, particularly focusing on the serotypes included in the PCV10 vaccine | Fieldworkers visited households approximately every four months to record demographic events and monitor changes within the population, collecting health data with a focus on hospital visits, especially | HIV/AIDS; Pneumonia | County | Since 2000 | Surveillance for human diseases | Disease burden & outbreak surveillance | Not specified | Hospital sources |

| **Author, year** | **AW&R system name** | **Objective** | **Description** | **Diseases(s)** [1] | **Implementation level** | **Implementation dates of the AW&R system** | **AW&R System type** | **Purpose and scope** | **Alert mechanisms** | **Data sources** |
|---|---|---|---|---|---|---|---|---|---|---|
| | | | at Kilifi County Hospital, which serves the entire Kilifi Health and Demographic Surveillance System (KHDSS) population. | | | | | | | |
| **Harris 2020**[155] | Malaria surveillance | To provide a platform for early detection and control of malaria | The malaria surveillance system detects early warning signals as critical points are approached, with increases in various summary statistics | **Malaria** | County | December 1981 to April 1993 | Surveillance for human diseases | Disease burden & outbreak surveillance | Not specified | Hospital sources |

| Author, year | AW&R system name | Objective | Description | Diseases(s) 1 | Implementation level | Implementation dates of the AW&R system | AW&R System type | Purpose and scope | Alert mechanisms | Data sources |
|---|---|---|---|---|---|---|---|---|---|---|
| | | | that precede a rise in cases at the onset of the supercritical period, triggering alerts in the moving epidemic method. | | | | | | | |
| **Hassan 2020**[27] | Community based syndromic surveillance | To monitor the occurrence of RVF-associated syndromes in livestock | Key actors include the County Departments of Health and Livestock, the community, KEMRI, the National Public Health Laboratorie | **Rift Valley Fever** | Multi-county | May to June 2018 | Animal diseases and disease syndromes surveillance | Disease burden & outbreak surveillance | Internal reports | Community sources; Hospital sources; Laboratory sources |

| Author, year | AW&R system name | Objective | Description | Diseases(s) [1] | Implementation level | Implementation dates of the AW&R system | AW&R System type | Purpose and scope | Alert mechanisms | Data sources |
|---|---|---|---|---|---|---|---|---|---|---|
| | | | s, the Central Veterinary Laboratory virology staff, and the Kenya Directorate of Veterinary Services. The system was previously utilized in 2015-2016 across 22 high-risk counties for Rift Valley Fever (RVF). | | | | | | | |
| **Hay 1998**[28] | Malaria surveillance | To predict the seasonality of malaria in Kenya | Predictions were made by analysing relationshi | **Malaria** | County | 1990 to 1994 | Surveillance for human diseases; Climate | Disease burden & outbreak surveillanc | Automated alerts | Climate sources; Remote sensing data |

| Author, year | AW&R system name | Objective | Description | Diseases(s) [1] | Implementation level | Implementation dates of the AW&R system | AW&R System type | Purpose and scope | Alert mechanisms | Data sources |
|---|---|---|---|---|---|---|---|---|---|---|
| | | using remotely sensed images from satellite sensors | ps between long-term data on paediatric severe malaria admissions and concurrently collected data from the Advanced Very High Resolution Radiometer (AVHRR) on NOAA polar-orbiting meteorological satellites and the High-Resolution Radiometer (HRR) on | | | | surveillance | e; Improve forecasting | | |

| Author, year | AW&R system name | Objective | Description | Diseases(s) 1 | Implementation level | Implementation dates of the AW&R system | AW&R System type | Purpose and scope | Alert mechanisms | Data sources |
|---|---|---|---|---|---|---|---|---|---|---|
| | | | EUMETSAT geostationary Meteosat satellites. | | | | | | | |
| **Hay 2000**[29] | Not specified | Not specified | Not specified | **Malaria** | International | 1996 to 1988 | Surveillance for human diseases; Climate surveillance | Disease burden & outbreak surveillance; Disaster preparedness | Manual notifications | Hospital sources |
| **Hendriksen 2019**[30] | Population Based Infectious Disease Surveillance | Not specified | Household morbidity and healthcare usage data were collected biweekly through home visits, while blood and stool samples were collected at | Infectious Diseases | County | June 16 to August 26, 2014 | Environmental surveillance | Environmental monitoring | Internal reports | Environment sources |

| Author, year | AW&R system name | Objective | Description | Diseases(s) [1] | Implementation level | Implementation dates of the AW&R system | AW&R System type | Purpose and scope | Alert mechanisms | Data sources |
|---|---|---|---|---|---|---|---|---|---|---|
| | | | the clinic and transported to the laboratory within two hours; sewage samples were collected every Monday and Wednesday for immediate processing. | | | | | | | |
| **Homan 2016**[129] | Health and Demographic Surveillance System (Rusinga) | To measure the effectiveness of the trial on clinical malaria incidence, and to monitor | The HDSS operates by house-to-house interviews. Clinical malaria is recorded during HDSS | **Malaria** | County | 2012 to 2014 | Surveillance for human diseases | Disease burden & outbreak surveillance | Automated alerts | Community sources |

| Author, year | AW&R system name | Objective | Description | Diseases(s) [1] | Implementation level | Implementation dates of the AW&R system | AW&R System type | Purpose and scope | Alert mechanisms | Data sources |
|---|---|---|---|---|---|---|---|---|---|---|
| | | demographic, environmental and malaria-related data variable | rounds, based on fever recalls and a conditional rapid diagnostic test. | | | | | | | |
| **Iacono 2018**[31] | Ecoepidemiological, compartmental, mathematical model | To simulate the population dynamics of Aedes and Culex mosquitos | It integrates an ecological, stage-structured population dynamics model for Aedes and Culex mosquito species | **Rift Valley fever** | Not applicable | Not applicable | Surveillance for health events | Disease outbreak surveillance | Not reported | Community sources; Environment sources |
| **Iuliano 2018**[32] | Influenza virus surveillance | Not specified | Not specified | Influenza | International | 1999 to 2015 | Surveillance for human diseases | Disease burden & outbreak surveillance | Not specified | Disease modelling |

| **Author, year** | **AW&R system name** | **Objective** | **Description** | **Diseases(s) 1** | **Implementation level** | **Implementation dates of the AW&R system** | **AW&R System type** | **Purpose and scope** | **Alert mechanisms** | **Data sources** |
|---|---|---|---|---|---|---|---|---|---|---|
| **Jebiwot 2018**[140] | Therapeutic efficacy studies (TES) | Surveillance of drug resistance | Tests involve treating a group of symptomatic and aparasitemic individuals with known doses of the drug and monitoring parasitological and/or clinical response. | **Malaria** | County level | 2016-2017 | Surveillance for health events | Drug resistance surveillance | Not specified | Laboratory sources |
| **Joesoef 2003**[33] | HIV/AIDS sentinel surveillance | To monitor the prevalence and trends of HIV infection among STD patients aged 15 to 49 across | Surveillance was conducted annually for a period of up to three months during which STD | HIV/AIDS | National | 1990 to 2001 | Surveillance for human diseases | Disease burden & outbreak surveillance | Manual notifications | Hospital sources |

| Author, year | AW&R system name | Objective | Description | Diseases(s) [1] | Implementation level | Implementation dates of the AW&R system | AW&R System type | Purpose and scope | Alert mechanisms | Data sources |
|---|---|---|---|---|---|---|---|---|---|---|
| | | various regions and cultural groups in the country. | patients were consecutively recruited from selected sentinel sites selected from around the country to ensure representation of different regional and cultural group. | | | | | | | |
| **Jones 2008**[34] | Malaria surveillance | To provide a platform for early detection and control of malaria | Key actors include the District Health Management Team and the | **Malaria** | Sub-county | 2001 to 2006 | Surveillance for human diseases | Disease burden & outbreak surveillance | Manual notifications | Hospital sources |

| Author, year | AW&R system name | Objective | Description | Diseases(s) 1 | Implementation level | Implementation dates of the AW&R system | AW&R System type | Purpose and scope | Alert mechanisms | Data sources |
|---|---|---|---|---|---|---|---|---|---|---|
| | | | Ministry of Health, with the system operating as a stand-alone initiative and plans for scale-up. | | | | | | | |
| **Kagendi 2023**[35] | Not specified | Not specified | Pseudonymized laboratory patient-level viral load data from NASCOP was used for the analysis. | HIV/AIDS | National | January 2015 to December 2019 | Surveillance for human diseases | Disease burden & outbreak surveillance | Automated alerts | Disease modelling |
| **Kaneko 2012**[36] | Health and Demographic Surveillance System | Not specified | Not specified | **Malaria; Schistosomiasis;** Lymphatic Filariasis; HIV/AIDS; | County | Since 2008 | Surveillance for human diseases | Disease burden & outbreak surveillance | Automated alerts | Community sources |

| **Author, year** | **AW&R system name** | **Objective** | **Description** | **Diseases(s)** [1] | **Implementation level** | **Implementation dates of the AW&R system** | **AW&R System type** | **Purpose and scope** | **Alert mechanisms** | **Data sources** |
|---|---|---|---|---|---|---|---|---|---|---|
| | | | | Tuberculosis; **Diarrheal Diseases** | | | | | | |
| **Kapesa 2018**[37] | Malaria surveillance | To utilize passive malaria surveillance in hospitals as a predictor of community transmission dynamics | Health care workers. | **Malaria** | Healthcare facility; Community | June 2015 to December 2016 | Surveillance for human diseases | Disease burden & outbreak surveillance | Not specified | Community sources; Hospital sources |
| **Kariithi 2021**[38] | Newcastle surveillance | To investigate the detectability of virulent Newcastle Disease viruses in both healthy birds and those exhibiting | Not specified | Newcastle Disease | Multi-county | 2017 to 2018 | Animal diseases and disease syndromes surveillance | Disease burden & outbreak surveillance | Not specified | Animal sources |

| Author, year | AW&R system name | Objective | Description | Diseases(s) [1] | Implementation level | Implementation dates of the AW&R system | AW&R System type | Purpose and scope | Alert mechanisms | Data sources |
|---|---|---|---|---|---|---|---|---|---|---|
| | | clinical signs consistent with Newcastle Disease across different ecogeographical and socio-cultural regions in Kenya | | | | | | | | |
| **Kasolo 2013**[39] | Integrated Disease Surveillance and Response | To detect, confirm, and respond to high-priority communicable and noncommunicable diseases | The IDSR strategy initially focused on integrating the guidance and requirements for collecting, analysing, and reporting | Infectious and non-infectious Diseases | International | Not specified | Surveillance for human diseases | Disease burden & outbreak surveillance; Disaster preparedness | Not specified | Not specified |

| Author, year | AW&R system name | Objective | Description | Diseases(s) [1] | Implementation level | Implementation dates of the AW&R system | AW&R System type | Purpose and scope | Alert mechanisms | Data sources |
|---|---|---|---|---|---|---|---|---|---|---|
| | | | data on 19 priority diseases at the district level, aiming to reduce the inefficiencies caused in many countries by parallel disease-specific surveillance program. | | | | | | | |
| **Katz 2014**[40] | National sentinel influenza surveillance | To identify circulating influenza strains, to understand the epidemiology and burden of influenza in Kenya, and to serve as | Key actors were the Kenyan Ministry of Health, with technical support from the Centers for Disease Control and | **Avian Influenza** | Multi-county | July 1, 2007, to June 30, 2013 | Surveillance for human diseases | Disease burden & outbreak surveillance | Not specified | Hospital sources |

| Author, year | AW&R system name | Objective | Description | Diseases(s) 1 | Implementation level | Implementation dates of the AW&R system | AW&R System type | Purpose and scope | Alert mechanisms | Data sources |
|---|---|---|---|---|---|---|---|---|---|---|
| | | a component of an early warning system for pandemic influenza | Prevention-Kenya | | | | | | | |
| **Keshavamurthy 2021**[41] | Not specified | Not specified | Not specified | **Rift Valley Fever; Anthrax;** Rabies; **Brucellosis; Trypanosomiasis** | National; County | Not specified | Surveillance for human diseases; Animal diseases and disease syndromes surveillance | Disease burden & outbreak surveillance | Public announcements | Media scanning |
| **Kimata 2020**[42] | Measles case-based surveillance for arboviruses | Not specified | The surveillance was based on an existing national measles/rubella case-based surveillance program. | **Chikungunya; O'-Nyong Nyong Viruses** | Multi-county | Not specified | Surveillance for human diseases | Disease burden & outbreak surveillance | Not specified | Hospital sources |

| Author, year | AW&R system name | Objective | Description | Diseases(s) 1 | Implementation level | Implementation dates of the AW&R system | AW&R System type | Purpose and scope | Alert mechanisms | Data sources |
|---|---|---|---|---|---|---|---|---|---|---|
| **Kirimi 2016**[141] | RHAI surveillance | To establish the incidence and risk factors of healthcare-associated infections | A daily line list was compiled for patients with clinical criteria for healthcare-associated infections (HAIs), and detailed information, including patient demographics, symptoms, and hospitalization details, was recorded using HAI Report Forms and tracked by research assistants | Respiratory Viral Healthcare Associated Infections (RHAI) | Multi-county level | 2009-2010 | Surveillance for health events | Disease outbreak surveillance | Manual notifications | Laboratory sources |

| Author, year | AW&R system name | Objective | Description | Diseases(s) [1] | Implementation level | Implementation dates of the AW&R system | AW&R System type | Purpose and scope | Alert mechanisms | Data sources |
|---|---|---|---|---|---|---|---|---|---|---|
| | | | until patient discharge or death. | | | | | | | |
| **Kirinyet 2016**[43] | Malaria surveillance | To provide a platform for early detection and control of malaria | The malaria surveillance and response system for the epidemic-prone districts, managed by the Division of Disease Surveillance and Response, is an important part of Kenya's 2009-2017 monitoring and | **Malaria** | County | Not specified | Surveillance for human diseases | Disease burden & outbreak surveillance; Disaster preparedness | Not specified | Community sources; Hospital sources |

| Author, year | AW&R system name | Objective | Description | Diseases(s) 1 | Implementation level | Implementation dates of the AW&R system | AW&R System type | Purpose and scope | Alert mechanisms | Data sources |
|---|---|---|---|---|---|---|---|---|---|---|
| | | | evaluation plan. | | | | | | | |
| **Kitala 2000**[44] | Rabies surveillance | To estimate the incidence of suspected and confirmed human and animal rabies cases | This one-year community-based active surveillance project involved rabies workers actively monitoring all animal bites and suspected rabies cases, categorizing suspect animals into primary and secondary cases while recording | Rabies | County | 1992 to 1993 | Surveillance for human diseases; Animal diseases and disease syndromes surveillance | Disease burden & outbreak surveillance | Manual notifications | Community sources; Hospital sources; Animal sources |

| Author, year | AW&R system name | Objective | Description | Diseases(s) [1] | Implementation level | Implementation dates of the AW&R system | AW&R System type | Purpose and scope | Alert mechanisms | Data sources |
|---|---|---|---|---|---|---|---|---|---|---|
| | | | detailed clinical signs and exposure circumstances, alongside existing passive surveillance. | | | | | | | |
| **Kyobutungi 2008**[45] | Urban Health and Demographic Surveillance System (Nairobi) | To collect detailed and reliable data on core demographic events (such as births, deaths, in-migrations, and out-migrations) and health-related outcomes | Collection of surveillance data on demographic events (birth, death, in-migration and out-migration) and updated every four months during routine DSS | Pneumonia; HIV/AIDS; Tuberculosis; **Malaria** | County | Since 2003 | Surveillance for human diseases | Disease burden & outbreak surveillance | Automated alerts | Community sources; Hospital sources |

| **Author, year** | **AW&R system name** | **Objective** | **Description** | **Diseases(s)** [1] | **Implementation level** | **Implementation dates of the AW&R system** | **AW&R System type** | **Purpose and scope** | **Alert mechanisms** | **Data sources** |
|---|---|---|---|---|---|---|---|---|---|---|
| | | | rounds where a detailed verbal autopsy questionnaire is administered to a close relative of the deceased or other credible respondent | | | | | | | |
| **Lickness 2020**[130] | Acute flaccid paralysis surveillance | To detect poliovirus circulation among children < 15 years with testing of stool specimens for wild poliovirus and | Key actors include the WHO Regional Office for Africa, the national team, the Global Immunization Division, the Centre | Poliomyelitis | International | 2018 to 2019 | Surveillance for human diseases; Environmental surveillance | Disease burden & outbreak surveillance; Environmental monitoring | Not specified | Community sources; Hospital sources; Environment sources; Laboratory sources |

| Author, year | AW&R system name | Objective | Description | Diseases(s) 1 | Implementation level | Implementation dates of the AW&R system | AW&R System type | Purpose and scope | Alert mechanisms | Data sources |
|---|---|---|---|---|---|---|---|---|---|---|
| | | vaccine-derived polioviruses in WHO-accredited laboratories and to supplement AFP with environmental surveillance, regular collection and testing of sewage to provide awareness of the extent and duration of poliovirus circulation | for Global Health at the CDC, the Polio and Picornavirus Laboratory Branch, the Division of Viral Diseases, the Geospatial Research, Analysis, and Services Program, the Agency for Toxic Substances and Disease Registry, and the Division of Emergency | | | | | | | |

| Author, year | AW&R system name | Objective | Description | Diseases(s) 1 | Implementation level | Implementation dates of the AW&R system | AW&R System type | Purpose and scope | Alert mechanisms | Data sources |
|---|---|---|---|---|---|---|---|---|---|---|
| | | | Operations at the CDC, with AFP surveillance being supplemented by environmental surveillance. | | | | | | | |
| **Lucinde 2024**[46] | SARS-CoV-2 Surveillance System | Not specified | Not specified | COVID-19 | National | Since 2020 | Surveillance for human diseases | Disease burden & outbreak surveillance | Automated alerts | Hospital sources |
| **Luka 2020**[47] | Health Demographic Surveillance System | Not specified | Not specified | Human Rhinovirus Infection | Community | May 2017 and April 2018 | Surveillance for human diseases | Disease burden & outbreak surveillance | Not specified | Community sources |
| **Maes 2017**[131] | Acute flaccid paralysis surveillance | To implement a system for collecting and testing stool | This system encompasses the collection and testing of stool specimens | Poliomyelitis | International | Not specified | Surveillance for human diseases | Disease burden & outbreak surveillance | Not specified | Community sources; Hospital sources |

| Author, year | AW&R system name | Objective | Description | Diseases(s) [1] | Implementation level | Implementation dates of the AW&R system | AW&R System type | Purpose and scope | Alert mechanisms | Data sources |
|---|---|---|---|---|---|---|---|---|---|---|
| | | specimens from individuals with acute flaccid paralysis (AFP) in WHO-accredited laboratories within the Global Polio Laboratory Network, and to utilize environmental surveillance by testing sewage samples for polioviruses to enhance AFP | from individuals with acute flaccid paralysis (AFP) in WHO-accredited laboratories within the Global Polio Laboratory Network, and it also incorporates environmental surveillance by testing sewage samples for polioviruses to enhance AFP | | | | | | | |

| Author, year | AW&R system name | Objective | Description | Diseases(s) [1] | Implementation level | Implementation dates of the AW&R system | AW&R System type | Purpose and scope | Alert mechanisms | Data sources |
|---|---|---|---|---|---|---|---|---|---|---|
| | | surveillance. | surveillance efforts. | | | | | | | |
| **Maina 2017**[48] | District Health Information System version 2 | To utilize DHIS2 as a platform for capturing, viewing, and analysing all health data, including malaria testing and positivity rates, across all levels of the health system, from reporting facilities to district and national aggregates | The DHIS2 system enables health facilities to report and visualize their monthly data on diseases, commodities, and services. | **Malaria** | National | November 2015 to October 2016 | Surveillance for human diseases | Disease burden & outbreak surveillance | Automated alerts | Hospital sources |

| Author, year | AW&R system name | Objective | Description | Diseases(s) 1 | Implementation level | Implementation dates of the AW&R system | AW&R System type | Purpose and scope | Alert mechanisms | Data sources |
|---|---|---|---|---|---|---|---|---|---|---|
| **Maraka 2020**[49] | Malaria surveillance | Not specified | Not specified | **Malaria** | Multi-county | Not specified | Surveillance for human diseases | Disease burden & outbreak surveillance | Not specified | Hospital sources |
| **Matete 2003**[50] | Integrated surveillance system for zoonotic diseases | To conduct surveillance and management of infectious diseases, including trypanosomiasis | The Integrated Surveillance and Response System combines community, clinical, epidemiological, and environmental information to provide a comprehensive view of disease trends and outbreaks. | **Trypanosomiasis** | Community | April 2001 to March 2002 | Surveillance for human diseases; Animal diseases and disease syndromes surveillance | Disease burden & outbreak surveillance | Automated alerts | Community sources; Hospital sources; Animal sources |

| Author, year | AW&R system name | Objective | Description | Diseases(s) [1] | Implementation level | Implementation dates of the AW&R system | AW&R System type | Purpose and scope | Alert mechanisms | Data sources |
|---|---|---|---|---|---|---|---|---|---|---|
| **Mathenge 2018**[142] | Geo-database | TB real-time surveillance and tracking | The spatial location of health facilities was collected by use of Handheld GPS devices | Tuberculosis | County level | Not reported | Surveillance for health events | Disease outbreak surveillance | Automated alerts (use of technology / pre-defined communication channels like SMS) | Community sources; Hospital sources |
| **Milliano 2022**[143] | Integrated Disease Surveillance and Response | To collect health data on priority diseases both communicable and non-communicable to enable prevention and control by connecting populations, Centres of health, | Constant, methodical gathering, review, and evaluation of information related to health. | IDSR Priority Diseases | County | Since 1998 | Surveillance for human diseases | Disease burden & outbreak surveillance | Automated alerts | Hospital sources |

| Author, year | AW&R system name | Objective | Description | Diseases(s) 1 | Implementation level | Implementation dates of the AW&R system | AW&R System type | Purpose and scope | Alert mechanisms | Data sources |
|---|---|---|---|---|---|---|---|---|---|---|
| | | regions, states, and nationwide levels | | | | | | | | |
| **Montana 2008**[51] | Antenatal care sentinel surveillance | To monitor HIV/AIDS prevalence in pregnancy. | Not specified. | HIV/AIDS | International | Since 2003 | Surveillance for human diseases | Disease burden & outbreak surveillance | Not specified | Community sources; Hospital sources |
| **Morobe 2019**[144] | Kilifi Health and Demographic Surveillance System (KHDSS) | NR | NR | Human Rhinovirus | County level | 2015-2016 | Surveillance for health events | Disease outbreak surveillance | Manual notifications | Laboratory sources |
| **Moturi 2014**[52] | Mobile-based Disease Surveillance Prototype System | To develop a prototype system in the form of an early-warning disease response system | The prototype enables medical notifiers in the field to report notifiable diseases by registering their SIM card, | Not Specified | County | Not specified | Surveillance for human diseases | Disease burden & outbreak surveillance | Automated alerts | Community sources, Hospital sources; Media scanning |

| Author, year | AW&R system name | Objective | Description | Diseases(s) [1] | Implementation level | Implementation dates of the AW&R system | AW&R System type | Purpose and scope | Alert mechanisms | Data sources |
|---|---|---|---|---|---|---|---|---|---|---|
| | | | entering case data, and sending messages to both district and national offices for faster responses, while also analysing data to identify outbreaks and generate reports, including charts | | | | | | | |
| **Mtetwa 2024**[53] | Wastewater based epidemiology | To conduct disease surveillance and monitor pharmaceutical | Sampled population were the wastewater treatment plant (WWTP) | Tuberculosis | International | Not specified | Environmental surveillance | Environmental monitoring | Not specified | Environment sources |

| Author, year | AW&R system name | Objective | Description | Diseases(s) 1 | Implementation level | Implementation dates of the AW&R system | AW&R System type | Purpose and scope | Alert mechanisms | Data sources |
|---|---|---|---|---|---|---|---|---|---|---|
| | | consumption | offering treatment of designated centres for TB treatments hospital sewage. | | | | | | | |
| **Munywoki 2023**[54] | Population Based Infectious Disease Surveillance | To monitor burden and aetiology of common and re-emerging acute infectious diseases and evaluate the impact of public health interventions in the two populations in Kenya | The surveillance of SARS-CoV-2 was integrated into the existing PBIDs system. | SARS-CoV-2 Infection | Multi-county; Community | March and June 2021 | Surveillance for human diseases | Disease burden & outbreak surveillance | Not specified | Community sources; Hospital sources |

| Author, year | AW&R system name | Objective | Description | Diseases(s) 1 | Implementation level | Implementation dates of the AW&R system | AW&R System type | Purpose and scope | Alert mechanisms | Data sources |
|---|---|---|---|---|---|---|---|---|---|---|
| **Muriithi 2022**[145] | Brucellosis Surveillance | Not reported | Not reported | Brucellosis | Not reported | Not reported | Not reported | Disease outbreak surveillance | Automated alerts (use of technology / pre-defined communication channels like SMS); Manual notifications; Internal reports | Community sources; Hospital sources; Animal sources |
| **Murray 2013**[55] | Hospital based influenza surveillance | Not specified | It has been part of the national sentinel surveillance system for influenza. | SARS-CoV-2; Influenza | Not specified | January 2007 to July 2010 | Surveillance for human diseases | Disease burden & outbreak surveillance | Not specified | Hospital sources |
| **Mwatondo 2016**[56] | Integrated Disease Surveillance and Response | To strengthen the availability and use of surveillanc | IDSR seeks to capture health information of priority communica | IDSR Priority Diseases | County | Not specified | Surveillance for human diseases | Disease burden & outbreak surveillance | Automated alerts | Not specified |

| Author, year | AW&R system name | Objective | Description | Diseases(s) 1 | Implementation level | Implementation dates of the AW&R system | AW&R System type | Purpose and scope | Alert mechanisms | Data sources |
|---|---|---|---|---|---|---|---|---|---|---|
| | | e data for detecting, reporting, investigating, confirming, and responding to preventable priority diseases as well as other public health events | ble and non-communicable diseases for prevention and control by linking communities, health facilities, districts, counties and national levels. | | | | | | | |
| **Namuye 2015**[57] | Early Warning Alert and Response System | To enable a user to visualize risk and resource maps based on individual data in several | A freely available EWARS Spatial Fuzzy Logic Demo was developed which enables a user to visualize | **Cholera; Malaria; Typhoid Fever;** Tuberculosis; HIV/AIDS; Sexually Transmitted Diseases; Pneumonia | County | Not specified | Surveillance for human diseases | Disease burden & outbreak surveillance | Automated alerts | Media scanning |

| Author, year | AW&R system name | Objective | Description | Diseases(s) 1 | Implementation level | Implementation dates of the AW&R system | AW&R System type | Purpose and scope | Alert mechanisms | Data sources |
|---|---|---|---|---|---|---|---|---|---|---|
| | | data formats | risk and resource maps based on individual data in several data formats. | | | | | | | |
| **Ndegwa 2023**[58] | Mobile event-based surveillance system | To streamline the reporting process throughout the public health structures | M-Dharura is a mobile phone-based SMS application with a web-based interface that is part of the community health toolkit, offering open-source resources to design, build, and | COVID-19 | County | Not specified | Surveillance for human diseases | Disease burden & outbreak surveillance | Automated alerts | Community sources; Hospital sources |

| Author, year | AW&R system name | Objective | Description | Diseases(s)[1] | Implementation level | Implementation dates of the AW&R system | AW&R System type | Purpose and scope | Alert mechanisms | Data sources |
|---|---|---|---|---|---|---|---|---|---|---|
| | | | deploy digital tools for community health, specifically targeting caregivers in hard-to-reach areas and functioning with or without Internet connectivity. | | | | | | | |
| **Nderu 2019**[59] | Malaria surveillance | To assess the population structure of P. falciparum populations | Not specified | **Malaria** | County | Since 2013 | Surveillance for human diseases | Disease burden & outbreak surveillance | Not specified | Hospital sources |
| **Ng'eno 2023**[60] | Population Based Infectious | To monitor, detect, and respond to | A population of 26,000 | **Typhoid Fever** | Healthcare facility; Community | 2010 to 2019 | Surveillance for | Disease burden & outbreak | Manual notifications | Community sources; |

| Author, year | AW&R system name | Objective | Description | Diseases(s) [1] | Implementation level | Implementation dates of the AW&R system | AW&R System type | Purpose and scope | Alert mechanisms | Data sources |
|---|---|---|---|---|---|---|---|---|---|---|
| | Disease Surveillance | infectious diseases within a population by systematically collecting, analysing, and interpreting health data | individuals of all ages was monitored through household visits and clinic-based surveillance, where community interviewers collected self- and proxy-reported demographic information during regular visits. | | | | human diseases | surveillance | | Hospital sources |
| **Ng'etich 2021**[64] | Integrated Disease Surveillance and Response | To detect disease outbreaks | Health management committees operate at the sub- | IDSR Priority Diseases | Multi-county | Not specified | Surveillance for human diseases | Disease burden & outbreak surveillance | Not specified | Not specified |

| Author, year | AW&R system name | Objective | Description | Diseases(s) [1] | Implementation level | Implementation dates of the AW&R system | AW&R System type | Purpose and scope | Alert mechanisms | Data sources |
|---|---|---|---|---|---|---|---|---|---|---|
| | | | national level and are integrated into the health information systems. | | | | | | | |
| **Ng'etich 2021**[63] | Integrated Disease Surveillance and Response | To develop strategies to guide strengthening surveillance and response systems concerning PC-NTDs | Focused on diseases targeted for elimination or eradication. | **Schistosomiasis; Trachoma;** Lymphatic Filariasis | Multi-county | Not specified | Surveillance for human diseases | Disease burden & outbreak surveillance | Automated alerts; Manual notifications | Community sources; Hospital sources |
| **Ng'etich 2021**[62] | Preventive Chemotherapy for Neglected Tropical Diseases | To strengthen and improve surveillance systems | The IDSR system was a regional strategy to strengthen and improve surveillance systems | Neglected Tropical Diseases | Multi-county; County; Sub-county | 2015 to 2017 | Surveillance for human diseases | Disease burden & outbreak surveillance | Manual notifications | Hospital sources |

| Author, year | AW&R system name | Objective | Description | Diseases(s) 1 | Implementation level | Implementation dates of the AW&R system | AW&R System type | Purpose and scope | Alert mechanisms | Data sources |
|---|---|---|---|---|---|---|---|---|---|---|
| | | | and use of data for public health actions and response at all levels of the national system. | | | | | | | |
| **Ng'etich 2021**[61] | Preventive Chemotherapy for Neglected Tropical Diseases | To establish a surveillance system capable of detecting, confirming, reporting, analyzing, and interpreting surveillance data to inform response actions | The IDSR system surveillance functions comprise core functions and support functions. | Neglected Tropical Diseases | County | Not specified | Surveillance for human diseases | Disease burden & outbreak surveillance; Disaster preparedness | Automated alerts; Manual notifications | Community sources; Hospital sources |

| Author, year | AW&R system name | Objective | Description | Diseases(s) 1 | Implementation level | Implementation dates of the AW&R system | AW&R System type | Purpose and scope | Alert mechanisms | Data sources |
|---|---|---|---|---|---|---|---|---|---|---|
| **Ngugi 2018**[65] | Rabies surveillance | To describe the prevalence of animal bites and rabies vaccine administration | Not specified | Rabies | County | Not specified | Surveillance for human diseases; Animal diseases and disease syndromes surveillance | Disease burden & outbreak surveillance | Automated alerts | Hospital sources |
| **Ngugi 2020**[132] | Community Health Surveillance System (Kaloleni/Rabai) | To (i) strengthen the capacity of the local department of health for the collection, processing, and utilization of population-level health and vital events data, and | Trained Community Health Volunteers (CHVs) conduct biannual surveys across 112 villages within 10 Community Health Units (CHUs), collecting data on demograph | Not Specified | County | 2018 to date | Surveillance for human diseases | Disease burden & outbreak surveillance | Manual notifications; Public announcements | Community sources |

| Author, year | AW&R system name | Objective | Description | Diseases(s) [1] | Implementation level | Implementation dates of the AW&R system | AW&R System type | Purpose and scope | Alert mechanisms | Data sources |
|---|---|---|---|---|---|---|---|---|---|---|
| | | (ii) provide a platform that meets the University's needs for population-level research and academic programming | ics, health status, births, deaths, migration, orphanhood, and educational attendance among children. | | | | | | | |
| **Njenga 2021**[133] | Kenya Animal Bio surveillance System | To enhance disease surveillance among domestic and wild animal populations in Kenya | The surveillance system includes a mobile application for data collection, a web-based application for account management and data collection | **Rift Valley Fever** | Not specified | 2017 to 2019 | Animal diseases and disease syndromes surveillance; Climate surveillance | Disease burden & outbreak surveillance | Automated alerts | Animal sources |

| Author, year | AW&R system name | Objective | Description | Diseases(s) [1] | Implementation level | Implementation dates of the AW&R system | AW&R System type | Purpose and scope | Alert mechanisms | Data sources |
|---|---|---|---|---|---|---|---|---|---|---|
| | | | form editing, and a web-based dashboard for data viewing, monitoring, and downloading. | | | | | | | |
| **Njeru 2020**[66] | District Health Information System version 2 | To report disease outbreaks effectively | Key actors include county and sub county disease surveillance officers | Acute Flaccid Paralysis; **Ebola; Marburg; Lassa Fever; Crimean-Congo Haemorrhagic Fever; Anthrax; Cholera; Dengue Fever;** Shigella; Dracunculia | Multi-county | February and March 2017 | Surveillance for human diseases | Disease burden & outbreak surveillance | Automated alerts | Hospital sources |

| Author, year | AW&R system name | Objective | Description | Diseases(s) [1] | Implementation level | Implementation dates of the AW&R system | AW&R System type | Purpose and scope | Alert mechanisms | Data sources |
|---|---|---|---|---|---|---|---|---|---|---|
| | | | | sis; **Malaria;** Measles; **Meningococcal Meningitis; Neonatal Tetanus; Plague; Rift Valley Fever;** Severe Acute Respiratory Illness; Rabies; **Typhoid; Yellow Fever;** Tuberculosis | | | | | | |
| **Njuguna 2013**[67] | Population Based Infectious Disease Surveillance | Not specified | Trained community interviewers visited enrolled participants every two | Shigellosis | County | January 1, 2007, to December 31, 2010 | Surveillance for human diseases | Disease burden & outbreak surveillance | Manual notifications | Hospital sources |

| Author, year | AW&R system name | Objective | Description | Diseases(s) [1] | Implementation level | Implementation dates of the AW&R system | AW&R System type | Purpose and scope | Alert mechanisms | Data sources |
|---|---|---|---|---|---|---|---|---|---|---|
| | | | weeks from March 1, 2007, to January 31, 2010, and subsequently every week to collect data on illnesses and deaths since the last visit. | | | | | | | |
| **Njuguna 2014**[68] | Influenza virus surveillance | Not specified | The Kenya Ministry of Health (MoH), with technical support from the KEMRI/ CDC, initiated influenza sentinel surveillance at 11 | SARS-CoV-2; Influenza | County | Since 2006 | Surveillance for human diseases | Disease burden & outbreak surveillance | Automated alerts | Hospital sources |

| Author, year | AW&R system name | Objective | Description | Diseases(s) [1] | Implementation level | Implementation dates of the AW&R system | AW&R System type | Purpose and scope | Alert mechanisms | Data sources |
|---|---|---|---|---|---|---|---|---|---|---|
| | | | hospitals across the country. | | | | | | | |
| **Njuguna 2019**[69] | Health and Demographic Surveillance System (Kilifi) | Not specified | Not specified | **Malaria** | County | 2002 to 2016 | Surveillance for human diseases | Disease burden & outbreak surveillance | Not specified | Community sources; Hospital sources |
| **Nokes 2009**[70] | Hospital based RSV surveillance | To conduct surveillance of respiratory syncytial virus (RSV) infection | Integrated to Kilifi Epidemiological and Demographic Surveillance Study. | Respiratory Syncytial Virus | County; Healthcare facility | January 2002 to December 2007 | Surveillance for human diseases | Disease burden & outbreak surveillance | Not specified | Hospital sources |
| **Nyagah 2019**[71] | Mortuary based surveillance | To describe HIV/AIDS deaths | Cardiac blood samples were collected for this study using transthoracic aspiration and tested | HIV/AIDS | National | January 29 to March 3 2015 | Surveillance for human diseases | Disease burden & outbreak surveillance | Not specified | Hospital sources |

| **Author, year** | **AW&R system name** | **Objective** | **Description** | **Diseases(s)** [1] | **Implementation level** | **Implementation dates of the AW&R system** | **AW&R System type** | **Purpose and scope** | **Alert mechanisms** | **Data sources** |
|---|---|---|---|---|---|---|---|---|---|---|
| | | | for HIV/AIDS using the using the national diagnostic algorithm. | | | | | | | |
| **Nyawanda 2023**[72] | Population-based Infectious Disease Surveillance | To monitor several diseases, interventions and socio-economic factors within the population | Surveillance was conducted across 33 villages | **Malaria** | County; Sub-County | Since 2005 | Surveillance for human diseases; Climate surveillance | Disease burden & outbreak surveillance | Automated alerts | Hospital sources; Remote sensing data |
| **Odari 2014**[74] | Cholera Surveillance | To isolate and identify the strain of Vibrio cholera | Not specified | **Cholera** | Multi-county level | April and July 2007 | Surveillance for health events | Disease outbreak surveillance | Manual notifications | Hospital sources; Laboratory sources |
| **Ochieng 2013**[73] | Entomologic arbovirus surveillance | Not specified | Not specified | Arboviral infections | Multi-county | Not specified | Animal diseases and disease syndromes | Disease burden & outbreak surveillanc | Manual notifications | Animal sources |

| Author, year | AW&R system name | Objective | Description | Diseases(s) [1] | Implementation level | Implementation dates of the AW&R system | AW&R System type | Purpose and scope | Alert mechanisms | Data sources |
|---|---|---|---|---|---|---|---|---|---|---|
| | | | | | | | surveillance | e; Improve forecasting | | |
| **Odhiambo 2012**[134] | KEMRI/CDC Health and Demographic Surveillance System | To provide population-based information on disease burden | During the HDSS data collection, information on births, deaths, pregnancies, migrations, morbidity, parent survival status, immunization for children under 2 years, educational status, religion, marital status, and ethnicity is collected, along with | Influenza; Tuberculosis; HIV/AIDS; Acute Respiratory Infection; **Malaria; Diarrheal Diseases** | County | Since 2001 | Surveillance for human diseases | Disease burden & outbreak surveillance | Automated alerts | Community sources |

| Author, year | AW&R system name | Objective | Description | Diseases(s) [1] | Implementation level | Implementation dates of the AW&R system | AW&R System type | Purpose and scope | Alert mechanisms | Data sources |
|---|---|---|---|---|---|---|---|---|---|---|
| | | | house details and ITN information at the household level | | | | | | | |
| **Odhiambo 2015**[75] | Integrated Response System for Emerging Infectious Diseases | To improve the prediction and prevention of Rift Valley fever virus and other emerging arboviruses and to develop a model for response that could be expanded to other emerging diseases in | Not specified | Orthobunyavirus Infection | Not specified | January 2009 to April 2012 | Surveillance for human diseases | Disaster preparedness | Not specified | Laboratory sources |

| Author, year | AW&R system name | Objective | Description | Diseases(s) 1 | Implementation level | Implementation dates of the AW&R system | AW&R System type | Purpose and scope | Alert mechanisms | Data sources |
|---|---|---|---|---|---|---|---|---|---|---|
| | | the East African region. | | | | | | | | |
| **Oduor 2023**[76] | Population Based Infectious Disease Surveillance | Not specified | Surveillance occurs at the household level through regular visits by trained personnel using standardized questionnaires to collect data on births, deaths, and migrations | Infectious and non-infectious Diseases | County | Not specified | Surveillance for human diseases | Disease burden & outbreak surveillance | Manual notifications | Community sources |
| **Oduor 2023**[77] | Population Based Infectious Disease Surveillance | Not specified | Integrated with the Kenya Medical Research Institute | HIV/AIDS; Tuberculosis; Pneumonia; **Malaria** | Multi-county; Community | March 2020 to December 2021 | Surveillance for human diseases | Disease burden & outbreak surveillance | Not specified | Community sources |

| **Author, year** | **AW&R system name** | **Objective** | **Description** | **Diseases(s)** [1] | **Implementation level** | **Implementation dates of the AW&R system** | **AW&R System type** | **Purpose and scope** | **Alert mechanisms** | **Data sources** |
|---|---|---|---|---|---|---|---|---|---|---|
| | | | (KEMRI) and the Centres for Disease Control and Prevention (CDC). | | | | | | | |
| **Ogira 2022**[78] | Not specified | Not specified | Not specified | COVID-19 | National; County | Not specified | Surveillance for human diseases | Disaster preparedness | Manual notifications | Hospital sources |
| **Olack 2010**[79] | Population Based Infectious Disease Surveillance | Not specified | Integrated with the Kenya Medical Research Institute (KEMRI) and the Centres for Disease Control and Prevention (CDC). | Acute Febrile Illnesses; **Diarrheal Diseases** | Community | 2007 to 2010 | Surveillance for human diseases | Disease burden & outbreak surveillance | Not specified | Community sources |
| **Olotu 2010**[80] | Health Demographic | Not specified | All cohorts studied were nested | **Malaria** | County; Healthcare facility; Community | January 1998 to June 2009 | Surveillance for human diseases | Disease burden & outbreak | Not specified | Community sources; Hospital sources |

| Author, year | AW&R system name | Objective | Description | Diseases(s) [1] | Implementation level | Implementation dates of the AW&R system | AW&R System type | Purpose and scope | Alert mechanisms | Data sources |
|---|---|---|---|---|---|---|---|---|---|---|
| | Surveillance System | | within the wider demographic surveillance system (DSS). | | | | | surveillance | | |
| **Omemo 2012**[81] | Not specified | Not specified | Not specified | **Anthrax; Avian Influenza; Brucellosis; Rift Valley Fever**; rabies; Echinococcosis; Taeniasis; Swine flu | County | Not specified | Surveillance for human diseases; Animal diseases and disease syndromes surveillance | Disease burden & outbreak surveillance | Not specified | Community sources |
| **Omondi 2019**[146] | Integrated Disease Surveillance and Response | To utilize the Integrated Disease Surveillance and Response (IDSR) as a weekly reporting | The IDSR was integrated into the DHIS2 as a weekly reporting system. | Not Specified | National | Not specified | Surveillance for human diseases | Disease burden & outbreak surveillance | Manual notifications | Community sources; Hospital sources |

| Author, year | AW&R system name | Objective | Description | Diseases(s) [1] | Implementation level | Implementation dates of the AW&R system | AW&R System type | Purpose and scope | Alert mechanisms | Data sources |
|---|---|---|---|---|---|---|---|---|---|---|
| | | system, enabling surveillance officers to enter data into the reporting system for access by county and national health management teams | | | | | | | | |
| **Omondi 2020**[82] | Integrated Disease Surveillance and Response | To ensure the reporting of priority diseases, conditions, and events to the national level in accordance with Internation | Not specified | IDSR Priority Diseases | County | Not specified | Surveillance for human diseases | Disease burden & outbreak surveillance | Not specified | Community sources; Hospital sources |

| **Author, year** | **AW&R system name** | **Objective** | **Description** | **Diseases(s)** [1] | **Implementation level** | **Implementation dates of the AW&R system** | **AW&R System type** | **Purpose and scope** | **Alert mechanisms** | **Data sources** |
|---|---|---|---|---|---|---|---|---|---|---|
| | | al Health Regulations (IHR) requirements | | | | | | | | |
| **Omore 2024**[135] | Enterics for Global Health Shigella surveillance | To support incidence and consequences of Shigella diarrhea as part of multicountry surveillance aimed at preparing sites and assembling expertise for future Shigella vaccine trials | Not specified | Shigellosis | County | Not specified | Surveillance for human diseases | Disease burden & outbreak surveillance | Not specified | Community sources; Hospital sources |
| **Onyango 2012**[83] | Health and Demographic | Not specified | Not specified | **Avian Influenza** | County; Healthcare facility | 2007-2010 | Surveillance for | Disease burden & outbreak | Not specified | Hospital sources |

| Author, year | AW&R system name | Objective | Description | Diseases(s) 1 | Implementation level | Implementation dates of the AW&R system | AW&R System type | Purpose and scope | Alert mechanisms | Data sources |
|---|---|---|---|---|---|---|---|---|---|---|
| | Surveillance System (Kilifi) | | | | | | human diseases | surveillance | | |
| **Opiyo 2017**[147] | Kilifi Health and Demographic Surveillance System (KHDSS) | To create a community-based surveillance system linked to hospital-based disease surveillance | KHDSS covers 15 administrative locations in Kilifi County. It collects and records information about the patient's demographic information | **Malaria** | County-level | 2001-date | Surveillance for health events | Disease outbreak surveillance | Manual notifications | Community sources; Hospital sources |
| **Otambo 2023**[84] | Malaria surveillance | To ensure early detection of malaria cases, enabling prompt provision of appropriat | Active surveillance of malaria requires the collection of reliable and accurate | **Malaria** | County | Not specified | Surveillance for human diseases | Disease burden & outbreak surveillance | Not specified | Community sources |

| Author, year | AW&R system name | Objective | Description | Diseases(s) 1 | Implementation level | Implementation dates of the AW&R system | AW&R System type | Purpose and scope | Alert mechanisms | Data sources |
|---|---|---|---|---|---|---|---|---|---|---|
| | | e treatment | data to track disease trends, inform public health policies, and provide a solid evidence base for healthcare providers in making treatment and community case management decisions for malaria. | | | | | | | |
| **Otieno 2019**[85] | Flood impact-based | To assess the relationship between | The system used openly available | **Rift Valley Fever** | County | Not specified | Climate surveillance | Improve forecasting | Not specified | Climate sources; Remote sensing |

| Author, year | AW&R system name | Objective | Description | Diseases(s) [1] | Implementation level | Implementation dates of the AW&R system | AW&R System type | Purpose and scope | Alert mechanisms | Data sources |
|---|---|---|---|---|---|---|---|---|---|---|
| | forecasting system | flood exposure, vulnerability, coping capacity, and the impacts of flooding using openly available satellite rainfall data and other relevant datasets, thereby identifying critical factors that influence flood resilience in affected communities. | data on flood exposure, vulnerability, lack of coping capacity, flood impacts and observed satellite rainfall. | | | | | | | data; Secondary sources |

| Author, year | AW&R system name | Objective | Description | Diseases(s) [1] | Implementation level | Implementation dates of the AW&R system | AW&R System type | Purpose and scope | Alert mechanisms | Data sources |
|---|---|---|---|---|---|---|---|---|---|---|
| **Otieno 2022**[148] | Ecological Niche Model (ENM). | Prediction of the distribution of anthrax | ENM fits occurrence data as responses against several predictor variables. | Anthrax | National level | 2011-2017 | Surveillance for health events | Disease outbreak surveillance | Automated alerts (use of technology / pre-defined communication channels like SMS) | Hospital sources |
| **Ototo 2015**[86] | Not specified | To determine the impact of ITNs on indoor vector densities and biting behaviour in western Kenya | Adult mosquitoes were collected monthly using pyrethrum spray collections at six study sites in western Kenya from 2012 to 2014, with rotator trap collections conducted | **Malaria** | Multi-county | January 2012 to August 2014 | Surveillance for human diseases; Animal diseases and disease syndromes surveillance | Disease burden & outbreak surveillance; Improve forecasting | Manual notifications | Animal sources |

| Author, year | AW&R system name | Objective | Description | Diseases(s) [1] | Implementation level | Implementation dates of the AW&R system | AW&R System type | Purpose and scope | Alert mechanisms | Data sources |
|---|---|---|---|---|---|---|---|---|---|---|
| | | | in July–August 2013 and May–June 2014, and sampling occurring every 2 hours between 18:00 and 08:00 hrs. | | | | | | | |
| **Owor 2016**[87] | Not specified | Not specified | They conducted paediatric inpatient surveillance for HMPV at Kilifi County Hospital (KCH) in Coastal Kenya from 2007 to 2011. | Human Metapneumovirus infection | County | 2007 and 2011 | Surveillance for human diseases | Disease burden & outbreak surveillance | Manual notifications | Hospital sources |
| **Owuor 2022**[88] | Health facility- | To monitor and track | Integrated with the | **Avian Influenza** | Multi-county; | June 2009 and | Surveillance for | Disease burden & | Not specified | Hospital sources |

| Author, year | AW&R system name | Objective | Description | Diseases(s) [1] | Implementation level | Implementation dates of the AW&R system | AW&R System type | Purpose and scope | Alert mechanisms | Data sources |
|---|---|---|---|---|---|---|---|---|---|---|
| | based surveillance networks | influenza cases | CDC-Kenya surveillance system | | Healthcare facility | December 2018 | human diseases | outbreak surveillance | | |
| **Owuor 2023**[89] | Child Health and Mortality Prevention Surveillance Network | To document all causes of under-five mortality, prompt immediate public health actions, and develop interventions aimed at reducing future child mortality | Not specified. | Not specified | International; County | May 2017 to December 2021 | Surveillance for human diseases | Disease burden & outbreak surveillance | Not specified | Hospital sources |
| **Owusu 2024**[90] | Influenza and SARS-CoV-2 sentinel surveillance | To describe influenza and SARS-CoV-2 cocirculation in Kenya and how | At 8 sentinel influenza surveillance sites in Kenya, data on severe | Influenza; SARS-CoV-2 | National | Since 2007 | Surveillance for human diseases | Disease burden & outbreak surveillance | Not specified | Hospital sources |

| Author, year | AW&R system name | Objective | Description | Diseases(s) [1] | Implementation level | Implementation dates of the AW&R system | AW&R System type | Purpose and scope | Alert mechanisms | Data sources |
|---|---|---|---|---|---|---|---|---|---|---|
| | | the SARS-CoV-2 data from influenza sentinel surveillance correlated with that of universal national surveillance | acute respiratory illness or influenza-like illness is collected, including demographic, clinical, underlying medical conditions, vaccination, exposure information, and respiratory specimens. | | | | | | | |
| **Oyas 2018**[91] | Rift Valley Fever surveillance | To increase the chances of early detection of RVF disease in livestock (cattle, sheep, | Key actors were Kenya Directorate of Veterinary Services (KDVS, farmers and county | **Rift Valley Fever** | Multi-county; Sub-County; Community | November 2015 and February 2016 | Animal diseases and disease syndromes surveillance | Disease burden & outbreak surveillance | Automated alerts | Animal sources |

| Author, year | AW&R system name | Objective | Description | Diseases(s) 1 | Implementation level | Implementation dates of the AW&R system | AW&R System type | Purpose and scope | Alert mechanisms | Data sources |
|---|---|---|---|---|---|---|---|---|---|---|
| | | goats, and camels) | surveillance officers. | | | | | | | |
| **Park 2012**[92] | Modified TB treatment surveillance programme | To perform surveillance for multi drug resistant TB | Key actors include Moi University School of Medicine (MUSOM) and Moi Teaching and Referral Hospital (MTRH). | Tuberculosis | County; Healthcare facility | Not specified | Surveillance for human diseases | Disease burden & outbreak surveillance | Not specified | Hospital sources |
| **Pedro 2016**[93] | Rift Valley Fever Monitoring and Risk Mapping (RVFMRM) | To provide early warning and risk assessments for Rift Valley fever (RVF) outbreaks in Kenya and other regions susceptible to RVF in | The system integrates environmental, climatic, and epidemiological data to predict and map areas at risk of RVF outbreaks using GIS | Rift Valley Fever (RVF) | International level | Not specified | Surveillance for health events, Animal diseases and disease syndromes | Disease outbreak surveillance | Automated alerts (use of technology / pre-defined communication channels like SMS) | Environment sources; Climate sources; Animal sources, Remote sensing data |

| Author, year | AW&R system name | Objective | Description | Diseases(s) [1] | Implementation level | Implementation dates of the AW&R system | AW&R System type | Purpose and scope | Alert mechanisms | Data sources |
|---|---|---|---|---|---|---|---|---|---|---|
| | | Sub-Saharan Africa | and predictive modelling. It provides early warnings and supports active surveillance to help prevent and control RVF spread | | | | | | | |
| **Peletz 2018**[94] | Water quality monitoring system | To monitor water safety | Not specified | **Diarrheal Diseases** | International | Since 2013 | Environmental surveillance | Environmental monitoring | Not specified | Environment sources |
| **Pilgrim 2018**[95] | Early Warning Indicator Surveillance | To identify and correct gaps in ART program functioning to improve HIV care and treatment outcomes | The paediatric sites were a subset of the 32 sites, where NASCOP conducted the 2012 | HIV/AIDS | National; Healthcare facility | Not specified | Surveillance for human diseases | Antimicrobial resistance monitoring system | Not specified | Hospital sources |

| Author, year | AW&R system name | Objective | Description | Diseases(s) [1] | Implementation level | Implementation dates of the AW&R system | AW&R System type | Purpose and scope | Alert mechanisms | Data sources |
|---|---|---|---|---|---|---|---|---|---|---|
| | | | EWI monitoring assessments. | | | | | | | |
| **Post 2020**[96] | SARS-CoV-2 Surveillance System | To calculate the four novel surveillance indicators as weekly averages for the comparison of COVID-19 transmission rates on a week-to-week basis | The surveillance system data elements include the average weekly number of daily positive tests per 100,000 population, known as speed; the weekly average of day-to-day changes in the number of positives per day per 100,000 | SARS-CoV-2 Infection | National | Not specified | Surveillance for human diseases | Disease burden & outbreak surveillance | Not specified | Disease modelling |

| Author, year | AW&R system name | Objective | Description | Diseases(s) [1] | Implementation level | Implementation dates of the AW&R system | AW&R System type | Purpose and scope | Alert mechanisms | Data sources |
|---|---|---|---|---|---|---|---|---|---|---|
| | | | population, referred to as acceleration; the change in acceleration, termed jerk, which is the acceleration in the current week minus the acceleration in the prior week (with a sustained positive jerk typically indicating explosive growth); and the 7-day | | | | | | | |

| Author, year | AW&R system name | Objective | Description | Diseases(s) [1] | Implementation level | Implementation dates of the AW&R system | AW&R System type | Purpose and scope | Alert mechanisms | Data sources |
|---|---|---|---|---|---|---|---|---|---|---|
| | | | persistence, which measures how much an increase in speed from the previous week (last week's speed) persists into the current week by comparing it to the current speed of new COVID-19 cases reported per 100,000 population. | | | | | | | |
| **RinkedeWit 2022**[97] | Mobile event- | To enhance global | The Safe Care 4 | COVID-19 | National | Since 2017 | Surveillance for | Disease burden & | Automated alerts | Hospital sources |

| Author, year | AW&R system name | Objective | Description | Diseases(s) [1] | Implementation level | Implementation dates of the AW&R system | AW&R System type | Purpose and scope | Alert mechanisms | Data sources |
|---|---|---|---|---|---|---|---|---|---|---|
| | based surveillance system | COVID-19 preparedness and support for healthcare workers in low- and middle-income countries through initiatives such as SafeCare4Covid, Mom Care via the M-TIBA digital platform, CovidConnect for efficient case management, and the fully digitalized COVID-Dx | Covid App builds on the existing Pharm Access Safe Care approach, utilizing an IEEA-accredited, standards-based, stepwise quality improvement methodology, while Mom Care has been implemented since 2017 via the M-TIBA digital health platform, connecting | | | | human diseases | outbreak surveillance; Disaster preparedness | | |

| **Author, year** | **AW&R system name** | **Objective** | **Description** | **Diseases(s)** [1] | **Implementation level** | **Implementation dates of the AW&R system** | **AW&R System type** | **Purpose and scope** | **Alert mechanisms** | **Data sources** |
|---|---|---|---|---|---|---|---|---|---|---|
| | | patient journey tool | expectant mothers with healthcare providers and payers through their mobile phones. | | | | | | | |
| **Ruttoh 2023**[98] | SARS-CoV-2 Surveillance System | To detect community transmission of SARS-CoV-2 and monitor the co-circulation of SARS-CoV-2 with other acute respiratory pathogens | Sentinel surveillance maps the evolution of epidemics and provides evidence to inform control approaches in advance since hospital admissions and mortality | SARS-CoV-2; Respiratory Syncytial Virus; Influenza | County; Sub-County | January 2022 to December 2022 | Surveillance for human diseases | Disease burden & outbreak surveillance | Not specified | Hospital sources |

| Author, year | AW&R system name | Objective | Description | Diseases(s) 1 | Implementation level | Implementation dates of the AW&R system | AW&R System type | Purpose and scope | Alert mechanisms | Data sources |
|---|---|---|---|---|---|---|---|---|---|---|
| | | | indicators lag community transmission. | | | | | | | |
| **Sanders 1998**[99] | Rift Valley Fever surveillance | To implement hospital-based disease surveillance for Rift Valley Fever (RVF) | Hospital-based disease surveillance. | **Rift Valley Fever** | Multi-county | September 1992 to March 1993 | Surveillance for human diseases | Disease burden & outbreak surveillance | Manual notifications | Hospital sources |
| **Scobie 2017**[100] | Integrated Disease Surveillance and Response | To assess tetanus seroprotection levels by age group and sex among individuals in Kenya, Tanzania, and Mozambique | The system utilizes a multiplex bead assay (MBA) to evaluate seroprotection and immunity gaps against various diseases, including | **Tetanus** | International; County | May to July 2012 | Surveillance for human diseases | Disease burden & outbreak surveillance | Automated alerts; Internal reports | Community sources; Laboratory sources |

| Author, year | AW&R system name | Objective | Description | Diseases(s) [1] | Implementation level | Implementation dates of the AW&R system | AW&R System type | Purpose and scope | Alert mechanisms | Data sources |
|---|---|---|---|---|---|---|---|---|---|---|
| | | | tetanus, in the context of community surveys for neglected tropical diseases (NTDs). | | | | | | | |
| **Scott 2012**[136] | Health and Demographic Surveillance System (Kilifi) | To (i) define the incidence and prevalence of significant local childhood diseases, and (ii) evaluate the impact of new community-based interventions against infectious | The surveillance area was selected to capture most patients admitted to Kilifi District Hospital where morbidity events and vaccine coverage are linked in real time by a | Not Specified | County | 2000 to date | Surveillance for human diseases | Disease burden & outbreak surveillance | Automated alerts; Manual notifications | Hospital sources |

| Author, year | AW&R system name | Objective | Description | Diseases(s) 1 | Implementation level | Implementation dates of the AW&R system | AW&R System type | Purpose and scope | Alert mechanisms | Data sources |
|---|---|---|---|---|---|---|---|---|---|---|
| | | diseases while providing an epidemiological sampling frame | computer search of the population register. | | | | | | | |
| **Seheri 2018**[101] | World Health Organization's rotavirus surveillance | To document the impact of vaccination on disease burden and epidemiology | The system was coordinated by African Rotavirus Surveillance Network WHO-African region office. | **Rotavirus Diarrhoeal Disease** | International; multi-county | 2010 to 2015 | Surveillance for human diseases | Disease burden & outbreak surveillance | Not specified | Hospital sources |
| **Sewe 2016**[102] | Malaria surveillance | To investigate the relationship between malaria disease and | Surveillance took place in Asembo, Gen and Karemo (all part of the KEMRI/CDC | **Malaria** | County | 2003 to 2012 | Surveillance for human diseases; Environmental surveillance | Disease burden & outbreak surveillance; Environmental monitoring | Automated alerts | Remote sensing data |

| Author, year | AW&R system name | Objective | Description | Diseases(s) [1] | Implementation level | Implementation dates of the AW&R system | AW&R System type | Purpose and scope | Alert mechanisms | Data sources |
|---|---|---|---|---|---|---|---|---|---|---|
| | | weather conditions | HDSS areas). | | | | | | | |
| **Sewe 2017**[103] | Malaria surveillance | To develop forecasting models for malaria | Not specified | **Malaria** | County | Not specified | Climate surveillance | Improve forecasting | Not specified | Remote sensing data |
| **Sharma 2015**[104] | Treatment Information from Basic Unit | To store details on individual patient episodes of TB reported to the national TB program | TIBU is a national case-based surveillance system that has provided nationwide coverage since 2012, storing details on individual TB patient episodes reported to the national TB program, including demographic | Tuberculosis | County | Since 2012 | Surveillance for human diseases | Disease burden & outbreak surveillance | Automated alerts | Hospital sources |

| Author, year | AW&R system name | Objective | Description | Diseases(s) [1] | Implementation level | Implementation dates of the AW&R system | AW&R System type | Purpose and scope | Alert mechanisms | Data sources |
|---|---|---|---|---|---|---|---|---|---|---|
| | | | characteristics, clinical details, laboratory results, treatment outcomes, and patient locating information. | | | | | | | |
| **Shih 2022**[105] | Acute febrile illness surveillance | To detect and characterize SARS-CoV-2 infections | SARS-CoV-2 testing was integrated into the AFI surveillance systems. | COVID-19 | International | Since May 2020 | Surveillance for human diseases | Disease burden & outbreak surveillance | Automated alerts | Hospital sources; Laboratory sources |
| **Sifuna 2014**[106] | Health and Demographic Surveillance System (Kombewa) | To conduct population and disease surveillance through the Kombewa Health and Demographic Surveillanc | Part of Kombewa Clinical Research Centre with collaboration with United States Army Medical | **Diarrhoea; Fever;** Flu/Cold; **Malaria;** HIV/AIDS | County; Community | Not specified | Surveillance for human diseases | Disease burden & outbreak surveillance | Not specified | Community sources |

| Author, year | AW&R system name | Objective | Description | Diseases(s) [1] | Implementation level | Implementation dates of the AW&R system | AW&R System type | Purpose and scope | Alert mechanisms | Data sources |
|---|---|---|---|---|---|---|---|---|---|---|
| | | e System (HDSS) by integrating household-based and health facility-based data, with a focus on the health conditions of the population | Research Unit-Kenya (USAMRU-K) and the Kenya Medical Research Institute (KEMRI). | | | | | | | |
| **Sirengo 2016**[107] | Routine HIV/AIDS testing surveillance | To estimate HIV prevalence among pregnant women | The system uses routinely collected HIV testing data from ANC to estimate HIV prevalence among pregnant women. | HIV/AIDS | National | Not specified | Surveillance for human diseases | Disease burden & outbreak surveillance | Not specified | Hospital sources |

| Author, year | AW&R system name | Objective | Description | Diseases(s) [1] | Implementation level | Implementation dates of the AW&R system | AW&R System type | Purpose and scope | Alert mechanisms | Data sources |
|---|---|---|---|---|---|---|---|---|---|---|
| **Soti 2015**[108] | Mobile event-based surveillance system | To enable health program stakeholders to remotely monitor diagnostic performance, adherence to case management protocols, healthcare workers' activity levels, and other health performance indicators through the Fionet portal | Fionet is an innovation that integrates diagnostics, data capture, and cloud services within its malaria control program to demonstrate usability and feasibility for primary level workers in a remote setting in Kenya. | **Malaria** | County; Sub-County; Healthcare facility | Not specified | Surveillance for human diseases | Disease burden & outbreak surveillance | Automated alerts | Hospital sources |
| **Soura 2015**[109] | Urban Health and | To establish | Not specified | Not Specified | Community | Since 2002 | Surveillance for | Disease burden & | Not specified | Community sources |

| **Author, year** | **AW&R system name** | **Objective** | **Description** | **Diseases(s)** [1] | **Implementation level** | **Implementation dates of the AW&R system** | **AW&R System type** | **Purpose and scope** | **Alert mechanisms** | **Data sources** |
|---|---|---|---|---|---|---|---|---|---|---|
| | Demographic Surveillance Systems (Nairobi) | research and intervention platforms addressing population and health issues | | | | | human diseases | outbreak surveillance | | |
| **Stehling-Ariza 2023**[137] | Acute flaccid paralysis surveillance | To detect poliovirus | Integrated to the World Health Organization's surveillance. | Poliomyelitis | International | 2021 to 2022 | Surveillance for human diseases | Disease burden & outbreak surveillance | Not specified | Not specified |
| **Swierczewski 2013**[110] | Enteric Disease Surveillance | To accurately predict the frequency of pathogens and potential changes in antibiotic resistance patterns | Not specified | **Enteric Diarrheal Diseases;** Shigellosis | Multi-county; Healthcare facility | 25 September 2009 to 30 September 2011 | Surveillance for human diseases | Disease burden & outbreak surveillance | Not specified | Hospital sources |

| Author, year | AW&R system name | Objective | Description | Diseases(s) 1 | Implementation level | Implementation dates of the AW&R system | AW&R System type | Purpose and scope | Alert mechanisms | Data sources |
|---|---|---|---|---|---|---|---|---|---|---|
| **Taylor 2020**[156] | Child Health and Mortality Prevention Surveillance Network | To gather standardized, population-based, longitudinal data on under-5 mortality and stillbirths in sub-Saharan Africa and South Asia to enhance the precision of identifying causes of death | The CHAMPS Network was established to collect standardized, population-based longitudinal data at sites in areas with high child mortality, aiming to understand and track preventable causes of childhood deaths and stillbirths globally | Not Specified | Not specified | Dec 10, 2016, to Dec 31, 2018 | Surveillance for human diseases | Disease burden & outbreak surveillance | Automated alerts | Community sources; Hospital sources |
| **Tesfaye 2020**[157] | Acute flaccid paralysis | To detect, investigate, report, | Key actors included healthcare | Acute Flaccid Paralysis | National | January 2016 to | Surveillance for | Disease burden & outbreak | Not specified | Hospital sources |

| Author, year | AW&R system name | Objective | Description | Diseases(s) [1] | Implementation level | Implementation dates of the AW&R system | AW&R System type | Purpose and scope | Alert mechanisms | Data sources |
|---|---|---|---|---|---|---|---|---|---|---|
| | surveillance | disseminate information, and promptly implement control measures for indigenous and imported cases of wild poliovirus (WPV) or circulating vaccine-derived poliovirus (cVDPV) | workers and disease surveillance officers | | | December 2018 | human diseases | surveillance | | |
| **Thomas 2021**[113] | Cross-sectoral zoonotic disease surveillance | To improve public health outcomes through the rapid detection | Joint outbreak responses between human and animal health | Zoonotic Diseases | County; Sub-County | Not specified | Surveillance for human diseases; Animal diseases and disease | Disease burden & outbreak surveillance | Not specified | Community sources; Hospital sources; Animal sources |

| Author, year | AW&R system name | Objective | Description | Diseases(s) 1 | Implementation level | Implementation dates of the AW&R system | AW&R System type | Purpose and scope | Alert mechanisms | Data sources |
|---|---|---|---|---|---|---|---|---|---|---|
| | | of zoonotic disease events prior to widespread transmission in humans | sectors at the national and sub-national level. | | | | syndromes surveillance | | | |
| **Thumbi 2015**[114] | Population-based animal syndromic surveillance; Population-based infectious disease surveillance | To investigate and understand the links between human health, animal health | This platform for integrated health and economic welfare analysis addresses the challenge by simultaneously collecting data from a population-based animal syndromic | Human Syndromes; Fever, Jaundice; **Diarrhea;** Respiratory Illness and Animal Syndromes; Death; Respiratory; Reproductive; Musculoskeletal; Nervous; Urogenital; Digestive; Udder | County | Since 2001 | Surveillance for human diseases; Animal diseases and disease syndromes surveillance | Disease burden & outbreak surveillance | Automated alerts; Manual notifications; Internal reports | Community sources; Animal sources |

| Author, year | AW&R system name | Objective | Description | Diseases(s) [1] | Implementation level | Implementation dates of the AW&R system | AW&R System type | Purpose and scope | Alert mechanisms | Data sources |
|---|---|---|---|---|---|---|---|---|---|---|
| | | | surveillance (PBASS) system and an ongoing human population-based infectious disease surveillance (PBIDS) system within the same rural households in Western Kenya, linking this data to a quantitative assessment of socio-economic dynamics over time. | Disorders; Skin Disorders in Cattle; Sheep; Goats; Chickens | | | | | | |
| **Thumbi 2019**[115] | Not specified | To compare | The mobile phone- | Animal Syndromes; | Not specified | Not specified | Animal diseases | Disease burden & | Automated alerts | Community sources |

| Author, year | AW&R system name | Objective | Description | Diseases(s) [1] | Implementation level | Implementation dates of the AW&R system | AW&R System type | Purpose and scope | Alert mechanisms | Data sources |
|---|---|---|---|---|---|---|---|---|---|---|
| | | syndromic surveillance data for livestock diseases collected through household visits and a mobile phone-based method to investigate the temporal patterns of reported disease events and the accuracy of both methods | based surveillance system was established with a toll-free line and four "hunting lines" to allow up to five simultaneous callers, each managed by a study staff member, while information about the toll-free number was shared with households via cards | Death; Respiratory; Reproductive; Musculoskeletal; Nervous; Urogenital; Digestive; Udder Disorders; Skin Disorders in Cattle; Sheep; Goats; Chickens | | | and disease syndromes surveillance | outbreak surveillance | | |

| Author, year | AW&R system name | Objective | Description | Diseases(s) 1 | Implementation level | Implementation dates of the AW&R system | AW&R System type | Purpose and scope | Alert mechanisms | Data sources |
|---|---|---|---|---|---|---|---|---|---|---|
| | | | instructing farmers to report illness or death at no charge. | | | | | | | |
| **Toda 2016**[116] | Mobile short messaging service-based disease outbreak alert system | To test the effectiveness of a mobile short-message-service (SMS)-based disease outbreak alert system (mSOS) for reporting immediately notifiable diseases | mSOS is a formatted text-messaging system that enables communications between healthcare facility workers and Ministry of Health managers and uses a Web-based portal to monitor disease notification | Not Specified | National; County; Sub-County | Not specified | Surveillance for human diseases | Disease burden & outbreak surveillance | Automated alerts | Hospital sources |

| Author, year | AW&R system name | Objective | Description | Diseases(s)[1] | Implementation level | Implementation dates of the AW&R system | AW&R System type | Purpose and scope | Alert mechanisms | Data sources |
|---|---|---|---|---|---|---|---|---|---|---|
| | | | s and response actions taken by health manager. | | | | | | | |
| **Toda 2017**[117] | Integrated Disease Surveillance and Response | To serve as a disease outbreak alert system | mSOS consisted of formatted short-message-service (SMS or text messaging) communication between health workers at local facilities and MOH health managers comprising of disease | IDSR Priority Diseases | County; Sub-County; Healthcare facility | 2013 to 2014 | Surveillance for human diseases | Disease burden & outbreak surveillance | Automated alerts | Hospital sources |

| Author, year | AW&R system name | Objective | Description | Diseases(s) 1 | Implementation level | Implementation dates of the AW&R system | AW&R System type | Purpose and scope | Alert mechanisms | Data sources |
|---|---|---|---|---|---|---|---|---|---|---|
| | | | surveillance coordinators at the national, county, and sub county levels. | | | | | | | |
| **Toda 2018**[118] | Mobile short messaging service-based disease outbreak alert system | Not specified | Not specified | Shigellosis; Measles; **Dengue Fever** | County | Not specified | Surveillance for human diseases | Disease burden & outbreak surveillance | Automated alerts | Hospital sources |
| **Umuhoza 2020**[119] | Influenza virus surveillance | To identify changes in circulating influenza virus subtypes and genotype strains which may impact | USAMRU-K, Kenya Defence Forces (KDF), Kenya Medical Research Institute (KEMRI), the | **Avian Influenza** | Multi-county; Healthcare facility | January 2007 to December 2013 | Surveillance for human diseases | Disease burden & outbreak surveillance | Not specified | Hospital sources |

| Author, year | AW&R system name | Objective | Description | Diseases(s) [1] | Implementation level | Implementation dates of the AW&R system | AW&R System type | Purpose and scope | Alert mechanisms | Data sources |
|---|---|---|---|---|---|---|---|---|---|---|
| | | disease severity, transmissibility, and treatment and prevention effectiveness across time and geography | Ministry of Health (MoH), and WHO collaborate, and this system complements the one established by CDC Kenya for provincial hospitals. | | | | | | | |
| **VanHemelrijck 2009**[120] | Longitudinal health facility surveillance | Not specified | Not specified | Upper Respiratory Tract Infection; **Malaria** | County | 1997 to 2006 | Surveillance for human diseases | Disease burden & outbreak surveillance | Automated alerts; Manual notifications | Hospital sources |
| **VanZyl 2019**[158] | Bag-mediated filtration environmental surveillance | To detect poliovirus, an enteric virus, in environmental waters, with initial testing | The bag-mediated filtration system (BMFS), a novel environmental surveillanc | **Human Enteric Viruses;** Hepatitis A | County | 14 April 2015 to 16 April 2016 | Environmental surveillance | Environmental monitoring | Not specified | Environment sources |

| Author, year | AW&R system name | Objective | Description | Diseases(s) [1] | Implementation level | Implementation dates of the AW&R system | AW&R System type | Purpose and scope | Alert mechanisms | Data sources |
|---|---|---|---|---|---|---|---|---|---|---|
| | | conducted in Kenya | e method first tested in Kenya for detecting poliovirus in environmental waters, enables in-field collection and filtration of large sample volumes (3–6 L) of wastewater or wastewater-impacted waters. | | | | | | | |
| **Walker 2014**[122] | Acute flaccid paralysis surveillance | To detect paralytic polio cases | AFP surveillance systems are expected | Acute Flaccid Paralysis | County | Not specified | Surveillance for human diseases | Disease burden & outbreak surveillance | Not specified | Hospital sources |

| Author, year | AW&R system name | Objective | Description | Diseases(s) 1 | Implementation level | Implementation dates of the AW&R system | AW&R System type | Purpose and scope | Alert mechanisms | Data sources |
|---|---|---|---|---|---|---|---|---|---|---|
| | | | to identify at least 2 nonpolio AFP (NPAFP) cases per 100,000 population under 15 years of age annually to demonstrate their sensitivity in detecting paralytic polio cases. | | | | | | | |
| **Wathondu 2016**[149] | Malaria surveillance | To predict the occurrence of malaria cases/ epidemics | NR | **Malaria** | Sub-county level | NR | Surveillance for health events | Disease outbreak surveillance | Not specified | Community sources; Hospital sources |
| **Worsley-Tonks 2022**[123] | Kenya Animal Bio | To provide early warning of | The Kenya Animal Bio surveillanc | Zoonotic Diseases | National | Not specified | Animal diseases and disease | Disease burden & outbreak | Not specified | Community sources; |

| **Author, year** | **AW&R system name** | **Objective** | **Description** | **Diseases(s) 1** | **Implementation level** | **Implementation dates of the AW&R system** | **AW&R System type** | **Purpose and scope** | **Alert mechanisms** | **Data sources** |
|---|---|---|---|---|---|---|---|---|---|---|
| | surveillance System | emerging zoonotic diseases | e System (KABS) is an integrated human and animal syndromic surveillance system that targets disease syndromes prioritized by national health agencies, enabling the rapid transfer of health data from across the country to health authorities. | | | | syndromes surveillance | surveillance | | Animal sources |
| **Zhou 2015**[124] | Malaria surveillance | To monitor malaria transmission | Not specified | **Malaria** | Multi-county; Healthcare facility; Community | February to March and from May to June in | Surveillance for human diseases | Disease burden & outbreak surveillance | Not specified | Community sources; Hospital sources |

| Author, year | AW&R system name | Objective | Description | Diseases(s) [1] | Implementation level | Implementation dates of the AW&R system | AW&R System type | Purpose and scope | Alert mechanisms | Data sources |
|---|---|---|---|---|---|---|---|---|---|---|
| | | | | | | both 2009 and 2010 | | | | |
| **Zhou 2020**[125] | Bag-mediated filtration environmental surveillance | To facilitate poliovirus (PV) environmental surveillance as a supplement to acute flaccid paralysis surveillance in poliovirus eradication efforts | The bag-mediated filtration system (BMFS) was developed to facilitate poliovirus (PV) environmental surveillance, a supplement to acute flaccid paralysis surveillance in PV eradication efforts. | Poliomyelitis | County | April to September 2015 | Environmental surveillance | Environmental monitoring | Not specified | Environment sources |

**Table 4: Outcome Features of the Advance Warning and Response Systems**

| Study ID | AW&R system name | Key performance indicators | Success extent | Lessons learnt |
|---|---|---|---|---|
| **Abidha 2017**[138] | Kilifi Health and Demographic Surveillance System (KHDSS) | Not reported | Not reported | Not reported |
| **Adazu 2005**[1] | Health and Demographic Surveillance System | Mortality rates; morbidity rates; cause of mortality; entomologic parameters of malaria transmission; fertility; migration; hospital visits; socioeconomic status. | Within one year the surveillance was able to detect high mortality rates, especially in children and young adults. | Long-term data collection poses challenges such as high costs, the need for adequate field staff and supervision, extensive training before each data collection round, weekly meetings to ensure adherence to SOPs, quality control measures like repeat interviews and detecting inaccuracies, and sophisticated computer systems for data management. |
| **Anyamba 2010**[2] | Rift Valley Fever surveillance | Improved temporal warnings, early disease detection, and implementation of control strategies during outbreaks. | The system successfully predicted RVF outbreaks. | Approximately half of all recorded cases of illness and death in animals were reported through a toll-free number, highlighting it as a relatively inexpensive and efficient alternative surveillance tool. |
| **Arale 2019**[3] | Acute flaccid paralysis surveillance | Increase surveillance sensitivity. | The recruitment of CHVs serves as a link between the hard-to-reach nomadic | The capacity developed through these committees for polio eradication can also |

| Study ID | AW&R system name | Key performance indicators | Success extent | Lessons learnt |
|---|---|---|---|---|
| | | | communities and health facilities. | support other health initiatives. |
| **Bello 2021**[4] | Integrated supportive supervision | Proportion of health care workers with adequate knowledge of AFP; measles case definition in the health facilities. | Not specified | Countries could explore opportunities for maintaining supportive supervision during the pandemic by utilizing virtual remote supervision when in-person visits are not feasible. |
| **Bigogo 2018**[5] | Population Based Infectious Disease Surveillance | Not specified | Greater TB detection rates among participants screened for TB at health facilities compared to those screened at home. | A combined strategy that utilizes both intensified facility-based screening and community outreach may provide a comprehensive approach to TB control, addressing barriers and reaching underserved populations. |
| **Bisanzio 2015**[6] | Surveillance of febrile cases | Prevalence of malaria; mosquito infestation rates; coverage, usage, and impact of LLINs in the target population; and the number of febrile cases seeking treatment at health facilities. | Better detection of malaria cases through georeferenced data and monitoring temporal and spatial patterns of malaria prevalence effectively. The system successfully identified geographical hotspots of malaria risk. | The use of georeferenced data for monitoring malaria prevalence allows for a better understanding of spatial and temporal patterns. |
| **Bolu 2007**[7] | Antenatal care sentinel surveillance | HIV/AIDS prevalence. | Another benefit is the capacity to compare the prevalence among those who accept HIV testing under PMTCT and those who either | ANC sentinel surveillance can enhance PMTCT programs by incorporating PMTCT variables into ANC data collection forms or by comparing both data |

| Study ID | AW&R system name | Key performance indicators | Success extent | Lessons learnt |
|---|---|---|---|---|
| | | | refuse or who are not offered the test. | sets for monitoring and evaluation. |
| **Boru 2023**[8] | Integrated Disease Surveillance and Response | Not specified | Not specified | Not specified |
| **Breiman 2014**[9] | Population Based Infectious Disease Surveillance | Not specified | Estimations of the number of people affected by a disease and project the number of illness episodes that can be prevented by applying interventions of known effectiveness. | Rotavirus significantly contributes to gastroenteritis in adults in Kenya. |
| **Byrne 2020**[126] | Community Epidemic and Pandemic Preparedness Programme | Coverage; accuracy and validity; timeliness; response and action. | Not specified | CBS systems must be tailored to each context, as there is no one-size-fits-all model; each country has unique reporting, investigation, and response systems, as well as varying cultural and socio-economic factors at the community level. |
| **Campbell 2019**[10] | Rift Valley Fever surveillance | Accuracy and reliability of model predictions for Aedes mcintoshi mosquito abundances over time and space; disease incidence rates; ability of the model to correlate mosquito abundance with environmental and climatic factors (rainfall patterns and temperature variations). | Not specified | Increased mosquito sampling is needed to accurately capture the environmental conditions associated with high mosquito abundances. However, the model's limitations, including its failure to account for mosquito density dependence, must also be acknowledged. |

| Study ID | AW&R system name | Key performance indicators | Success extent | Lessons learnt |
|---|---|---|---|---|
| **Cheluget 2004**[11] | HIV/AIDS sentinel surveillance | Incidence of STDs. | Proportion of patients with STDs with specific STD syndromes declined from 1994 to 2000 and increased again in 2001. | Not specified |
| **Curran 2018**[12] | Cholera outbreak surveillance | Response speed. | Contact tracing and administration of antibiotic prophylaxis to family members and neighbours of cholera patients as a success of the cholera response. Health education was conducted both in the communities and at healthcare facilities. Strong collaboration- CHEWS, nurses. | During health system transformations, there is a need to adapt, implement, and communicate health strategies, including public health emergency preparedness, at the county level to effectively inform and train healthcare workers. |
| **Ear 2013**[139] | Global Emerging Infections Surveillance and Response System | Not specified | The existence of USAMRU in Kenya has strengthened local governmental institutions. | To further enhance surveillance success rates, it is important to involve the populace, either as part of the surveillance team or as security officers. |
| **Fagnant 2018**[13] | Bag-mediated filtration environmental surveillance | Duration of clarification; sensitivity of the BMFS. | BMFS results in an increase of 5 to 16.7times the effective volume assayed compared to volumes assayed for 500-mL two-phase grab sample. | Not specified |
| **Falzon 2019**[14] | Integrated surveillance system for zoonotic diseases | Efficacy; cost effectiveness; coverage and effort. | The novel design of this surveillance system allows us to investigate multiple zoonoses in livestock and | Engaging with local stakeholders in the field and providing timely feedback through regular public engagement sessions are |

| Study ID | AW&R system name | Key performance indicators | Success extent | Lessons learnt |
|---|---|---|---|---|
| | | | human populations living in the area. | essential for ensuring ongoing compliance. |
| **Geteri 2024**[15] | Integrated Disease Surveillance and Response | Outbreak detection. | Identified that improved surveillance rather than an increase in the occurrence of diseases led to the rise in reports over time. | The increase in the magnitude of reported outbreaks is indicative of both enhanced surveillance capabilities and a greater awareness of disease management. |
| **Feikin 2010**[79] | Population Based Infectious Disease Surveillance | Prevalence and incidence of infectious disease syndromes every 2 weeks. | Not specified | The period of recall should not exceed 3 days in children and 4 days in adults to identify at least 80% of the maximum rate of disease. |
| **Feikin 2011**[17] | Population Based Infectious Disease Surveillance | Clinic visitation rates; household morbidity; mortality; burden projections. | The system successfully gathered comprehensive data on morbidity and mortality, capturing various diseases across both urban (Kibera) and rural (Asembo) settings. | There is a significant underreporting of illness in rural areas like Asembo, where many disease episodes go undetected at clinics, highlighting the need for better surveillance and health care access. |
| **Fujii 2014**[18] | Health and Demographic Surveillance System | Participation rates from the two HDSS sites; seropositivity rates for various pathogens; sensitivities and specificities of the antigen detection. | Not specified | While the low cost of materials (around one dollar per sample) enhances the feasibility of widespread surveillance in resource-limited settings, the high initial investment in assay equipment presents a significant barrier. |

| Study ID | AW&R system name | Key performance indicators | Success extent | Lessons learnt |
|---|---|---|---|---|
| **Furukawa 2016**[19] | Population Based Infectious Disease Surveillance | Not specified | Not specified | Not specified |
| **Gachohi 2012**[20] | Rift Valley Fever surveillance | Responses to warnings. | Forty-two percent (22/52) of the respondents received a warning of a heightened RVF risk in 2008. The majority (59%, 13/22) of them obtained the warnings from the directorate of veterinary services headquarters in April 2008. | Warnings regarding the disease were, however, issued late on a short timeline. |
| **Gardner 2018**[127] | Not specified | The number of non-polio AFP; collection of adequate stool specimens from 80% of patients with AFP. | National NPAFP rate of 2.8 identified, 89% of national areas with NPAFP rate of 2. | Integrating poliovirus surveillance with other vaccine-preventable diseases is crucial to sustain capacity and ensure performance quality, while routinely assessing surveillance data to identify suboptimal quality and reaching geographically challenging, security-compromised areas, as well as mobile and migrant populations. |
| **Geiger 2024**[128] | Global Polio Laboratory Network | Number of non-polio AFP cases; percentage of AFP cases with stool specimens collected within 14 days of onset; percentage of AFP cases with two stool specimens collected 24-48 hours apart; extent of surveillance | Kenya reported 0 cases of WPV1 and cVDPV in 2022 and 2023 but reported 8 case of cVDPV in 2023. | Not specified |

| Study ID | AW&R system name | Key performance indicators | Success extent | Lessons learnt |
|---|---|---|---|---|
| | | activities across different regions and populations. | | |
| **Gerken 2022**[21] | Rift Valley Fever surveillance | Sample collection and seroprevalence. | The study involved stakeholders from slaughterhouses and livestock markets, who participated in mapping livestock movement routes and shared insights into animal selection and transportation. | There is need for improved veterinary inspection and management to ensure animal health during transport. |
| **Gikungu 2016**[22] | Rift Valley Fever surveillance | Positive outbreak predictions; rainfall and minimum temperature. The predictor variables were rainfall, NDVI, relative humidity at 06:00 and at 12:00 GMT, maximum and minimum temperatures and SST. | Positive predictions that were confirmed by reports of actual outbreaks, especially the 1983, 1989, 1993, 1997-98 and 2006-07 RVF outbreaks. Rainfall and minimum temperature were found to be the significant predictors of the model | Regular surveillance can enhance livestock immunity and mitigate the risk of RVF outbreaks, even under favourable meteorological conditions for infection. |
| **Githinji 2014**[23] | Mobile short Message Service for Life system | Response rates: timeliness and accuracy of SMS reported surveillance data. | Eighty-seven per cent of facilities responded within 27-hour incentive period, 9% responded after the incentive period while 4% did not respond at all. | Delayed response rates were noted in weeks 5 and 25 due to network issues. |
| **Githure 2022**[24] | Malaria surveillance | Mosquito collection efficacy; reduction in malaria transmission and incidence; positivity rates from diagnostic methods; cost per person protected; data monitoring and reporting for malaria burden. | Successful introduction of PBO nets in specific counties, leading to significant reductions in malaria incidence. | A substantial number of malaria cases go unreported due to poor treatment-seeking behaviours, underscoring the importance of enhancing community awareness. |

| Study ID | AW&R system name | Key performance indicators | Success extent | Lessons learnt |
|---|---|---|---|---|
| **Hammitt 2019**[25] | Health and Demographic Surveillance System (Kilifi) | Vaccination coverage; incidence of invasive pneumococcal disease; carriage surveys. | The introduction of PCV10 led to a dramatic decrease in the incidence of VT-IPD, indicating a successful impact on public health. | Continuous monitoring of vaccine impact on disease incidence and carriage patterns is crucial for adapting public health strategies. |
| **Harris 2020**[26] | Malaria surveillance | Analysis of the data distributions and trends through variance, decay time or mean index of dispersion. | Nine of the 10 statistical indicators increased during the approach to criticality, as predicted by the theory of critical slowing down. | Not specified |
| **Hassan 2020**[27] | Community based syndromic surveillance | Number of suspected/confirmed human cases with RVF; number of mortalities linked to the outbreak; number of suspected livestock cases (including history of abortions or deaths of young animals and haemorrhagic syndrome); number of confirmed livestock cases; and duration between the detection of a suspected first case and the first laboratory-confirmed case. | The investigation established a 30 delay between the detection of the suspected first case and the first laboratory-confirmed cases. | The outbreak in Siaya highlighted the spread of RVF into previously low-risk ecological niches, revealing a 30-day delay between the detection of the suspected first case and the first laboratory-confirmed case, likely due to a low suspicion index among health workers and limited capacity for differential diagnosis with other acute febrile illnesses. |
| **Hay 1998**[28] | Malaria surveillance | Not specified | Not specified | The lack of basic epidemiological tools, such as disease risk maps, in many national malaria control programs presents significant challenges to effective malaria management and control. |
| **Hay 2000**[29] | Not specified | Annual and super annual variations in disease incidence, with approximately | Not specified | Intrinsic population dynamics between host and pathogen |

| Study ID | AW&R system name | Key performance indicators | Success extent | Lessons learnt |
|---|---|---|---|---|
| | | 3-year interepidemic periods for both diseases. | | populations provide a more plausible explanation for the periodic outbreaks, which is crucial for enhancing epidemic prediction models. |
| **Hendriksen 2019**[30] | Population Based Infectious Disease Surveillance | Weekly changes in pathogen abundance; read abundance flagged as potential outbreaks. | The metagenomics approach showed potential as an early warning system by detecting upsurges in pathogen levels before widespread clinical symptoms were reported. | Metagenomics has the potential to provide complete taxonomic and functional profiles of environmental and human microbiomes and epistomes. |
| **Homan 2016**[129] | Health and Demographic Surveillance System (Rusinga) | Coverage of reporting sites per year; data completeness for the years 2013 and 2014; sensitivity of outbreak detection; and specificity of outbreak detection. | Complete demographic data was collected for the entire population in the HDSS area. | Not specified |
| **Iacono 2018**[31] | Eco-epidemiological, compartmental, mathematical model | "1. Population Dynamics: Understanding the population dynamics of Aedes and Culex mosquito species, including their life stages. | Not specified | Eco-epidemiological, compartmental, mathematical model |
| **Iuliano 2018**[32] | Influenza virus surveillance | Mortality estimates for influenza-associated respiratory deaths. | Not specified | The research encountered data limitations due to the lack of high-quality mortality data, particularly in lower-income countries, and revealed considerable variability in influenza-associated deaths across years, likely stemming from differences in circulating virus strains and their severity, |

| Study ID | AW&R system name | Key performance indicators | Success extent | Lessons learnt |
|---|---|---|---|---|
| | | | | highlighting the necessity for ongoing monitoring of these strains and their health impacts. |
| **Jebiwot 2018**[140] | Therapeutic efficacy studies (TES) | Not reported | Not reported | Not reported |
| **Joesoef 2003**[33] | HIV/AIDS sentinel surveillance | HIV/AIDS prevalence and incidence rates. | Not specified | Not specified |
| **Jones 2008**[34] | Malaria surveillance | Feasibility of the system. | Increasing attitudes towards record-keeping and ownership of data. | Not specified |
| **Kagendi 2023**[35] | Not specified | Not specified | The paper reports the accuracy (78%), sensitivity (70%), specificity (83%), precision (68%), and F1 score (69%), indicating the effectiveness of the random forest model for predicting VL hotspots. | The partnership between Janssen and KEMRI demonstrates a commitment to capacity building in data science through knowledge transfer in research settings, while highlighting the potential of large datasets to simplify complex health situations and improve the understanding and management of HIV treatment. |
| **Kaneko 2012**[36] | Health and Demographic Surveillance System | Under-5 mortality rate; infant mortality rate; life expectancy; total fertility rate. | The ability to capture data for a significant number of residents (53,437 in Mbita) indicates some success in population surveillance efforts. | The need for comprehensive datasets and recognition of limitations in available data highlight the importance of robust data collection methods. |
| **Kapesa 2018**[37] | Malaria surveillance | Data collection efficiency; enhanced community engagement; timely | Not specified | The use of hospital malaria data in endemic setting as a |

| Study ID | AW&R system name | Key performance indicators | Success extent | Lessons learnt |
|---|---|---|---|---|
| | | identification of outbreaks; better resource allocation. | | reflection of the surrounding catchment area may be unreliable despite provision of free anti-malarial drugs at health facilities. |
| **Kariithi 2021**[38] | Newcastle surveillance | Not specified | Not specified | Not specified |
| **Kasolo 2013**[39] | Integrated Disease Surveillance and Response | Event detection and reporting. | Countries in the Horn of Africa, including Kenya, experienced severe drought resulting in famine, subsequent severe malnutrition, and population displacement into camps. An encamped population later experienced a cholera outbreak. | Not specified |
| **Katz 2014**[40] | National sentinel influenza surveillance | Not specified | Not specified | Effective surveillance, vaccination, and management strategies are needed to mitigate its impact on public health. |
| **Keshavamurthy 2021**[41] | Not specified | Early detection and warning of zoonotic disease events. | Most events were first reported by news media (78.2%) followed by international health organisations (16.4%). News media reported the events 4.1 days faster than the official reports. | Addressing the reporting gaps related to human health and expanding the scope of diseases monitored will be essential for achieving comprehensive surveillance and improving health outcomes. |
| **Kirimi 2017**[141] | RHAI surveillance | Not reported | Not reported | Not reported |

| Study ID | AW&R system name | Key performance indicators | Success extent | Lessons learnt |
|---|---|---|---|---|
| **Kirinyet 2016**[43] | Malaria surveillance | Not specified | Not specified | Training staff on outbreak response is essential for strengthening public health systems and enhancing readiness for future outbreaks. |
| **Kitala 2000**[44] | Rabies surveillance | Percentage of reported cases; forecasted rabies case numbers against actual occurrences. | The system identified a gross underestimation of the incidence of animal-and human-rabies. | The current passive surveillance program significantly underestimates the incidence of both animal and human rabies in Machakos County. |
| **Kyobutungi 2008**[45] | Urban Health and Demographic Surveillance System (Nairobi) | Estimation of years of life lost. | The NUHDSS effectively covers large parts of the Korogocho and Viwandani slums, gathering data on core demographic events such as births, deaths, in-migrations, and out-migrations. | Health planners could adopt a similar planning approach based on data from the NUHDSS to effectively benefit slum dwellers. |
| **Lickness 2020**[130] | Acute flaccid paralysis surveillance | Sensitivity of non-polio AFP; collection of adequate stool specimens from AFP patients; arrival of these specimens at a WHO-accredited laboratory by reverse cold chain; and ensuring that 80% of specimens are in good condition (without leakage or desiccation). | In 2018 Kenya, had 72.3% of the subnational areas had an NPAFP rate of greater or equal to 2 and greater or equal to 80% adequate specimens. | Environmental surveillance improves the sensitivity of poliovirus monitoring by detecting virus circulation that may occur without identified AFP cases, as seen in Cameroon, Côte d'Ivoire, and Kenya in 2018 and 2019 |
| **Lucinde 2024**[46] | SARS-CoV-2 Surveillance System | Mortality rates among SARI patients; vaccine effectiveness; testing coverage; access to specialised care. | Successfully identified at-risk populations, especially older patients and males, and provided actionable data to inform pharmaceutical and | Variability in documentation practices hindered the ability to comprehensively assess risk factors and co-morbidities, indicating a |

| Study ID | AW&R system name | Key performance indicators | Success extent | Lessons learnt |
|---|---|---|---|---|
| | | | non-pharmaceutical interventions. | pressing need for improved record-keeping systems to enhance data quality and patient management. |
| **Luka 2020**[47] | Health Demographic Surveillance System | Not specified | Not specified | Schools are a focal point in understanding human rhinovirus transmission in the community. |
| **Maes 2017**[131] | Acute flaccid paralysis surveillance | Number of non-polio AFP cases per 100,000 children under 15 years per year; adequate stool collection from ≥80% of AFP cases. | The number of reported WPV cases declined to its lowest point ever in 2016, as most national-level surveillance quality indicators improved. | Effective supervision and monitoring of Acute Flaccid Paralysis (AFP) surveillance are crucial for identifying, reporting, and investigating all actual AFP cases, especially as polio case counts decline; in this context, environmental surveillance serves as a vital supplement to enhance the detection of circulating viruses in high-risk areas with inadequate AFP surveillance. |
| **Maina 2017**[48] | District Health Information System version 2 | Percentage of facilities reporting any data for the whole 12-month period or for 3, 6, and 9 months. | Only 8% of public facilities had data reported for every month. | Not specified |
| **Maraka 2020**[49] | Malaria surveillance | Not specified | Not specified | Understanding patterns of gene flow and resistance genotypes is essential for developing targeted malaria control strategies in Kenya. |
| **Matete 2003**[50] | Integrated surveillance system for zoonotic diseases | Prevalence of diseases; time to detection; time to response. | Successful identification of 200 dogs, of which 5 were positive for trypanosomes, | Not specified |

| Study ID | AW&R system name | Key performance indicators | Success extent | Lessons learnt |
|---|---|---|---|---|
| | | | resulting in a prevalence of 2.5%. | |
| **Mathenge 2018**[142] | Geo-database | Not reported | The system-aided visualisation reported TB cases linked to specific health facilities through maps, charts and graphs | Not reported |
| **Milliano 2022**[143] | Integrated Disease Surveillance and Response | Level of utilisation of IDSR data; proportion of trained healthcare workers; level of knowledge of the IDSR data. | Not specified | Not specified |
| **Montana 2008**[51] | Antenatal care sentinel surveillance | HIV/AIDS prevalence. | Not specified | Assessing HIV prevalence estimates for bias is necessary, and we recommend using population-based survey data to calibrate clinic-based surveillance estimates. |
| **Morobe 2019**[144] | Kilifi Health and Demographic Surveillance System (KHDSS) | Not reported | Not reported | Not reported |
| **Moturi 2014**[52] | Mobile based Disease Surveillance Prototype System | Not specified | Not specified | Not specified |
| **Mtetwa 2024**[53] | Wastewater based epidemiology | Correlation coefficient; limit of detection for TB drugs; limit of quantification for TB drugs; matrix effects; stability of TB drugs during storage; and detection levels of TB drugs in treated wastewater. | The system successfully detected multiple TB drugs, including ethambutol, erythromycin, pyrazinamide, and isoniazid, in both untreated and treated wastewater. | Accounting for matrix effects in wastewater surveillance is crucial for ensuring accurate measurements and reliable data interpretation. |

| Study ID | AW&R system name | Key performance indicators | Success extent | Lessons learnt |
|---|---|---|---|---|
| **Munywoki 2023**[54] | Population Based Infectious Disease Surveillance | Not specified | Not specified | Not specified |
| **Muriithi 2022**[145] | Brucellosis Surveillance | Not reported | Not reported | Not reported |
| **Murray 2013**[55] | Hospital based influenza surveillance | Sensitivity and specificity of influenza surveillance. | Not specified | To determine disease burden and maximize sensitivity in surveillance, combining cough with any fever (either subjective or measured at 38°C) can provide a simple, sensitive case definition for hospitalized laboratory-confirmed influenza across all age groups. |
| **Mwatondo 2016**[56] | Integrated Disease Surveillance and Response | Completeness of reports; timeliness of report submission. | 87 % of the weekly forms were filled to completeness while timeliness of the reports was at 93%. | Adequate reporting in Nairobi County is promoted by having weekly reporting forms available, a dedicated surveillance focal person, and posters displaying Integrated Disease Surveillance and Response (IDSR) functions in facilities. |
| **Namuye 2015**[57] | Early Warning Alert and Response System | Processed and visualised data. | Not specified | Different geographical areas require tailored information on diseases, but the system faces challenges such as high internet costs, poor infrastructure, lack of awareness, and low literacy levels in rural areas. |

| Study ID | AW&R system name | Key performance indicators | Success extent | Lessons learnt |
|---|---|---|---|---|
| **Ndegwa 2023**[58] | Mobile event-based surveillance system | Established reporting system. | From June 2020 to June 2021, 8734 signals were reported through CEBS, with 2961 (33.9%) verified as events. | The existing Early Warning Surveillance (EBS) structure enabled rapid adaptation for detecting COVID-19 cases during the pandemic, while both Community Early Warning Surveillance (CEBS) and Health Early Warning Surveillance (HEBS) continued to report non-COVID-19-related events; the CEBS system consistently identified COVID-19 signals even amid healthcare worker strikes, underscoring the vital role of community structures in early detection. |
| **Nderu 2019**[60] | Malaria surveillance | Total number of isolates; mean number of alleles; allelic richness; expected heterozygosity; polyclonal infections; multilocal linkage disequilibrium. | Not specified | Not specified |
| **Ng'eno 2023**[60] | Population Based Infectious Disease Surveillance | Not specified | A reduction in turnaround times for laboratory results by 30%. | Not specified |
| **Ng'etich 2021**[63] | Integrated Disease Surveillance and Response | Simplicity; acceptability; stability; flexibility; usefulness; data quality; surveillance data reporting timeliness; data completeness. | 55% of the respondents (73/134) perceived the surveillance system to be simple, 50% (83/165) to be acceptable, 41% (66/162) to be stable, 41% (42/102) to be flexible, 51% (52/103) to be | Health workers' perceptions on the surveillance systems' attributes influences the overall performance of other surveillance core and support functions. |

| Study ID | AW&R system name | Key performance indicators | Success extent | Lessons learnt |
|---|---|---|---|---|
| | | | useful and 25% (41/165) indicated that the surveillance system provided quality PC-NTDs surveillance data. | |
| **Ng'etich 2021**[64] | Integrated Disease Surveillance and Response | Capacity to detect, confirm, report, analyse, and interpret surveillance data to inform response actions. | At the sub-national level, 84% of sub-counties had the capacity to collect and store PC-NTD specimens. | Targeted interventions are essential to enhance surveillance effectiveness. |
| **Ng'etich 2021**[61] | Preventive Chemotherapy for Neglected Tropical Diseases | Perception of simplicity; perception of acceptability; perception of stability; perception of flexibility; perception of usefulness; perception of data quality; reporting timeliness; reporting completeness; trends in reporting timeliness and completeness. | Health workers expressed low satisfaction with their involvement in facility-based surveillance activities and perceived PC-NTDs as a low priority compared to other health conditions. | The transition from paper-based to electronic systems must be managed carefully to avoid disrupting reporting mechanisms and ensure that health workers can easily adapt to new technologies. |
| **Ng'etich 2021**[62] | Preventive Chemotherapy for Neglected Tropical Diseases | Feasibility of the system. | Not specified | Limited data can hinder the processes of monitoring and evaluation, making it difficult to determine what works and what doesn't. |
| **Ngugi 2018**[65] | Rabies surveillance | Number of patients receiving rabies vaccine; incidence of animal bites | Passive surveillance identified high risk groups for animal-bite, culpable animals that are most likely to cause bites and exposure factors associated with animal-bite. | Passive surveillance often leads to incomplete data due to underreporting, inconsistencies in data collection, and reliance on healthcare providers to voluntarily submit reports |
| **Ngugi 2020**[132] | Community Health Surveillance System (Kaloleni/Rabai) | Household participation rate. | This system found that 19% of the respondents reported an illness in the preceding month, of whom 77% sought health care in a health facility. | Not specified |

| Study ID | AW&R system name | Key performance indicators | Success extent | Lessons learnt |
|---|---|---|---|---|
| **Njenga 2021**[133] | Kenya Animal Bio surveillance System | Time to report submission; System geographical coverage. | As of December 2019, 11,399 domestic and 205 wild animal disease reports were received through KABS. | The immediate submission of reports and automated collection of geographic coordinates in the AW&R system significantly improved the timeliness of data reporting and enhanced follow-up by health officers in Kenya. |
| **Njeru 2020**[66] | District Health Information System version 2 | Completeness; timeliness of reporting | Demonstrated impact of training in reporting indicators. | Not specified |
| **Njuguna 2013**[67] | Population Based Infectious Disease Surveillance | Not specified | It revealed a high burden of shigellosis within a densely populated urban slum in Kenya. | Not specified |
| **Njuguna 2014**[68] | Influenza virus surveillance | Data completeness and logical consistency; data timeliness; start-up costs; year 1 operational costs; two-year operational costs | Electronic data collection was employed by the system to improve timeliness, data integrity and reduce costs. | Not specified |
| **Njuguna 2019**[69] | Health and Demographic Surveillance System (Kilifi) | Not specified | Not specified | Not specified |
| **Nokes 2009**[70] | Hospital based RSV surveillance | Not specified | Demonstrated the power of long-term surveillance in understanding the role of RSV in morbidity and mortality patterns in children in a low resource setting, and the requirement for population evidence in developing a | Not specified |

| Study ID | AW&R system name | Key performance indicators | Success extent | Lessons learnt |
|---|---|---|---|---|
| | | | rational strategy for future RSV vaccine intervention. | |
| **Nyagah 2019**[71] | Mortuary based surveillance | Prevalence of HIV/AIDS-related deaths. | It identified that females continue to bear the greatest burden of HIV infection. | Not specified |
| **Nyawanda 2023**[72] | Population Based Infectious Disease Surveillance | Incidence rate of malaria; demographic breakdown of malaria cases; seasonal patterns of malaria incidence; climatic factors (LSTD and rainfall); bed net usage; correlations between climatic factors and malaria incidence; model-based estimates of factors influencing malaria incidence; and projected trends for future malaria incidence. | The surveillance has been successful to a significant extent, as indicated by the ability to monitor and analyse trends in malaria incidence, seasonal patterns, and the impacts of climatic factors on malaria transmission, which informs public health interventions and resource allocation. | Understanding the relationship between climatic factors and malaria incidence is essential for developing effective surveillance and response strategies. |
| **Ochieng 2013**[73] | Entomologic arbovirus surveillance | Not specified | Not specified | Not specified |
| **Odari 2014**[74] | Cholera Surveillance | Not reported | Not reported | Not reported |
| **Odhiambo 2012**[134] | KEMRI/CDC Health and Demographic Surveillance System | Not specified | It led to the discovery that bed nets reduce community infant mortality by 26%,3 and that over time, bed net use does not reverse gains in mortality reduction in young children. | Not specified |
| **Odhiambo 2015**[75] | Integrated Response System for Emerging Infectious Diseases | Not specified | Not specified | Not specified |

| Study ID | AW&R system name | Key performance indicators | Success extent | Lessons learnt |
|---|---|---|---|---|
| **Oduor 2023**[76] | Population Based Infectious Disease Surveillance | Not specified | Not specified | Not specified |
| **Oduor 2023**[77] | Population Based Infectious Disease Surveillance | Not specified | Over the 10-year study period, the system observed an 88% decline in the rate of pneumonia/ARI deaths among children aged <5 year. | Not specified |
| **Ogira 2022**[78] | Not specified | Not specified | Not specified | Effective pandemic preparedness requires improved investments across all health system building blocks. |
| **Olack 2010**[79] | Population Based Infectious Disease Surveillance | Not specified | Not specified | Not specified |
| **Olotu 2010**[80] | Health Demographic Surveillance System | Sensitivity and specificity of outbreak detection and incidence. | Not specified | Passive surveillance without specific prompts for visits detected only about one-third of the cases that active surveillance would have identified. |
| **Omemo 2012**[81] | Not specified | Not specified | Not specified | While rabies is the most frequently encountered zoonotic disease among respondents (56%), nearly all (98%) engage their communities in discussions about zoonotic diseases. |
| **Omondi 2019**[146] | Integrated Disease Surveillance and Response | Not specified | 83% of respondents used the IDSR every week, and 12% used it daily | Not specified |

| Study ID | AW&R system name | Key performance indicators | Success extent | Lessons learnt |
|---|---|---|---|---|
| **Omondi 2020**[82] | Integrated Disease Surveillance and Response | Availability of IDSR system tools; timely dissemination of IDSR data; uptake of IDSR data in community health facilities. | Not specified | Training community health workers (CHWs) is crucial for enhancing IDSR data utilization. |
| **Omore 2024**[135] | Enterics for Global Health Shigella surveillance | Not specified | Not specified | Not specified |
| **Onyango 2012**[83] | Health and Demographic Surveillance System (Kilifi) | Not specified | Not specified | Not specified |
| **Opiyo 2017**[147] | Kilifi Health and Demographic Surveillance System (KHDSS) | Not reported | Not reported | Not reported |
| **Otambo 2023**[84] | Malaria surveillance | Early detection. | The CHVs correctly classified malaria symptoms, used the active case-detection chart. | The number of health trainings received by community health volunteers (CHVs) was significantly associated with their correct use of job aids and safety procedures during active case detection (ACD) activities. |
| **Otieno 2019**[85] | Flood impact-based forecasting system | Flood events count; number of houses destroyed; flood extent maps; observed rainfall during flood events; community vulnerability assessment; flood risk scores. | By identifying vulnerable populations and areas at risk, flood impact-based forecasting (FIBF) enabled more effective and targeted responses from government agencies and humanitarian organizations. | Not specified |

| Study ID | AW&R system name | Key performance indicators | Success extent | Lessons learnt |
|---|---|---|---|---|
| **Otieno 2022**[148] | Ecological Niche Model (ENM). | Not reported | Not reported | Not reported |
| **Ototo 2015**[86] | Not specified | Average number of female mosquitoes per house per night; number and type of mosquitoes collected using rotator traps (indoors and outdoors); proportion of the population staying outdoors during peak mosquito biting times (evening and dawn). | The rotator traps collected mosquitoes every 2 hours, revealing that 68% of An. funfests collected were mostly blood fed, and 58% of An. gambiae vectors collected were not fed. | Not specified |
| **Owor 2016**[87] | Not specified | HMPV prevalence in child admissions; temporal occurrence and circulation patterns of HMPV; genetic diversity of HMPV samples. | HMPV was detected in 4.8 % (160/3320) of children [73.8 % (118/160) of these less than one year of age] ranging between 2.9 and 8.8 % each year over the 5 years of study. HMPV infections were seasonal in occurrence, with cases predominant in the months of November through April. These months frequently coincided with low rainfall, high temperature and low relative humidity in the location. | Not specified |
| **Owuor 2022**[88] | Health facility-based surveillance networks | Not specified | Not specified | Not specified |
| **Owuor 2023**[89] | Child Health and Mortality Prevention Surveillance Network | Prompt immediate public health actions. | Provided an insight into gaps in the diagnosis and management of RDS, coupled with increasing knowledge of lung tissue histology, and | Not specified |

| Study ID | AW&R system name | Key performance indicators | Success extent | Lessons learnt |
|---|---|---|---|---|
| | | | highlighted opportunities for clinical care improvement. | |
| **Owusu 2024**[90] | Influenza and SARS-CoV-2 sentinel surveillance | Outbreak detection. | The system detected influenza (A or B) in 8.7% (n=629); 7.4% (n=103) tested positive among outpatients and 9% (n=526) among inpatients. | The sentinel system effectively detected increased SARS-CoV-2 activity during the study period. |
| **Oyas 2018**[91] | Rift Valley Fever surveillance | Early detection. | RVF enhanced surveillance pilot demonstrated the capacity and need for establishing a national syndromic surveillance system in livestock in Kenya. | Not specified |
| **Park 2012**[92] | Modified TB treatment surveillance programme | Not specified | The modified TB retreatment surveillance programme at AMPATH MRL resulted in a larger proportion of retreatment patients accessing DST service within the catchment area. | Identifying existing health system resources, such as the expertise of regional laboratory staff in training and access, the AMPATH transport system, and the DLTLD's supply chain with the MRL's inventory monitoring, can support national or provincial-level programs. |
| **Pedro 2016**[93] | Rift Valley Fever Monitoring and Risk Mapping (RVFMRM) | Not reported | Not reported | Not reported |
| **Peletz 2018**[94] | Water quality monitoring system | Microbial water quality testing performance; water capacity rating diagnostic scores. | Not specified | Not specified |
| **Pilgrim 2018**[95] | Early Warning Indicator Surveillance | On-time pill pick-up; retention in care; virological suppression | The monitoring of EWIs has not been routine in health facilities in Kenya. | Establishing a robust monitoring and evaluation framework is essential to |

| Study ID | AW&R system name | Key performance indicators | Success extent | Lessons learnt |
|---|---|---|---|---|
| | | | | assess the effectiveness of the implemented strategies and make data-driven adjustments as needed. |
| **Post 2020**[96] | SARS-CoV-2 Surveillance System | Seven-day persistence indicate rates of COVID-19 transmissions. | The system recorded a seven-day persistence effect on speed (new cases per day per 100,000), n = 2150. | While standard surveillance metrics like daily reported COVID-19 cases and deaths are essential, they are insufficient alone to effectively mitigate and prevent the transmission of the virus. |
| **RinkedeWit 2022**[97] | Mobile event-based surveillance system | Tailored quality improvement plan focused on COVID-19; follow up potential COVID-19 cases. | Adherence to the continuum of maternal care was not disrupted among Mom Care clients. In Kenya by mid-October 2020 only 800 users had subscribed to the service, 650 active users on the Covid Connect platform, 195 were high risk users and 2,018 patient alerts were processed by Care Coordination Centre. | In the later phases of the pandemic, the renewed use of the SafeCare4COVID App remained relatively limited, while the implementation of COVID-Dx was notably successful, advancing toward a broader digital epidemic preparedness and monitoring approach; its local adoption was facilitated by its co-design with local stakeholders, making it user-friendly and encouraging for users to complete. |
| **Ruttoh 2023**[98] | SARS-CoV-2 Surveillance System | Outbreak detection. | SARS-CoV-2 was detected in 177 study participants, indicating a prevalence rate of 13.9%, with higher rates observed among females | There is a need for syndromic testing for acute respiratory infections (ARIs) to enhance clinical decision-making, while sustained surveillance efforts |

| Study ID | AW&R system name | Key performance indicators | Success extent | Lessons learnt |
|---|---|---|---|---|
| | | | (15.5%, n = 118) compared to males (11.6%, n = 59). | are essential to monitor disease trends and guide public health actions. |
| **Sanders 1998**[100] | Rift Valley Fever surveillance | Number of cases identified; overall mortality rate among identified cases. | 55 persons with haemorrhagic fever were identified from September 1992 through March 1993. | Not specified |
| **Scobie 2017**[100] | Integrated Disease Surveillance and Response | Tetanus sero-protection rate (< 70%); median antibody concentration (≤ 0.05 IU/mL). | Identified that tetanus sero-protection was lower among children 5-14 years versus 1-4 years of age in Kenya (66% versus 90%) and among males than females. | Not specified |
| **Scott 2012**[136] | Health and Demographic Surveillance System (Kilifi) | Not specified | Identified the incidence of vaccine preventable diseases caused by S. pneumoniae and H. influenzae was 111 and 60/100 000/year, respectively. We used the KHDSS to examine the interactions between significant infectious pathogens demonstrating, for example, that over half of all invasive bacterial disease was attributable to prior exposure to P. falciparum infection. | Not specified |
| **Seheri 2018**[101] | World Health Organization's rotavirus surveillance | Not specified | Not specified | There is no evidence of increased selection pressure on specific genotypes that would compromise vaccine effectiveness. |

| Study ID | AW&R system name | Key performance indicators | Success extent | Lessons learnt |
|---|---|---|---|---|
| **Sewe 2016**[102] | Malaria surveillance | Total malaria deaths; deaths by area; mean weekly deaths; peak death counts; mean day land surface temperature; risk of mortality; overall relative risk of malaria mortality associated with precipitation. | The EWS was effective in proactively monitoring environmental variables such as land surface temperature (LST), precipitation, and vegetation index (NDVI), which were associated with malaria mortality. | Early warning systems (EWS) should be designed to be flexible and responsive to changing environmental conditions, such as shifts in climate or land use, which can influence disease dynamics. |
| **Sewe 2017**[103] | Malaria surveillance | Prediction accuracy ($R^2$ values); root mean squared error; mean absolute error. | The high predictive accuracy and performance of the 1-month lead GAMBOOST model. | Not specified |
| **Sharma 2015**[104] | Treatment Information from Basic Unit | Completeness of the data. | The concordance between the registers and TIBU was better than that between TB patient cards and TIBU, although the district registration number and the registration date had kappa scores below 0.40. | Enhancing TB surveillance data quality in Kenya requires additional oversight and training in data entry. |
| **Shih 2022**[105] | Acute febrile illness surveillance | Enrolment rates; age distribution of enrolled patients; testing coverage for SARS-CoV-2; positivity rates of SARS-CoV-2 among tested patients; sex distribution of enrolled patients; geographic coverage of surveillance sites; integration timeline for COVID-19 testing into AFI surveillance systems. | The detection of SARS-CoV-2 in patients with AFI indicates that the surveillance system successfully identified positive cases. | Not specified |
| **Sifuna 2014**[106] | Health and Demographic Surveillance System (Kombewa) | Not specified | The Kombewa HDSS has successfully supported recruitment and retention activities for several research | Not specified |

| Study ID | AW&R system name | Key performance indicators | Success extent | Lessons learnt |
|---|---|---|---|---|
| | | | studies nested within it, conducted at the Kombewa Clinical Research Centre, including the phase3 randomized, controlled trial of RTS, S/AS01 malaria vaccine candidate and the associated Malaria Transmission Intensity study. | |
| **Sirengo 2016**[107] | Routine HIV/AIDS testing surveillance | Infection detection. | The proportion of women recorded in the ANC registry sample who had a HIV-positive result from routine antenatal HIV testing was 6.5 % and among those surveyed in the ANC SS sample was 6.2 %. | The low positive percent agreement between routine and UAT HIV test results, along with poor PT performance, indicates a need for routine evaluations of HIV testing data quality at ANC testing sites and a review of central laboratory testing data for HIV surveillance before discontinuing ANC sentinel surveillance. |
| **Soti 2015**[108] | Mobile event-based surveillance system | Data capture, transmission and aggregation, timeliness of data capture and transmission. | A total of 5812 malaria RDTs were run and uploaded to the cloud database during the programme. | This paperless, mHealth innovation brought about unprecedented quality control and quality assurance in diagnosis of malaria, patient care and data capture, all in the hands of the health worker at point of care in an integrated way. |
| **Soura 2015**[109] | Urban Health and Demographic | Not specified | Not specified | Not specified |

| Study ID | AW&R system name | Key performance indicators | Success extent | Lessons learnt |
|---|---|---|---|---|
| | Surveillance Systems (Nairobi) | | | |
| **Stehling-Ariza 2023**[137] | Acute flaccid paralysis surveillance | Sensitivity and specificity of AFP surveillance. | Not specified | Continuous surveillance allows for the early identification of disease outbreaks, enabling timely public health responses to contain and control the spread of infections. |
| **Swierczewski 2013**[110] | Enteric Disease Surveillance | Frequency of pathogen detection. | Not specified | Future investigations should establish additional surveillance sites in the rural areas of Kericho and Kisumu to include individuals who lack access to district hospitals. |
| **Taylor 2020**[111] | Child Health and Mortality Prevention Surveillance Network | Not specified | Not specified | Not specified |
| **Tesfaye 2020**[157] | Acute flaccid paralysis surveillance | Sensitivity and specificity of AFP surveillance. | Not specified | Kenya possesses a sensitive and high-quality AFP surveillance system capable of effectively detecting and reporting poliovirus cases. |
| **Thomas 2021**[113] | Cross-sectoral zoonotic disease surveillance | Cross-sectoral collaboration. | Not specified | Political will not only facilitates resource allocation and operational structures but also shapes common objectives, creating a sequential interaction where each theme reinforces the |

| Study ID | AW&R system name | Key performance indicators | Success extent | Lessons learnt |
|---|---|---|---|---|
| | | | | next, enabling sectors to align their actions effectively. |
| **Thumbi 2015**[114] | Population-based animal syndromic surveillance; Population-based infectious disease surveillance | Described epidemiologic patterns of disease; aetiology of diseases; implemented health impact of interventions in the study population. | In cattle, goats and sheep, accounting for 56% of all livestock syndromes, followed by respiratory illnesses (18%) while in humans, respiratory illnesses accounted for 54% of all illnesses reported, followed by acute febrile illnesses (40%) and diarrhea illnesses (5%). | Antenatal care sentinel surveillance can strengthen PMTCT programs by integrating PMTCT variables into ANC data collection forms or by comparing both data sets for effective monitoring and evaluation. |
| **Thumbi 2019**[115] | Not specified | Real-time surveillance data of infectious disease. | A total of 20340 animal and death events were reported from the community through the two surveillance systems, half of which were confirmed as valid disease events. | Mobile phone-based animal health surveillance system effectively empowers rural African communities to report disease events directly, bypassing the need for healthcare or veterinary workers. |
| **Toda 2016**[116] | Mobile short messaging service-based disease outbreak alert system | Cases detected; notifications submitted; responses undertaken. | During the 6-month period after the intervention, 169 immediately notifiable cases (130 for the intervention group, 39 for the control group) were detected: 160 measles, 6 anthrax, 2 Q fever, and 1 guinea worm. | While the SMS intervention significantly improved timely notifications, response rates remained suboptimal, with only one-fifth of detected cases being reported; IDSR refresher training likely enhanced case detection, and the combination of interventions, including technology, yielded better results, yet weaknesses such |

| Study ID | AW&R system name | Key performance indicators | Success extent | Lessons learnt |
|---|---|---|---|---|
| | | | | as insufficient on-the-job training for staff who missed training, and a lack of post-training support hindered the overall effectiveness. |
| **Toda 2017**[117] | Integrated Disease Surveillance and Response | Outbreak detection. | Users thought that the system was reliable because it reduced the need for multiple officers entering and sending the data up the chain. | While innovative mobile technology for disease surveillance is widely accepted and appreciated for its simplicity, addressing broader health system challenges—such as post-training follow-up and supportive supervision—is crucial for successful national rollout. |
| **Toda 2018**[118] | Mobile short messaging service-based disease outbreak alert system | Not specified | Not specified | Not specified |
| **Umuhoza 2020**[119] | Influenza virus surveillance | Not specified | Contributed to responding to influenza outbreaks that occurred in Kenya and the East African region during the study period. | Not specified |
| **VanHemelrijck 2009**[120] | Longitudinal health facility surveillance | Not specified | Not specified | Not specified |
| **VanZyl 2019**[158] | Bag-mediated filtration environmental surveillance | Detection of poliovirus from environmental waters. | Diverse enteric viruses were frequently detected in BMFS samples, with detection in 88%-100% of samples, depending on the virus. | The consistent detection of enteroviruses indicates their potential as indicators in forcibly contaminated sites |

| Study ID | AW&R system name | Key performance indicators | Success extent | Lessons learnt |
|---|---|---|---|---|
| | | | | and as process controls in the BMFS. |
| **Walker 2014**[122] | Acute flaccid paralysis surveillance | Effectiveness of AFP surveillance in detecting and responding to acute flaccid paralysis and monitoring poliovirus transmission. | Not specified | Deficiencies were reported at multiple levels of the health system, primarily related to funding, training, and supervision. Notable shortcomings in surveillance and routine immunization activities were particularly evident at the local level. While AFP surveillance met national performance standards, widespread deficiencies and limited resources were observed at subnational levels. |
| **Wathondu 2016** | Malaria surveillance | Not reported | Not reported | Not reported |
| **Worsley-Tonks 2022**[123] | Kenya Animal Bio surveillance System | Sensitivity and specificity of zoonotic disease surveillance. | Not specified | Establishing zoonotic disease units, such as Kenya's Zoonotic Disease Unit (ZDU), would enhance One Health training and foster cross-sector collaborations at national and subnational levels, with targeted efforts needed for remote rural areas. |
| **Zhou 2015**[124] | Malaria surveillance | Not specified | Not specified | Various surveillance methods yield distinct epidemiological profiles. |

| Study ID | AW&R system name | Key performance indicators | Success extent | Lessons learnt |
|---|---|---|---|---|
| **Zhou 2020**[125] | Bag-mediated filtration environmental surveillance | Polio virus detection. | Polioviruses were detected in most samples using BMFS (37/42) and two-phase separation (32/42). | BMFS can be used for PV environmental surveillance. |

**CLIMATE-SENSITIVE INFECTIOUS DISEASES**

**State of Climate Kenya**

**Table 5.1: State of Climate Kenya Summaries**

| Year | Environmental Condition | Climate-sensitive infectious diseases |
|---|---|---|
| **2022**[150] | **Monthly Temperature Distribution:**<br>• Hottest month: March (average 23.3°C)<br>• Coolest months: August (average 19.7°C) and July (average 19.8°C)<br>• 2022 was warmer than normal, with significant increases in Mandera, Lodwar, Kakamega, and coastal stations<br>• 2022 was the 3rd warmest year on record<br><br>**Monthly Rainfall Distribution:**<br>• Annual Overview: Below-average rainfall in most parts of the country<br>• March to May (MAM): Depressed rainfall, with exceptions in Moyale, Kisumu, Voi, and Laikipia in March; Wajir and Nakuru in May<br>• June to August (JJA): Dry conditions in most parts, except for near-average to above-average rainfall in the western sector, coastal region, and Highlands East of the Rift Valley<br>• October to December (OND): Depressed rainfall, with delayed onset in some areas and a false onset in others, leading to poor distribution<br><br>**Flood Events** | • 1,409 suspected Cholera cases reported nationwide between December 5th and 18th<br>• This contributed to 2,959 confirmed cases between October and December 2022<br>• County distribution of Suspected Cholera Cases:<br> ◦ Garissa County: 654 cases<br> ◦ Nairobi County: 384 cases<br> ◦ Machakos County: 231 cases<br> ◦ Kiambu County: 164 cases |

| | | |
|---|---|---|
| | Some isolated flooding incidents were reported<br>• Marsabit, January 17th: Received heavy rainfall (89.9mm)<br>• Kisumu, September: Heavy rainfall caused rivers draining into Lake Victoria to burst their banks<br>• Taveta, November 16th-18th: Heavy rains within the national park caused flash floods in the neighbouring Mata Ward<br><br>**Seasonal Monsoon Winds:**<br>• North East Monsoon (DJF): Low wind speeds along EEA coast, high speeds along Somali coast<br>• South East Monsoon (JJA): High wind speeds throughout the WIO basin, rough seas with winds >17 knots and waves >2 meters<br><br>**Drought**<br>• The prevailing extreme event of 2022 was drought, extending from 2021<br>• The drought situation remained critical in 22 of the 23 arid and semi-arid (ASAL) counties due to the late onset and poor performance of October to December 2022 short rains, coupled with four previous consecutive failed rainfall seasons<br>○ Counties affected;<br>▪ Alarm drought phase: Kilifi, Mandera, Marsabit, Samburu, Turkana, Wajir, Isiolo, Kitui, and Kajiado<br>▪ Alert drought phase: Garissa, Lamu, Narok, Tana River, Makueni, Tharaka Nithi, Baringo, Laikipia, Meru, Taita Taveta, West Pokot, Nyeri, and Kwale<br>▪ Normal drought phase: Embu | |
| **2023**[151] | **Temperature Trends**<br>• 2023 was the warmest year globally<br>• Kenya experienced a clear warming trend, with average maximum temperatures rising from 29°C in the late 1970s to 30.5°C in the early 2020s, an increase of approximately 1.5°C<br>• Monthly temperatures in 2023 showed a pattern of above-normal temperatures in most parts of Kenya, though some areas like the Central Highlands and parts of Nairobi and the Southeastern lowlands experienced cooler conditions | • Cholera outbreak affected 27 counties: Nairobi, Kiambu, Nakuru, Uasin Gishu, Kajiado, Murang'a, Machakos, Garissa, Meru, Nyeri, Wajir, Tana River, Kitui, Homa Bay, Mandera, West Pokot, Bomet, Samburu, Marsabit, Kirinyaga, |

| | | |
|---|---|---|
| | **Rainfall Patterns**<br>• Overview: Kenya experiences significant rainfall variability due to factors such as altitude, the Indian Ocean Dipole, and El Niño-Southern Oscillation (ENSO)<br>• MAM season: Most regions experienced near to above-average rainfall, with southwestern areas and the Lake Victoria Basin receiving significant rainfall. Coastal areas like Taita Taveta and Kwale had a late onset of rains<br>• JJA season: Characterized by dry conditions in most areas, though rainfall was recorded in specific regions like the Highlands West of the Rift Valley and the Central and South Rift Valley<br>• OND season: Varied rainfall distribution with severe storms in November, particularly in the coastal region, southeastern lowlands, and northeastern areas<br><br>**Extreme Weather Events**<br>• Drought: The year began with drought due to five consecutive failed rainy seasons since 2020.<br>• Flooding: Heavy rainfall in 2023 led to extreme flooding, particularly in the coastal and low-lying regions. Flash events were recorded in March, April, and October-November<br><br>**Ocean and Marine Weather**<br>• Cyclones: Tropical cyclones, such as Cyclone Freddy, impacted the region with no major direct impacts on Kenya but contributed to extreme rainfall and storms<br>• Sea-Level Rise: The Western Indian Ocean region, including Kenya, experienced higher sea-level rise rates compared to the global average, contributing to coastal erosion and salinisation | Kisumu, Siaya, Isiolo, Mombasa, Kwale, Migori and Busia<br>• As of 2nd October 2023, a total of 12,123, vector-borne disease cases, with 627 confirmed by culture, and 202 deaths were reported |

**Kenya Meteorological Department- climate outlook for the June-July-August (JJA) 2022 season and rainfall performance during the March-April-May (MAM) 2022 “long rains” season**

**Table 5.2: Kenya Meteorological Department, 2022 Report**

| Season | Climatic Condition | Climate-sensitive infectious disease |
|---|---|---|
| **MAM performance 2022**[152] | **Rainfall Performance**<br>• Generally poor distribution of rainfall over most parts of the country<br>• Delayed onset: Expected rainfall delayed in the Highlands West of the Rift Valley, Coastal region, and parts of the northwest<br>• Below-average rainfall was recorded in most regions, with notable exceptions in Eldoret, Kisumu, Nakuru, Moyale, and Narok<br>• Depressed rainfall (<75% of long-term averages) was reported in most regions, including parts of the northeastern and ASAL areas<br>• Kisii recorded the highest rainfall (529mm), followed by Kisumu (514.3mm)<br>• Lowest rainfall: Lodwar recorded only 7.5mm for the entire season | No disease outbreaks directly related to weather conditions. |
| **Outlook for JJA 2022** | **Rainfall Forecast**<br>• Highlands West of Rift Valley, Lake Victoria Basin, Central Rift Valley, Northwestern region, and parts of Central Kenya are expected to receive slightly above-average rainfall<br>• Coastal Strip (Mombasa, Tana River, Kilifi, Lamu, Kwale) is likely to experience below-average rainfall<br>• Highlands East of Rift Valley (Nairobi, Nyandarua, Kiambu, Meru, etc.) are expected to be cool and cloudy with occasional drizzle/light rains<br>• The North-Eastern Region and parts of Northwestern Kenya is expected to be generally sunny and dry conditions<br><br>**Temperature Forecast**<br>• Highlands East of Rift Valley is expected to be cool and cloudy, with maximum temperatures occasionally falling below 18°C<br>• Northeastern Region (Marsabit, Isiolo, Mandera, etc.): Sunny and dry with near-normal temperatures<br>• Highlands West of Rift Valley, Lake Victoria Basin, and Central Rift Valley are expected to be slightly cooler than usual temperatures<br>• Coastal Region, Southeastern Lowlands, and parts of Northeastern Kenya are expected to be warmer than usual temperatures | • Respiratory diseases (asthma, pneumonia, flu) expected to increase in cold regions (Nairobi, Highlands East and West of Rift Valley)<br>• Malaria cases may rise, but not beyond normal transmission due to reduced temperatures<br>• Waterborne diseases may increase due to contamination of water sources |

**MALARIA EPIDEMIC EARLY PREDICTION SYSTEM FOR WESTERN KENYA HIGHLAND FOR FEBRUARY 2021**

**Table 5.3: Malaria Epidemic Early Prediction System for Western Kenya Highland for February 2021**

| Region | Climatic condition | Infectious diseases |
|---|---|---|
| **Kakamega** [153] | • Maximum temperature decreased slightly, with a Nil anomaly in January 2021 (compared to the monthly mean)<br>• Rainfall decreased from 108.1mm in December to 104.2mm in January | The model output indicates a low risk of malaria outbreak, with an additive percentage risk of 18.2%, below the epidemic threshold of 30% |
| **Kisii** | • Slight decrease in maximum temperature (26.1°C in Dec 2020 to 26.0°C in Jan 2021)<br>• Rainfall decreased from 178.4mm in December to 73.8mm in January | The model output indicates a Nil risk for the malaria epidemic, as the risk is below the threshold level of 20% |
| **Nandi** | • Slight decrease in maximum temperature (25.1°C in Dec 2020 to 25.0°C in Jan 2021)<br>• Significant increase in rainfall from 70mm in December to 187.7mm in January | The model output indicates a Nil risk for the malaria epidemic, with no threat based on the model's multiplicative calculation |

**National Drought Early Warning Bulletin**

Table 4.4 below summaries the findings from the National Drought Early Warning Bulletins presented by the National Drought Management Authority[154]. Out of 83 bulletins, only 14 reported data on climate-sensitive infectious diseases. The table below summarises the 14 months between April 2023 and September 2024.

**Table 5.4: Bulletin Summaries**

| Bulletin month, year | Climatic Condition | Climate-sensitive infectious diseases reported |
|---|---|---|
| **April 2023** | **Flash floods** were reported in Marsabit, Mandera, Garissa, Isiolo, Samburu, Narok, Taita Taveta, Tana River and Wajir Counties<br><br>**Rainfall Distribution**: | **Diseases Reported**: Contagious Caprine Pleuropneumonia (CCPP), Contagious Bovine Pleuropneumonia (CBPP), Pest Petis Ruminantes (PPR), Foot and Mouth Disease |

| | | |
|---|---|---|
| | • <u>Pastoral North East counties</u> (Mandera, Wajir, Isiolo, Tana River, Garissa) received <1mm to 25mm of rainfall<br>• <u>Pastoral North West counties</u> (Turkana, Marsabit, Samburu) received <1mm to 50mm.<br>• <u>South East Marginal Agriculture counties</u> (Tharaka Nithi, Embu, Kajiado, Meru, Makueni, Kitui) received <1mm to 25mm<br>• <u>Agro Pastoral cluster counties</u> (Kajiado, Laikipia, Narok, Baringo, Nyeri, West Pokot) received <1mm to 50mm<br>• <u>Coast Marginal Agriculture counties</u> (Kwale, Kilifi, Lamu) received 26mm to 100mm<br><br>**Vegetation Condition;**<br>• <u>Extreme Deficit:</u> Mandera, Tana River<br>• <u>Severe Deficit</u>: Marsabit, Wajir, Isiolo<br>• <u>Moderate Deficit:</u> Samburu, Garissa, Laikipia, Kilifi, Taita Taveta<br>• <u>Normal Greenness:</u> Turkana, Kajiado, Kitui<br>• <u>Above Normal Greenness:</u> Baringo, Tharaka Nithi, West Pokot, Embu, Makueni, Meru, Nyeri, Kwale, Lamu, Narok | **Counties Affected**: Garissa, Isiolo, Marsabit, Samburu, Tana River, Baringo, Turkana, Kajiado, Kilifi, Meru North, Narok, West Pokot |
| **August 2023** | **Rainfall distribution:**<br>• <u>Pastoral North East Counties:</u> Counties: Mandera, Wajir, Isiolo, Tana River, Garissa received between less than 1mm to 25mm<br>• <u>Pastoral North West Counties:</u> Turkana, Marsabit, Samburu received 1mm to 50mm<br>• <u>South East Marginal Agriculture Counties:</u> Tharaka Nithi, Embu, Kajiado, Meru, Makueni, Kitui received between less than 1mm to 25mm<br>• <u>Agro Pastoral Cluster Counties: Kajiado</u>, Laikipia, Narok, Baringo, Nyeri, West Pokot received between less than 1mm to 50mm<br>• <u>Coast Marginal Agriculture Counties</u>: Kwale, Kilifi, Lamu received between 26mm to 100mm | **Diseases Reported**: Contagious Caprine Pleuropneumonia (CCPP), Contagious Bovine Pleuropneumonia (CBPP), Peste des Petits Ruminants (PPR), Foot and Mouth Disease<br><br>**Counties Affected**: Garissa, Isiolo, Marsabit, Samburu, Tana River, Baringo, Turkana, Kajiado, Kilifi, Meru North, Narok, West Pokot |

| | | |
|---|---|---|
| | **Vegetation Condition**<br>• Deterioration in Vegetation Condition: Noted across ASAL counties compared to July 2023 due to ongoing dry season<br>• Moderate Vegetation Deficit: Samburu, Kajiado, Taita Taveta<br>• Normal Vegetation Greenness: West Pokot, Baringo, Laikipia, Turkana, Marsabit, Mandera, Garissa, Tana River<br>• Above Normal Vegetation Greenness: Narok, Makueni, Kitui, Nyeri, Embu, Tharaka Nithi, Meru, Isiolo, Wajir, Kilifi, Kwale, Lamu | |
| **September 2023** | **Rainfall distribution:**<br>• Pastoral Northeast (e.g., Mandera, Wajir, Isiolo, Tana River, Garissa): Received <1mm to 20mm<br>• Pastoral Northwest (Turkana, Marsabit, Samburu): Received <1mm to 50mm<br>• Southeast Marginal Agriculture: Near to above-average rainfall, except Embu and Nyeri (below average)<br>• Coast Marginal Agriculture (Kwale and Kilifi): Below average rainfall<br><br>**Drought Phases**<br>Normal Drought Phase: 18 counties.<br>Alert Phase: 5 counties-Laikipia, Samburu, Turkana, Tana River, Taita Taveta<br><br>**Vegetation Index**<br>• Vegetation Condition Index (VCI): A slight decline was noted in some ASAL counties compared to August 2023 due to high temperatures<br>• Moderate Vegetation Deficit: Reported in 5 counties (Taita Taveta, Samburu, Turkana, West Pokot, Laikipia)<br>• Normal Vegetation Greenness: Observed in 7 counties (Kilifi, Marsabit, Tana River, Narok, Baringo, Garissa, Kajiado)<br>• Above Normal Vegetation Greenness: Found in 11 counties (Embu, Isiolo, Kitui, Kwale, Lamu, Makueni, Meru, Nyeri, Tharaka Nithi, Wajir, Mandera) | • Endemic diseases: Contagious Bovine Pleural Pneumonia, Contagious Caprine Pleural Pneumonia, Lumpy Skin Disease, Peste des petits ruminants, goat/sheep pox, Heartwater disease, and Mange<br><br>• Suspected Diseases: Foot and Mouth Disease was reported in Embu, West Pokot, Samburu, and Narok counties<br><br>• Tick-Borne Diseases: Cases of anaplasmosis, babesiosis, and East Coast Fever were noted, along with abortions among goats due to suspected Q-Fever disease |
| **November 2023** | **Rainfall Situation** | Diseases reported: Unconfirmed cases of Contagious Caprine Pleuropneumonia (CCPP), Contagious Bovine |

| | | |
|---|---|---|
| | • Impact of Short Rains: The October-December short rains, influenced by the El Niño and a positive Indian Ocean Dipole, resulted in normal conditions across all 23 Arid and Semi-Arid Lands (ASAL) counties<br><br>**Flooding Impacts:**<br>• Counties Affected: ASAL counties, notably in Isiolo, Samburu, Wajir, Garissa, Mandera, and Tana River<br><br>**Vegetation Condition:**<br>• Normal Vegetation Greenness: Samburu, Tana River, Laikipia, Kitui, Garissa, and Kilifi<br>• Above-Normal Vegetation Greenness: Counties included Turkana, West Pokot, Baringo, Narok, Nyeri, Makueni, Embu, Tharaka Nithi, Meru, Isiolo, Marsabit, Wajir, Mandera, Lamu, and Kwale | Pleuropneumonia (CBPP), Peste des Petits Ruminants (PPR), and Foot and Mouth Diseases were reported<br><br>Counties affected: Garissa, Isiolo, Marsabit, Samburu, Tana River, Baringo, Turkana, Kajiado, Kilifi, Meru North, Narok, and West Pokot |
| **December 2023** | **Drought phase classification:**<br>• All ASAL counties are classified under the 'Normal' drought phase due to environmental, production, access, and utilisation indicators falling within usual ranges<br><br>**Rainfall performance**<br>• The first ten days of December experienced reduced rainfall intensity compared to November, leading to the cessation of the October-November-December rains<br>• Many counties received below-normal rainfall, with notable exceptions where some areas, such as parts of Kilifi and Lamu, recorded significantly above-average rain (up to 180% of normal)<br>• The highest total rainfall of over 225 mm was observed in Embu and Taita Taveta counties<br><br>**Vegetation Condition:**<br>• Overall improvement in vegetation conditions was reported across ASAL counties due to cumulative rainfall during the October-December season | • Garissa: Outbreak of sheep and goat pox and suspected foot and mouth disease<br>• Kajiado: There has been an outbreak of sheep and goat pox and suspected foot and mouth disease<br>• Narok: Outbreak of sheep and goat pox and suspected foot and mouth disease<br>• Samburu: An upsurge of listeriosis (circling disease) in goats<br>• Wajir: Outbreak of Rift Valley Fever, 52 of the 275 samples returned positive for RVF<br>• Marsabit: Cases of tsetse flies, especially in the plains of North Horr, were noted, and a rabies outbreak was reported in Tigo |

<table>
<tr><td></td><td><ul><li>By December, all counties exhibited normal or above-normal vegetation greenness, with a notable recovery in Taita Taveta</li></ul></td><td></td></tr>
<tr><td>January 2024</td><td>Rainfall Distribution:<ul><li><u>Northern Sector</u>: Mostly sunny and dry conditions; rainfall reported in isolated areas of Isiolo, Marsabit, Turkana, Kitui, Tana River, Taita Taveta, Kilifi, Lamu, West Pokot</li><li><u>Southern Sector</u>: Rainfall near to above average, with below-average rainfall in the Coastal region and parts of the Southeastern lowlands</li><li><u>Storm Activity</u>: Isolated storms experienced over various highland regions</li></ul>Drought Situation:<ul><li>Following the good performance of the 2023 short rain season, all 23 counties classified as Arid and Semi-Arid Lands (ASALs) were categorised under the ‘Normal’ phase</li></ul>Vegetation Index:<ul><li>Normal Vegetation: All ASAL counties presented normal vegetation greenness due to cumulative rainfall, indicating stable vegetation conditions</li><li>Vegetation Condition Index (VCI) Summary:<ul><li>Extreme (0): 0 counties</li><li>Severe Vegetation Deficit (0): 0 counties</li><li>Moderate Vegetation Deficit (0): 0 counties</li><li>Normal Vegetation Greenness (6 counties): Baringo (Mogotio), Turkana (East, Central, North), Garissa (Balambala, Township)</li><li>Above-normal Vegetation Greenness (20 counties): Includes counties such as Embu, Kajiado, Kilifi, Kitui, Kwale, Laikipia, Lamu, Makueni, Meru, Nyeri, Taita Taveta, Tharaka Nithi, West Pokot, Narok, Mandera, Marsabit, Samburu, Tana River, Turkana, and Wajir</li></ul></li></ul></td><td><ul><li>Rift Valley Fever (RVF): Reported in Marsabit and Wajir counties</li><li>Foot and Mouth Disease: Suspected in Garissa, Kajiado and Narok</li><li>Listeriosis (circling disease): Reported in Waso Ward, Samburu County</li><li>Tsetse Flies: Noted in Marsabit</li><li>Rabies: Outbreak reported in Marsabit County</li></ul></td></tr>
<tr><td>February 2024</td><td>Drought Phase</td><td><ul><li>Outbreaks of Rift Valley Fever in Marsabit and Wajir, with 52 positive samples from Wajir</li></ul></td></tr>
</table>

| | | |
|---|---|---|
| | • General Condition: All counties are classified under the 'Normal' phase based on environmental, production, access, and utilization indicators. This is attributed to the successful performance of the 2023 short rains season<br><br>**Rainfall Performance**<br>• <u>Pastoral North East:</u> Mandera, Wajir, Isiolo, Tana River, Garissa received rainfall between <1mm to 50mm<br>• <u>Pastoral North West:</u> Turkana, Marsabit, and Samburu received rainfall ranging from 11mm to 50mm<br>• <u>South East Marginal Agriculture:</u> Tharaka Nithi, Embu, Kajiado, Meru, Makueni, Kitui counties received rainfall between 11mm to 70mm<br>• <u>Agro-Pastoral Cluster</u>: Kajiado, Laikipia, Narok, Baringo, Nyeri, West Pokot recorded 11mm to 100mm of rainfall<br>• <u>Coastal Marginal Agriculture:</u> Kwale, Kilifi, and Lamu received between 2mm to 50mm<br><br>**Vegetation Condition**<br>February 2024 saw stability in vegetation conditions across ASAL counties compared to January 2024<br><br>• No counties reported extreme, severe, or moderate vegetation deficits<br>• Normal Vegetation: Turkana and West Pokot.<br>• Above Normal Vegetation: 21 counties, including Samburu, Laikipia, Kajiado, Kitui, Tana River, and others | • Foot and Mouth Disease suspected in Garissa, Kajiado, and Narok.<br>• Listeriosis reported in goats in Samburu<br>• Tsetse flies and rabies outbreaks noted in Marsabit<br>• Instances of camel deaths in Mandera with undetermined causes |
| **March 2024** | **Drought Situation:**<br>• The drought situation remains stable across the 23 ASAL counties, classified mainly under the 'Normal' drought phase<br><br>**Rainfall Performance:**<br>Early onset of the March to May (MAM) long rains has been observed, with varying rainfall amounts: | • Reports of endemic diseases (e.g., sheep and goat pox, foot and mouth disease) were reported in Garissa, Kajiado and Narok<br>• Listeriosis (circling) disease in goats was observed in Samburu County.<br>• Outbreaks of Rift Valley Fever in Marsabit and Wajir reported and controlled |

| | | |
|---|---|---|
| | • <u>Pastoral Northeast Counties:</u> 0mm to 125mm (Mandera, Wajir, Isiolo, Tana River, Garissa)<br>• <u>Pastoral Northwest Counties</u>: Trace amounts (Turkana, Marsabit, Samburu)<br>• <u>Southeast Marginal Agriculture Counties:</u> 78mm to 200mm (Tharaka Nithi, Embu, Kajiado, Meru, Makueni, Kitui)<br>• <u>Coastal Counties:</u> Up to 288mm (Kwale, Kilifi, Taita Taveta, Lamu)<br><br>**Vegetation Condition:**<br>• Improvement in vegetation conditions is noted across ASAL counties compared to February 2024<br>• No counties reported severe vegetation deficits, while West Pokot recorded normal greenness; other counties showed above-normal greenness | |
| **April 2024** | **Rainfall distribution**<br>Most ASAL counties received near to above-average rainfall, with some areas experiencing extreme storms<br><br>• Samburu, Isiolo, Wajir, Mandera, Meru, and Baringo received 151-200% of the long-term mean (LTM) for April<br>• Kitui, Makueni, Kajiado, and Nyeri received 201-300% of the LTM<br>• Turkana recorded rainfall exceeding 300% of the LTM<br><br>**Vegetation Condition**<br>• Favourable vegetation conditions were noted across ASAL counties, attributed to good rains and lower land surface temperatures<br>• All counties recorded above-normal vegetation greenness<br>• Vegetation Condition Index (VCI) for April 2024<br> ○ No extreme, severe, or moderate vegetation deficits were reported<br> ○ 23 counties recorded above-normal greenness | • Suspected foot and mouth disease (FMD) cases were noted in Narok and West Pokot counties<br>• Lumpy skin disease (LSD) was reported in Laikipia county<br>• Camel abortions were observed in Garissa and Mandera |

| | | |
|---|---|---|
| **May 2024** | **Drought Situation Overview**<br>• All counties in the Arid and Semi-Arid Lands (ASAL) have been categorised under the 'Normal' phase due to favourable environmental, production, access, and utilisation indicators, primarily from the good performance of the March-May 2024 (MAM) rainfall season<br><br>**Rainfall Performance**<br>Several ASAL counties received above-average rainfall<br>• Pastoral counties like Mandera, Wajir, Isiolo, Tana River, and Garissa received 125mm to 300mm. Wajir received (25mm – 125mm)<br>• Pastoral North West counties, including Turkana, Marsabit, and Samburu, experienced rainfall amounts ranging from 75mm to 300mm<br>• Coastal counties like Kwale, Kilifi, Taita Taveta, and Lamu received lower rainfall amounts (<25mm to 76mm), with Taita Taveta recording the least<br>**Vegetation Condition**<br>• The Vegetation Condition Index (VCI) indicates sustained improvement in vegetation across ASAL counties in May 2024, with no counties reporting extreme, severe, or moderate vegetation deficits<br>• Only Baringo recorded average vegetation greenness, while 22 counties exhibited above-normal greenness due to ongoing MAM 2024 long rains<br>• The overall vegetation condition showed improvement compared to previous months, indicating favourable conditions for pastoral and agricultural activities | • Suspected cases of foot and mouth disease (FMD) in cattle in Narok county<br>• Confirmed cases of lumpy skin disease (LSD) were recorded in Laikipia and Turkana<br>• Alarming occurrences of camel abortions were reported in Garissa, with further investigations ongoing in Marsabit and Mandera<br>• Cattle deaths attributed to suspected vector-borne illnesses were reported in Garissa, with trypanosomiasis, anaplasmosis, and babesiosis suspected as contributing factors<br>• Foot rot disease cases were also reported in Tharaka Nithi |
| **June 2024** | **Rainfall Performance by Regions**<br>• Pastoral North East Counties (Mandera, Wajir, Isiolo, Tana River, Garissa): Less than 25mm of rainfall recorded<br>• Pastoral North West Counties (Turkana, Marsabit, Samburu): Rainfall amounts ranged from 26mm to 75mm<br>• South East Marginal Agriculture Counties (Tharaka Nithi, Embu, Kajiado, Meru, Makueni, Kitui): Mostly dry with trace rainfall amounts<br>• Agro-Pastoral Counties (Kajiado, Laikipia, Narok, Baringo, Nyeri, West Pokot): Received good rainfall amounts | • Foot and Mouth Disease (FMD) is suspected in cattle in Laikipia, Narok Wajir, Samburu and Meru counties<br>• There have been reports of Peste des Petits Ruminants (PPR) in goats in Garissa and Marsabit counties, with high case fatality rates<br>• Increased respiratory and septicemic diseases reported in camels in Marsabit |

| | | |
|---|---|---|
| | • <u>Coast Marginal Agriculture Counties</u> (Kwale, Kilifi, Taita Taveta, Lamu): Received rainfall amounts between 51-76mm<br><br>**Vegetation Condition Index (VCI)**<br>• The VCI remained stable in June compared to May, indicating continued improvement in vegetation conditions across the ASAL counties<br>• None of the counties reported extreme, severe, or moderate vegetation deficits<br>• All 23 ASAL counties reported above-normal vegetation greenness, with no counties classified under a negative vegetation condition category | |
| **July 2024** | **Drought Phase Classification:**<br>• <u>Normal Phase</u>: Twenty-two counties are classified under the 'Normal' drought phase due to favourable environmental and production indicators following good rainfall during the March-April-May (MAM) 2024 season<br>• <u>Alert Phase</u>: Kilifi County is noted to be under an 'Alert' phase, indicating potential concerns despite overall positive trends<br><br>**Rainfall Performance:**<br>• North East Counties (Mandera, Wajir, Isiolo, Tana River, Garissa) received minimal rainfall (2-20 mm)<br>• North West Counties (Turkana, Marsabit, Samburu) experienced moderate rainfall (61-121 mm)<br>• Marginal Agricultural Counties (e.g., Tharaka Nithi, Kajiado, Meru) received trace amounts (2-50 mm)<br>• Coastal Marginal Counties (Kwale, Kilifi, Taita Taveta) also saw minimal rainfall (11-60 mm)<br><br>**Vegetation Condition:**<br>• The Vegetation Condition Index (VCI) remained stable compared to June 2024, with no extreme deficits observed | • Abortions in goats and sheep in Mandera, Turkana and Samburu<br>• Suspected foot-and-mouth disease in Embu, Kitui, Kwale, Laikipia, Lamu, Wajir<br>• Outbreaks of bluetongue disease in sheep in Garissa, Laikipia and Kajiado Counties<br>• Unconfirmed incidences of lumpy skin disease (LSD) were reported in Narok<br>• Samburu reported outbreak of Pest De Petit Ruminants (PPR) in small stock<br>• suspected cases of sudden death syndrome (SDS) in camels were witnessed in Garissa<br>• Q-fever being recorded across Kajiado |

| | | |
|---|---|---|
| | • Above-Normal Greenness: Twenty-one counties, including Samburu, Laikipia, and Turkana, reported above-normal vegetation, reflecting a positive impact from the MAM rains and subsequent JJA rains<br>• Normal Vegetation Greenness: Kilifi and Kwale reported normal conditions, indicating a need for monitoring | |
| **August 2024** | **Rainfall Distribution**<br>• Pastoral North East Counties: Mandera, Wajir, Isiolo, Tana River, and Garissa received 2-50 mm of rainfall<br>• Pastoral North West Counties: Turkana, Marsabit, and Samburu received 51-200 mm of rainfall<br>• South East Marginal Agriculture Counties: Tharaka Nithi, Embu, Kajiado, Meru, Makueni, and Kitui received 2-20 mm of rainfall<br>• Agro-pastoral Counties: Kajiado, Laikipia, Narok, Baringo, Nyeri, and West Pokot received 11-50 mm of rainfall<br>• Coast Marginal Agriculture Counties: Kwale, Kilifi, Taita Taveta, and Lamu received 11-50 mm of rainfall<br><br>**Drought**<br>21 ASAL counties were categorised under the 'Normal' drought phase due to the good performance of the March-May (MAM) 2024 and light June-September (JJAS) rains<br><br>**Vegetation Index**<br>• Vegetation conditions remained stable in August 2024 compared to July 2024<br>• Normal Vegetation Greenness: Kilifi and Kwale<br>• Above Normal Vegetation Greenness: 21 counties including Samburu, Laikipia, Kajiado, Kitui, Turkana, Tana River, Garissa, Baringo, Narok, Nyeri, Makueni, Embu, Tharaka Nithi, Meru, Isiolo, Marsabit, Wajir, Mandera, Taita Taveta, West Pokot, and Lamu<br>• No counties recorded extreme, severe, or moderate vegetation deficit | • Foot and Mouth Disease (FMD): Reported in Kitui and Kajiado counties<br>• Trypanosomiasis: Recorded in Mutha and Kaziku wards in Kitui South Sub- County<br>• Endo and Ecto Parasites: Increased incidences at congested water points in Garissa<br>• Sudden Death Syndrome (SDS) in Camels and Peste des Petits Ruminants (PPR) in Goats: Reported in Garissa County |

| | | |
|---|---|---|
| **September 2024** | **Rainfall Distribution**<br>• Pastoral North East counties (Mandera, Wajir, Isiolo, Tana River, Garissa): Minimal rainfall (2-50 mm)<br>• Pastoral North West counties (Turkana, Marsabit, Samburu): Significant rainfall (51-200 mm)<br>• South East Marginal Agriculture counties (Tharaka Nithi, Embu, Kajiado, Meru, Makueni, Kitui): Very low rainfall (2-20 mm)<br>• Agro-Pastoral areas (Kajiado, Laikipia, Narok, Baringo, Nyeri, West Pokot): Moderate rainfall (11-50 mm)<br>• Coastal Marginal Agriculture counties (Kwale, Kilifi, Taita Taveta, Lamu): Low rainfall (11-50 mm)<br><br>**Vegetation Index**<br>• General Condition: Stability in vegetation across the Arid and Semi-Arid Lands (ASAL) counties compared to August 2024. The vegetation remained above normal greenness, attributed to the cumulative impacts of good MAM 2024 long rains and moderate JJA rainfall.<br>• Above Normal Vegetation Greenness: 21 ASAL counties including Samburu, Laikipia, Kajiado, Kitui, Turkana, Tana River, Garissa, Baringo, Narok, Nyeri, Makueni, Embu, Tharaka Nithi, Meru, Isiolo, Marsabit, Wajir, Mandera, Taita Taveta, West Pokot, and Lamu<br>• Normal Vegetation Greenness: Kilifi and Kwale | • Lumpy Skin Disease (LSD): Reported in cattle in Turkana<br>• Tsetse Fly Infestation: Cross-border areas of Loima and Turkana West<br>• Clostridial Infections (Enterotoxaemia) in Sheep: Reported in Kajiado<br>• Suspected Foot and Mouth Disease (FMD): Reported in Kajiado, Kwale, Kitui, Laikipia<br>• Blue Tongue Disease: Confirmed cases in Laikipia.<br>• Pestes des Petits Ruminants (PPR): Outbreak in sheep and goats reported in Laikipia and West Pokot |

**Table 5.5 Publications on The Effect of Climate-On-Climate-Sensitive Infectious Diseases In Kenya**

| Study | Study objective | Finding |
|---|---|---|

| | | |
|---|---|---|
| **1. Ototo 2015**[86] | To determine the impact of ITNs on indoor vector densities and biting behaviour in western Kenya. | **Seasonal Rainfall Influence**: Densities of *An. gambiae* rise in April-May, about one month after the rains begin, highlighting how rainfall patterns directly affect vector abundance by providing breeding sites. Similarly, *An. funestus* peaks two months after the rains, around June-July.<br><br>**Highland and Lowland Comparison**: *An. gambiae* is more abundant in highland sites, while *An. funestus* and *An. arabiensis* dominate in lowland areas, suggesting that climate at different elevations (altitudes) favors different species.<br><br>**Dry Season Observations**: The study mentions lower mosquito densities in the dry season (January-March) in most sites, except Rae, which displays consistently high *An. gambiae* levels. |
| **2. Nyawanda 2023**[72] | To investigate the relative effect of climate variability on malaria incidence after scale-up of interventions in western Kenya. | **Land Surface Temperature Day (LSTD)**: The study reports a median LSTD of 32.2°C, ranging from 29.5 to 35.2°C. It notes year-to-year changes (e.g., an average decline to 31.9°C by 2013, followed by a rise to 34.8°C in 2017).<br>**Land Surface Temperature Night (LSTN)**: This variable has a median of 18.3°C and is not highly correlated with malaria incidence, so it is excluded from detailed modelling.<br>**Air Temperature**: The median air temperature is 22.9°C, and future projections suggest an increase from 24.4°C to 26.7°C by 2100, indicating potential long-term climate shifts.<br>**Rainfall:** Rainfall is noted to peak from March-May and again in November, with a median of 100 mm and a range of 64–143 mm, showing a strong bimodal seasonality that aligns with malaria incidence peaks. |

| | | |
|---|---|---|
| **3. Otieno 2019**[85] | To analyse and predict forecast-based impacts in Tana River using openly available data on flood exposure, vulnerability, lack of coping capacity, flood impacts and observed satellite rainfall. | The floods caused extensive damage to livelihoods, killing over 6,000 livestock, submerging 8,450 acres of farmland, and damaging homes and infrastructure, including roads.<br><br>Stagnant water from these floods increased the risk of disease outbreaks, including Rift Valley Fever (RVF), a mosquito-borne disease impacting both animals and humans.<br><br>There is a negative correlation (-0.212) between rainfall within the Tana River area and house destruction, suggesting that higher rainfall here is not strongly associated with increased house damage.<br><br>A positive correlation (0.233) between rainfall in the upper catchment areas (Meru and Tharaka counties) and house destruction indicates that more houses tend to be destroyed when there is higher rainfall upstream, contributing to downstream flooding. |
| **4. Hay 2000**[29] | To understand the factors contributing to the interepidemic periods of dengue haemorrhagic fever in Bangkok and Plasmodium falciparum malaria in western Kenya. | **Dengue Haemorrhagic Fever (DHF)**<br>• **Annual Variation**: Accounts for approximately **30%** of the series variance in incidence data.<br>• **Superannual Variation: This accounts for approximately 70% of the series variance, with a periodicity of** around **3 years**.<br>• **Predicted Interepidemic Period**: Approximately **35 months** based on model parameters (D = 5 days, D9 = 0.2 months, A = 84 months).<br>**2. Plasmodium falciparum Malaria**<br>• **Annual Variation**: Accounts for about **70%** of the series variance in incidence data. |

| | | |
|---|---|---|
|  |  | • **Superannual Variation: This accounts** for approximately **30%** of the series variance, with a periodicity also cantered around **3 years**.<br>• **Predicted Interepidemic Period**: Approximately **34 months** based on model parameters (D = 9–10 days, D9 = 9.5 months, A = 3.5 months).<br>**3. Climate Data (Bangkok)**<br>• **Temperature and Rainfall**:<br> ○ Over **90%** of series variance accounted for by annual cycles (12-month periodicity).<br>**4. Climate Data (Kericho, Kenya)**<br>• **Temperature and Rainfall**:<br> ○ **80%** of series variance in temperature and **90%** in rainfall explained by annual cycles (12-month periodicity).<br>**5. MENSOI Index (Climate Indicator)**<br>• **Periodic Peaks**: Show peaks of approximately **4-year periodicity**, with cyclical variations evident but not strongly influencing rainfall and temperature in the studied regions. |
| **5. Sewe 2016**[102] | To explore the lagged association of Normalized Difference Vegetation Index (NVDI), day Land Surface Temperature (LST) and precipitation on malaria mortality in three areas in Western Kenya | **LAND SURFACE TEMPERATURE**<br>• Increased risk of malaria deaths at temperatures below 28°C and above 34°C.<br>• Protective effect at temperatures above 28°C when adjusted for seasonal effects.<br><br>**PRECIPITATION**<br>• Increased malaria risk with rainfall above 20 mm weekly, peaking at 80 mm/week<br>• Higher risk with rainfall below 20 mm/week<br>• Seasonal adjustment shows a higher risk with rainfall above 80 mm/week<br><br>**NORMALISED DIFFERENCE VEGETATION INDEX (NVDI)** |

| | | |
|---|---|---|
| | | • There is a higher risk of malaria deaths with NDVI values below 0.4 (sparse or poor vegetation)<br>• NDVI values above 0.4 (dense, healthy vegetation) are associated with lower malaria mortality<br><br>**MONTHLY VARIATION**<br>• Highest malaria deaths in April<br>• Lowest in September<br>Peak deaths in 2008-2009 and declined in 2010 |
| **6. Campbell 2019** [10] | To compare retrospective predictions across 16 years (2002 to 2018) to observe general differences between predicted values during epizootic and inter-epizootic periods | **MINIMUM WETNESS INDEX VALUES**<br>• Increased mosquito abundance in wetter areas: A wetness index above 0.5 correlates with higher mosquito populations<br><br>**CUMULATIVE PRECIPITATION (0 to 14 days prior)**<br>• Mosquito numbers Increased by 3.5 times, with rainfall exceeding 20 mm/week<br><br>**LAND SURFACE TEMPERATURE (8-day mean composites)**<br>• Every 1°C increase in temperature correlates with a 15% increase in mosquito abundance<br><br>**CUMULATIVE PRECIPITATION (14 to 18 days and 14 to 28 days prior)**<br>• Cumulative precipitation from 14 to 28 days prior shows negligible effects on abundance compared to recent rainfall<br><br>**PERCENTAGE OF CLAY IN SOIL**<br>A decrease of 10% clay in the soil increases mosquito abundance by 12% |
| **7. Gikungu 2016** [22] | To develop a dynamic model for predicting the risk of RVF outbreaks in Kenya with a lead-time of at least three | **NDVI AND RAINFALL CORRELATION**<br>Higher rainfall increases NDVI, enhancing vegetation growth and creating more habitats for RVF vectors.<br>• **Garissa Model:** Significant predictors included NDVI (-10.285, P=0.0256) and rainfall (-7.279, P=0.0295). |

| | | |
|---|---|---|
| | months in epizootic areas of the country. | **MINIMUM TEMPERATURES**<br>Significant in all models, indicating that warmer conditions favour mosquito larvae development.<br><br>**HUMIDITY**<br>Higher relative humidity was significant in most models, supporting RVF vector proliferation.<br>• **Garissa Model:** RH12 (relative humidity at 12:00 GMT) had an estimated 34.972 (P=0.0449).<br>• **Kwale Model:** RH06 (relative humidity at 06:00 GMT) showed a significant pessimistic estimate of -55.101 (P=0.009<br><br>**SEA SURFACE TEMPERATURE (SST)**<br>SST was significant for Garissa (15.270, P=0.0387) and Kwale (364.209, P=0.018) but not for Thika, likely due to geographical proximity to the Indian Ocean |

**Table 6: Enablers Barriers to Implementing Advance warning and response systems**

| SURE Level | Key enabler themes | Key barrier themes |
|---|---|---|
| **Enablers and barriers at the patient, population or community level** | | |
| **Knowledge** | 1. Adequate and regular training of participants on the AW&R system, its processes and early warning signs[12,22,37,89,126] | 1. Poor health-seeking behaviour among participants[27,84,120] Long intervals between data collection affect symptom recall bias[16] |
| **Attitude regarding warning and response system acceptability, appropriateness, credibility, simplicity, flexibility and adaptability** | 1. Dissemination of surveillance findings encourages system acceptance[132] | 1. Fear of foreign healthcare workers inhibits involvement[80]<br>2. Fear of disease stigmatisation upon involvement in surveillance[139]<br>3. Reliance on traditional medicine hinders full participation[139] |
| **Motivation to change or adopt a new behaviour** | 1. Availability of timely professional medical care[114,115]<br>2. Community involvement[63]<br>3. Free SMS for data collection and reporting[126] | 1. High cost of healthcare treatment hinders involvement[84]<br>2. HIV/AIDS-related stigma hinders involvement due to fear of being known[95]<br>3. Delayed payments of incentives/allowances[126] |
| **Enablers and barriers at the providers of care level** | | |
| **Knowledge skills and experience** | 1. Training on data collection, management and analysis at the health centre[1,14,34,37,40,61,63,64,66,84,117,119,136,157]<br>2. The use of competent professionals[3,81]<br>3. Improved diagnostic methods[81]<br>4. Use of healthcare workers with increased expertise based on increased years of experience[118] | 1. Low disease awareness and suspicion index, especially for NTDs[24,27,122,128]<br>2. Poor understanding of the AW&R system functionality and steps[43,63,159]<br>3. Inadequate training and skills for disease identification, management or preparedness[81,118,143]<br>4. Poor adherence to medical and AW&R system guidelines[95,118]<br>5. Underreporting of findings[24] |

| | | |
|---|---|---|
| **Attitude regarding warning and response system acceptability, appropriateness, credibility, simplicity, flexibility and adaptability** | 1. Positive perceptions due to benefits of the system in early disease detection[117]<br>2. Positive experiences due to ease of use of technological systems[34]<br>3. Personal self-motivation and passion towards healthcare[12]<br>4. Dissemination of surveillance findings[92] | - |
| **Motivation to change or adopt a new behavior** | 1. Enhanced sense of ownership from understanding the system, its data and the procedures[34]<br>2. Simplicity and ease of use of the AW&R system technology[34,58,117] | 1. Inadequate healthcare supplies cause demotivation[12]<br>2. Lack of feedback and follow-up across stakeholders regarding findings or adverse events, respectively[58,113]<br>3. Delayed or lack of financial incentive[117]<br>4. Demotivated managers/supervisors negatively affect lower cadres[117] |
| **Enablers and barriers at the other stakeholders' level (including community health committees, community leaders, program managers, donors, policymakers and opinion leaders)** | | |
| **Knowledge and skills** | 1. Multi-stakeholder collaboration[39,134] | 2. Lack of specialised personnel at the hospital facility, e.g. data analysts[159]<br>3. Reliance on clinical assessment than laboratory assessment for disease diagnosis among most farmers[21] |
| **Attitude regarding advance warning and response system acceptability, appropriatenes s and credibility** | 1. Involvement of communities and health workers in surveillance processes[3,25]<br>2. Stakeholder collaboration in choosing CHWs[44]<br>3. Recognition of the burden of disease and the importance of its control[48] | - |

| | | |
|---|---|---|
| **Motivation to change or adopt a new behaviour** | 1. Availability of high-quality equipment[97]<br>2. Availability of training opportunities[94]<br>3. Involvement of external partners[97] | - |
| **Enablers and barriers at the health system constraints level** | | |
| **Accessibility of care** | 1. Improved healthcare infrastructure, facilities and supplies for the implementation of the AW&R system[1,3,9,76,79,92,97]<br>2. Use of more proximal specimen drop-off points to reduce turnaround time[5] | 1. Reduced health-seeking behaviour due to transport costs and remote location of facilities[64,84,110]<br>2. Halting of healthcare services due to the COVID-19 pandemic (e.g. immunisation, family planning, MCH)[3]<br>3. Repurposing of hospital units (e.g. maternity wards) to COVID-19 isolation units[3] |
| **Financial resources** | 1. Increased funding to support AW&R activities[14,63,64]<br>2. Use of technological methods to reduce costs, e.g. Point-of-capture digitisation and use of open-source and free applications[129,133]<br>3. Use of toll-free numbers for disease reporting[115]<br>4. Use of passive surveillance methods, such as collecting data at health facilities, ensuring year-round data collection[6] | 1. Inadequate financial resources for procurement and payments of staff[11,12,18,28,34,40,42,44,55,58,64,66,68,78,113,117,122,126,139,143,159]<br>2. Increased costs of modern, more-acceptable and user-friendly systems and their equipment[3,11,18,28,34,42,44,66,68,78,117,122,126]<br>3. Cost constraints due to long-term sequential data collection in AW&R systems[1]<br>4. High cost of programming due to the rugged terrain (poor roads and insecurity)[3] |
| **Human resources** | 1. A sufficient number of skilled and trained healthcare personnel[1,39,56,58,64,68,80,81,84,97,118,123,135]<br>2. Periodic refresher training on AW&R systems[58,81,116]<br>3. Use of a multi-disciplinary team[14] | 1. Under-trained healthcare staff[21,52,61,78,92,94,129,143,159]<br>2. Reduced number of healthcare workers[12,20,95,122]<br>3. High turnover of healthcare staff with new staff having limited knowledge and skills[23,82]<br>4. Regular industrial strikes from healthcare workers [66,129]<br>5. Brain-drain of skilled personnel seeking greener pastures abroad[139]<br>6. Negligence such as underreporting, incompleteness or inaccuracies in data[94] |

| | | |
|---|---|---|
| | | 7. Insecurity for staff working in conflict regions[122]<br>8. Insufficient supervision of staff[160] |
| **Education system** | 1. Improved education and training curriculum to include teaching on AW&R systems[1,3,12,14,62,66,123]<br>2. Regular updating of surveillance guidelines and education material[62] | 1. Low education level of healthcare workers negatively influences acceptance of surveillance systems[63]<br>2. Low suspicion index due to assuming a region is low risk[27] |
| **Internal communication** | 1. Adhering to formal lines and hierarchical protocols of communication and reporting[3,126,129]<br>2. Timely and regular feedback across the system's hierarchy[113]<br>3. Regular touch-base meetings[34]<br>4. Free or low-cost internet/ airtime for communication[126]<br>5. Availability of a rapid response team[61] | 1. Missing or inconsistent data[10,136]<br>2. Uncertainty on information to report[12]<br>3. Delayed reporting of findings[159]<br>4. Poor communication methods and frequency among internal staff[20] |
| **External communication** | 1. Use of media channels to disseminate findings to the public[41,57]<br>2. Provision of regular feedback to all stakeholders and participants[14]<br>3. Creation of accessible and user-friendly information-sharing channels[78] | 1. Undefined communication and reporting channels with the systems[8,113]<br>2. Poor feedback mechanisms across the different levels of surveillance[12] |
| **Accountability** | 1. Clearly articulated roles, responsibilities and deliverables[1,34] | 1. Lack of coordination and overlapping efforts[12,139]<br>2. Incomplete surveillance coverage[25] |
| **Management, leadership, governance or political will** | 1. Availability of regular and consistent supervision[34,84,118]<br>2. Co-designing systems among different leadership stakeholders allows for a sense of ownership and responsibility[3,97]<br>3. Strong management structures for successful leadership[1]<br>4. Comprehensive supervisory and managerial training[118]<br>5. Political good-will to support AW&R implementation and acceptance[14] | 6. Inadequate and inconsistent supervisory support in running the AW&R system[56,58,84,104,117]<br>7. Unclear and overlapping roles and responsibilities[12]<br>8. Devolution slows down transmission of data (1 study)<br>9. Power struggles between local government and NGOs[139]<br>10. Lack of government support[159] |
| **Information systems** | 1. Utilisation of digital-based methods in data collection, storage and reporting[21,24,34,52,56,66,92,104,108,115,116,132,134]<br>2. Timely collection and dissemination of AW&R data[60,91,102,123,157] | 1. Insufficient reporting and documentation of findings[12,23,56,63]<br>2. Fragmented data in different registries and systems, hindering consolidation and linking of data, e.g. for zoonoses[23,113,136] |

| | | |
|---|---|---|
| | 3. Integration of AW&R systems for generalizability of data, e.g., in the case of zoonotic diseases[1,48,56,122,126]<br>4. Use of modern disease mapping methods, e.g. geo-referencing and remote satellite sensing[6,91,102]<br>5. Availability of technological equipment, e.g. computers, phones, tablets and internet connectivity[108]<br>6. Regular data capturing using the reporting system to avoid missing cases[107] | 3. Outdated (paper-based) information systems challenged receiving real-time information[23,95]<br>4. Poor generalizability due to focusing on illnesses within a specific region only[30]<br>5. Underutilisation of data from AW&R systems [41]<br>6. Technological hindrances, e.g. power fluctuations, network downtime, and power outages[117]<br>7. Challenges in the long-term sustainability of the computers and databases[23]<br>8. Inaccessible health and medical records at the hospital[143]<br>9. Reduced coverage of systems in rural areas[41] |
| **Facilities (such as laboratory infrastructure and others the use of the system)** | 1. Increased number of well-equipped and stocked health facilities and laboratories[22,44,62,64,80]<br>2. Use of state-of-the-art laboratory equipment for faster linkages and diagnosis[136]<br>3. Reduced laboratory turnaround time due to improved facilities[18,92]<br>4. Adequate WASH facilities[64] | 1. Limited equipment, supplies, machinery and infrastructure in the health facilities[42,78,104,120]<br>2. Lack of state-of-the-art laboratory equipment, tests and supplies necessitating samples to be sent abroad[104,139]<br>3. Lack of adequate water, sanitation and hygiene facilities[12,36] |
| **Procurement and distribution systems, including supplies, medicines and technology** | 1. Stable supply chain of surveillance supplies[56,63,64,80,84,92,117] | 1. Shortages of surveillance supplies[6,12,15,43,56,64,78,92,94,97,117,120,122,143]<br>2. Multi-staged procurement steps delayed the running of the AW&R system[94]<br>3. Theft of AW&R supplies[117] |
| **Incentives** | 1. Provision of incentives in the form of money, fuel or airtime[1,23,52,94,114]<br>2. Timely response by the healthcare team to disease events[34] | 1. Low turn-around time of reports due to lack of incentives for healthcare workers[116] |
| **Bureaucracy** | 1. Setting up of government disease units to facilitate early engagement with AW&R systems[123] | 1. Sub-division of government ministries into divisions hinders collaboration[78,139]<br>2. Poor inter-county hospital referral systems[78] |

| | | |
|---|---|---|
| | | 3. Complicated administrative procedures impede hospital surveillance studies[78] |
| **Enablers and barriers at the social and political constraints level** | | |
| **Legislation or regulations (laws, regulations and policies that enabled the implementation of the system)** | 1. Availability of legislation (acts of parliament) provides a coordinated manner of executing the AW&R system[113] | 1. Lack of clarity in policies on emergency response, affecting service provision[78]<br>2. Lack of a national tracking system for AW&R guidelines and policy implementation [95]<br>3. Limited access to AW&R guidelines at the community and facility level[118] |
| **Donor policies** | 1. Financial and technical support from donors during the running of the systems[78] | 1. Fluctuating donor funding [78]<br>2. Donor influence on national and county AW&R prioritisation[113] |
| **Political stability** | - | 1. Political instability and protracted conflict hinder the accessibility and availability of healthcare[3,113,130,139] |

160. World Health Organization. WHO Report on Global Surveillance of Epidemic-prone Infectious Diseases - Introduction. Published online 2019.
