## Supplemental file 2_Annexes for "Advance Warning and Response Systems in Kenya: A Scoping Review"

### Supplementary file 2

### Annexes

**Annex 1: Full search strategy**

Ovid MEDLINE(R) ALL <1946 to August 26, 2024>

| | |
|---|---|
| 1 | Kenya/ 20468 |
| 2 | Kenya*.mp. |
| 3 | Africa, Eastern/ |
| 4 | East Africa*.mp. |
| 5 | 1 or 2 or 3 or 4 |
| 6 | (advance warning* or early warning*).mp. |
| 7 | epidemiological monitoring/ or sentinel surveillance/ |
| 8 | Response system*.mp. |
| 9 | Communicable Disease Control/ or epidemic intelligence.mp. |
| 10 | AW?R.mp. |
| 11 | Integrated Disease Surveillance.mp. |
| 12 | Disease* Surveillance.mp. |
| 13 | Pandemic Preparedness/ |
| 14 | preparedness.ti,ab. |
| 15 | IDSR.mp. |
| 16 | Public Health Surveillance/ |
| 17 | 6 or 7 or 8 or 9 or 10 or 11 or 12 or 13 or 14 or 15 or 16 |
| 18 | 5 and 17 |
| 19 | (event based surveillance or EBS).mp. |
| 20 | 5 and 19 |
| 21 | alert management.mp. |
| 22 | outbreak* response*.mp. |
| 23 | Disease Outbreaks/ |
| 24 | wastewater management.mp. or Environmental Monitoring/ |
| 25 | 21 or 22 |
| 26 | 5 and 25 |
| 27 | 5 and 23 |
| 28 | 5 and 24 |
| 29 | 18 or 20 or 26 or 28 |

**Annex 2: Data Extraction Form**

| **Questions** |
|---|
| **1. General study details** |
| Study title |
| Lead author's surname |
| Publication year |
| Study objectives |
| Article type |
| Overall study design<br>If quantitative, please select the specific study design |
| Study funding |
| Total number of participants |
| Age range of participants |
| **2. Target disease** |
| Name of disease(s) under surveillance |
| Type of target disease |
| Is the disease climate-sensitive? |
| If yes, in the above question, select the climate-sensitive disease |
| **3. Context** |
| Geographical location |
| Implementation level of the AW&R system |
| County |
| **4. Concept** |
| Name of the AW&R system |
| Objective of the AW&R system |
| Description of the AW&R system |
| Implementation dates of the AW&R system |
| Type of the AW&R system |
| AW&R system purpose and scope |
| Alert mechanisms |
| Sources of data |
| Key performance indicators and evaluation frameworks of the AW&R system |
| Lessons learnt (If reported) of the AW&R system |
| To what extent has the AW&R system been successful |
| **5. Enablers and Barriers to successful AW&R systems** |
| Barriers and facilitators at the patient or population or community level |

| |
|---|
| Barriers and facilitators at the providers of care level |
| Barriers and facilitators at the other stakeholders' level (including community health committees, community leaders, program managers, donors, policy makers and opinion leaders) |
| Barriers and facilitators at the health system constraints level |
| Barriers and facilitators at the social and political constraints level |
| Any other barrier or facilitator not captured in any of the five SURE domains above |

**Annex 3: PRISMA-ScR Checklist for Scoping Reviews**

| SECTION | ITEM | PRISMA-ScR CHECKLIST ITEM | REPORTED ON PAGE # |
|---|---|---|---|
| **TITLE** | | | |
| Title | 1 | Identify the report as a scoping review. | Cover Page |
| **ABSTRACT** | | | |
| Structured summary | 2 | Provide a structured summary that includes (as applicable): background, objectives, eligibility criteria, sources of evidence, charting methods, results, and conclusions that relate to the review questions and objectives. | 2 |
| **INTRODUCTION** | | | |
| Rationale | 3 | Describe the rationale for the review in the context of what is already known. Explain why the review questions/objectives lend themselves to a scoping review approach. | 3-4 |
| Objectives | 4 | Provide an explicit statement of the questions and objectives being addressed with reference to their key elements (e.g., population or participants, concepts, and context) or other relevant key elements used to conceptualise the review questions and/or objectives. | 5 |
| **METHODS** | | | |
| Protocol and registration | 5 | Indicate whether a review protocol exists; state if and where it can be accessed (e.g., a Web address); and provide registration information, including the registration number if available. | 5 |
| Eligibility criteria | 6 | Specify characteristics of the sources of evidence used as eligibility criteria (e.g., years considered, language, and publication status), and provide a rationale. | 5 |
| Information sources* | 7 | Describe all information sources in the search (e.g., databases with dates of coverage and contact with authors to identify additional sources) and the date the most recent search was executed. | 6 |
| Search | 8 | Present the full electronic search strategy for at least 1 database, including any limits used so that it could be repeated. | Annex 1 |
| Selection of sources of evidence† | 9 | State the process for selecting sources of evidence (i.e., screening and eligibility) included in the scoping review. | 6 |
| Data charting process‡ | 10 | Describe the methods of charting data from the included sources of evidence (e.g., calibrated forms or forms that the team has tested before their use, and whether data charting was done independently or in duplicate) and any processes for obtaining and confirming data from investigators. | 6 |
| Data items | 11 | List and define all variables for which data were sought and any assumptions and simplifications made. | 6 |
| Critical appraisal of individual sources of evidence§ | 12 | If done, provide a rationale for conducting a critical appraisal of included sources of evidence; describe the methods used and how this information was used in any data synthesis (if appropriate). | Not applicable |
| Synthesis of results | 13 | Describe the methods of handling and summarising the data that were charted. | 7 |
| **RESULTS** | | | |
| Selection of sources of evidence | 14 | Give the number of sources of evidence screened, assessed for eligibility, and included in the review, with reasons for exclusions at each stage, ideally using a flow diagram. | 8 |

| SECTION | ITEM | PRISMA-ScR CHECKLIST ITEM | REPORTED ON PAGE # |
|---|---|---|---|
| Characteristics of sources of evidence | 15 | For each source of evidence, present characteristics for which data were charted and provide the citations. | 8-9 |
| Critical appraisal within sources of evidence | 16 | If done, present data on critical appraisal of included sources of evidence (see item 12). | Not applicable |
| Results of individual sources of evidence | 17 | For each included source of evidence, present the relevant data that were charted that relate to the review questions and objectives. | 10 |
| Synthesis of results | 18 | Summarise and/or present the charting results as they relate to the review questions and objectives. | 12-16 |
| **DISCUSSION** | | | |
| Summary of evidence | 19 | Summarize the main results (including an overview of concepts, themes, and types of evidence available), link to the review questions and objectives, and consider the relevance to key groups. | 17 |
| Limitations | 20 | Discuss the limitations of the scoping review process. | 19 |
| Conclusions | 21 | Provide a general interpretation of the results with respect to the review questions and objectives, as well as potential implications and/or next steps. | 19 |
| **FUNDING** | | | |
| Funding | 22 | Describe sources of funding for the included sources of evidence, as well as sources of funding for the scoping review. Describe the role of the funders of the scoping review. | 20 |
